# Individual variation in susceptibility or exposure to SARS-CoV-2 lowers the herd immunity threshold

**DOI:** 10.1101/2020.04.27.20081893

**Authors:** M. Gabriela M. Gomes, Marcelo U. Ferreira, Rodrigo M. Corder, Jessica G. King, Caetano Souto-Maior, Carlos Penha-Gonçalves, Guilherme Gonçalves, Maria Chikina, Wesley Pegden, Ricardo Aguas

## Abstract

Individual variation in susceptibility and exposure is subject to selection by natural infection, accelerating the acquisition of immunity, and reducing herd immunity thresholds and epidemic final sizes. This is a manifestation of a wider population phenomenon known as “frailty variation”. Despite theoretical understanding, public health policies continue to be guided by mathematical models that leave out considerable variation and as a result inflate projected disease burdens and overestimate the impact of interventions. Here we focus on trajectories of the coronavirus disease (COVID-19) pandemic in England and Scotland until November 2021. We fit models to series of daily deaths and infer relevant epidemiological parameters, including coefficients of variation and effects of non-pharmaceutical interventions which we find in agreement with independent empirical estimates based on contact surveys. Our estimates are robust to whether the analysed data series encompass one or two pandemic waves and enable projections compatible with subsequent dynamics. We conclude that vaccination programmes may have contributed modestly to the acquisition of herd immunity in populations with high levels of pre-existing naturally acquired immunity, while being critical to protect vulnerable individuals from severe outcomes as the virus becomes endemic.

**Graphical Abstract:** 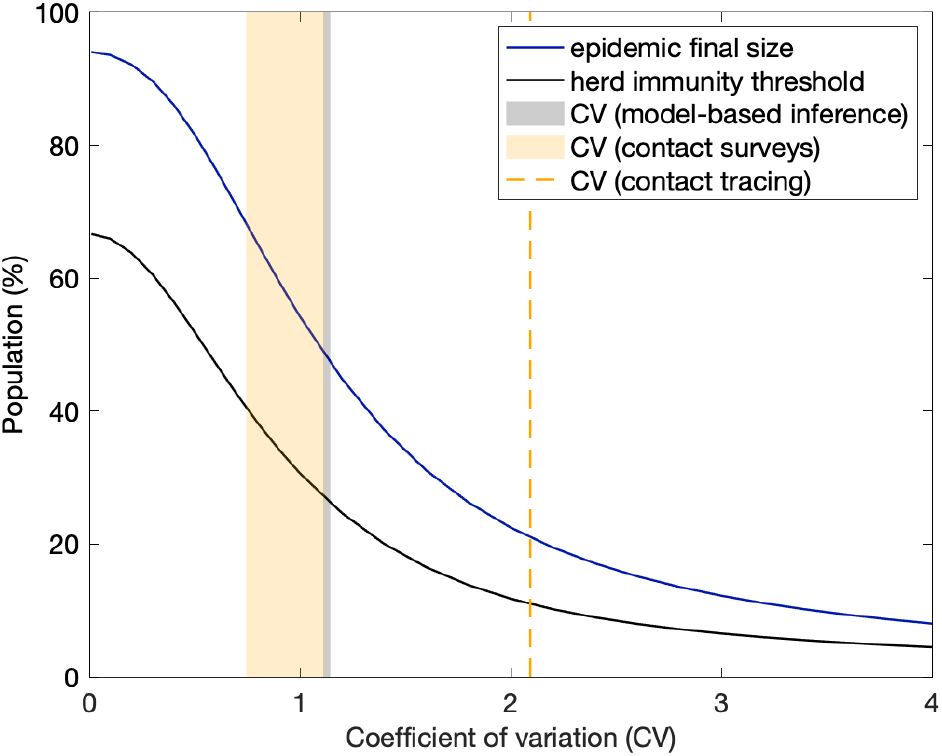

**Highlights:**

- Variation in susceptibility/exposure responds to selection by natural infection
- Selection on susceptibility/exposure flattens epidemic curves
- Models with incomplete heterogeneity overestimate intervention impacts
- Individual variation lowered the natural herd immunity threshold for SARS-CoV-2

## 1. Introduction

Almost 100 years ago, Kermack and McKendrick [Kermack and McKendrick 1927] published a general and flexible model to study epidemics and fitted particular versions to observed epidemics [Kermack and McKendrick 1927, McKendrick 1939]. Already back then they alerted for the simplifying assumption “that all infected persons are equivalent, and that all susceptible persons are equally liable to acquire infection” [McKendrick 1939]. In their fittings they adjust not only transmission parameters but also the size of the susceptible and exposed populations at epidemic onset. Susceptible and exposed population sizes needed to be adjusted so their homogeneous models could fit the data.

Thirty years later, [Gart 1968] admited that “it is difficult to define exactly the size of the population of susceptible hosts. In this instance the difficulty is associated with the heterogeneous nature of the population”. The author divided the population in two groups, depending on their history of infection, and allowed much greater susceptibility in the group with no history. This did not seem sufficient to provide good fit to observed epidemics as the author adds “we assume that the first group is a homogeneous group of susceptibles, while the second is actually a mixture of immune and susceptible individuals”. In [Gart 1971] the author extended the model to several susceptibility groups, and, more than a decade later, [Ball 1985] compared a model with several susceptibility groups to the homogeneous version and described how homogeneity assumptions increase epidemic size. [Coutinho *et al*. 1999] expanded the formalisms to also account for individual variation in incubation period and recovery but concluded that “at present practical applications might be difficult”. [Pastor-Satorras and Vespignani 2001] developed related formalisms to describe epidemics on contact networks.

Meanwhile, frailty variation had been formalised in demography [Vaupel *et al*. 1979] and introduced in practical survival analysis [Hougaard 1984, Aalen 1988] and non-communicable disease epidemiology to improve model fits and interpretation (for a review see [Aalen *et al*. 2015] and references therein).

On the experimental side, [Dwyer *et al*. 1997] measured nonlinear relationships between transmission and densities of susceptible hosts, implying that the bilinear term in the classical susceptible-infected-recovered (SIR) model may not be appropriate. The authors attributed the nonlinearity in transmission to heterogeneity in host susceptibility to infection which they estimated from the shapes of dose-response curves. [Finkenstädt and Grenfell 2000] fitted a model with nonlinear relationships between transmission and density of susceptible hosts to an observed epidemic and estimated the exponent, which they interpreting as heterogeneity in mixing. [Novozhilov 2008] derived the expressions for the exponents from explicit gamma distributions of susceptibility and [Montalbán *et al*. 2020] provided a more tractable derivation that applies exactly to heterogeneity in susceptibility and approximately to heterogeneity in exposure to infection.

Here we build on this history and analyse the coronavirus disease (COVID-19) pandemic, caused by the severe acute respiratory syndrome coronavirus 2 (SARS-CoV-2), with frailty variation models. The study is focused on England and Scotland, where the infection was first detected in early 2020, with major waves of the disease occurring in the spring of the same year and in the autumn-winter season of 2020-2021.

We use susceptible-exposed-infected-recovered (SEIR) models [Diekmann *et al*. 2013] incorporating continuous distributions of individual susceptibility or exposure to infection [Ball 1985, Pastor-Satorras and Vespignani 2001, Novozhilov 2008, Katriel 2012, Gomes 2020]. We then use Bayesian inference to estimate model parameters by fitting series of deaths while accounting for, and estimating the magnitude of, the combined effects of non-pharmaceutical interventions (NPIs), voluntary behavioural change, seasonality, viral evolution, and any other factors which might have contributed to time dependency in transmission potential. We estimate basic reproduction numbers (ℛ_0_) consistent with early literature [Flaxman *et al*. 2020, Hilton *et al*. 2020, Kwok *et al*. 2020] and coefficients of variation in agreement with direct measurements based on contact-pattern studies [Mossong *et al*. 2008, Hens *et al*. 2009, Willem *et al*. 2012]. We show that individual variation in susceptibility or exposure to infection can significantly affect basic metrics, such as the herd immunity thresholds (HIT), as well as model projections and assessment of intervention impacts.

## 2. Mathematical models

The basic compartmental SEIR model [Diekmann *et al*. 2013] describing the transmission dynamics of SARS-CoV-2 is represented diagrammatically in Fig. 1. The model accounts for individual variation in susceptibility or exposure to infection.

**Figure 1:**
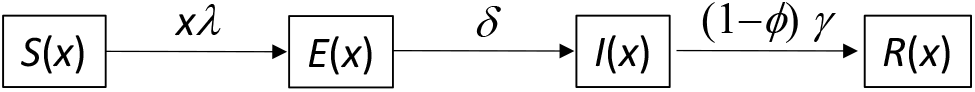
Susceptible-exposed-infected-recovered (SEIR) model representing the transmission dynamics of SARS-CoV-2 in a heterogeneous host population. Individual susceptibility or exposure to infection is denoted by *x*.

### 2.1. Individual variation in susceptibility to infection

Let *x* denote the individual susceptibility to infection in relation to the mean, which we describe by a continuous distribution *q*(*x*) with mean *∫xq*(*x*)*dx* = 1 parametrised by a coefficient of variation (CV), 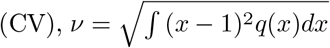. Susceptible individuals, *S*(*x*), become exposed at a rate that depends on their susceptibility, *x*, and on the average force of infection, *λ*, which accounts for the total number of infectious individuals in the population over time. Upon exposure, susceptible individuals enter an incubation phase, *E*(*x*), during which they gradually become infectious [Wei *et al*. 2020, To *et al*. 2020, Arons *et al*. 2020, He *et al*. 2020]. The average infectiousness in this phase is made to be half that in the following state (*ρ* = 0.5), to which individuals progress within an average of 5.5 days (*δ* = 1*/*5.5 per day) [McAloon *et al*. 2020, Lauer *et al*. 2020]. The fully infectious state is denoted by *I*(*x*). Infected individuals eventually become noninfectious, on average approximately 4 days after becoming fully infectious (*γ* = 1*/*4 per day) [Nishiura *et al*. 2020, Li *et al*. 2020]. A small fraction, *ϕ*(*x*), die due to COVID-19 while the remaining majority recover into *R*(*x*) where they are noninfectious and resistant to reinfection due to acquired immunity. The model is formalised mathematically by the infinite system of ordinary differential equations (ODEs):

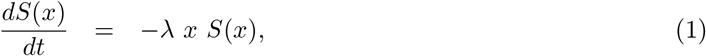

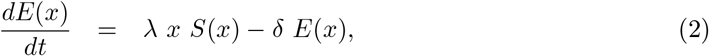

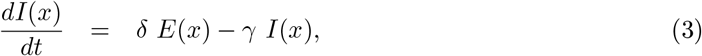

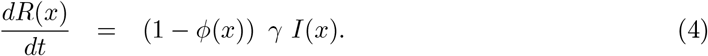

The average force of infection upon susceptible individuals in a population of size N and transmission coefficient *β* is defined by

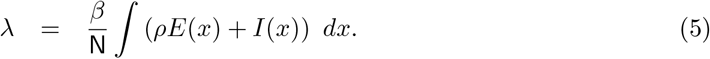

An epidemic is simulated by introducing a small seed of infectious individuals in a susceptible population. Initial growth of infected densities is near exponential but decelerates as individuals are removed from the susceptible pool by infection and immunity. With variation in susceptibility, highly susceptible individuals tend to be infected earlier, leaving behind a residual pool of lower mean susceptibility. This selective depletion intensifies the deceleration of epidemic growth and gives an efficient head start to the acquisition of population immunity. Eventually the epidemic will subside and the HIT, defining the percentage of the population that needs to be immune to reverse epidemic growth and prevent future waves, is lower when variation in susceptibility is higher.

The basic reproduction number, defined as the number of infections caused by an average infected individual in a totally susceptible population, is written for system (1)-(4) with force of infection (5) as

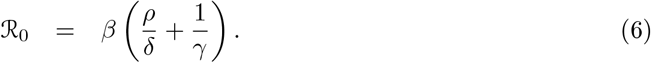

This is an indicator of early transmissibility but its use quickly becomes cumbersome. Several factors, such as NPIs, adaptive changes in human behaviour, seasonality and viral evolution, affect ℛ_0_ in a time-dependent manner. We denote the resulting quantity by ℛ_c_(*t*) = *c*(*t*) · ℛ_0_, where *c*(*t*) *>* 0 describes the basic risk of infection at time *t* in relation to baseline.

In the estimation of ℛ_c_(*t*) we assume a stylised profile for *c*(*t*) as illustrated in Fig. 2: *T*_0_ is time when ℛ_0_ begins to show decrease due to behavioural change or seasonality; *L*_1_ is the period of maximal transmission reduction due to a first lockdown in spring 2020 (48 days in England, from 26 March to 12 May; and 66 days in Scotland, from 24 March to 28 May) and *c*_1_ ≤ 1 is the value of ℛ_c_(*t*) during *L*_1_ in relation to the initial ℛ_0_; *T*_1_ (*> T*_0_) is the day first lockdown begins (transmission is allowed to decrease linearly between *T*_0_ and *T*_1_). After *L*_1_, contact restrictions are gradually relaxed and we allow transmission to begin a linear increase such that *c*(*t*) reaches the baseline (*c*(*t*) = 1) in *T*_2_ days, which may or may not be within the range of this study. Changes in non-behavioural factors that affect transmission (such as seasonality or viral evolution) are inseparable from contact changes in this framework and are also accounted for by *c*(*t*).

**Figure 2:**
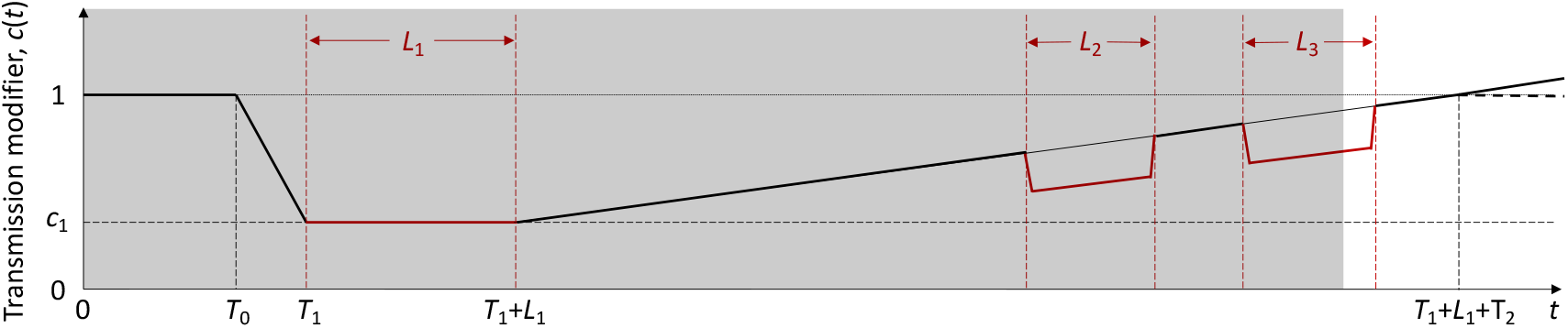
Schematic illustration of the factor *c*(*t*), representing the combined effects of NPIs, adaptive behavioural changes, seasonality and viral evolution on the reproduction number. *L*_1_, *L*_2_ and *L*_3_ represent the known durations (and timings) of first, second and third lockdowns, respectively, as imposed by governments. The gray area illustrates the time period included in our analyses, with a note that the third set of restrictions was still in place when the data fitting period ended. *T*_0_ is the number of days in the series prior to gradual contact reductions early in the pandemic (estimated). *T*_1_ (> *T*_0_) is the day first lockdown begins (informed by data). *T*_2_ is the number of days for the ramp of increasing transmission after first lockdown to return to baseline *c*(*t*) = 1 (technically, this is estimated and used to define the slope of the linear increase rather than to imply that it will continue to follow the trend beyond the study period). Whether *c*(*t*) is allowed to increase beyond baseline (solid) or not (dashed) does not affect our results as we consistently find the factor to remain below baseline throughout the study period.

The model will be used to analyse COVID-19 deaths recorded over approximately one year. Mathematically the time-dependent *c*(*t*) is constructed as

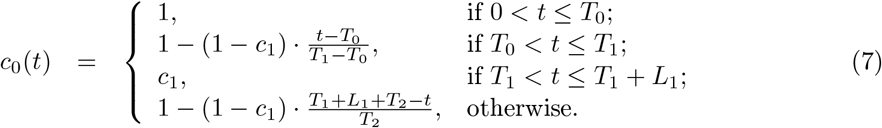

Second and third lockdowns in the autumn and winter are implemented more simply as a further reduction in transmission (by a factor *c*_2_) over the stipulated time periods. Specifically,

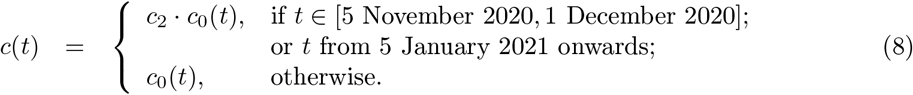

A finite version of system (1)-(4) with variable susceptibility force of infection (5) can be derived exactly, taking a previously obtained form when individual susceptibility is gamma distributed [Novozhilov 2008, Montalbán *et al*. 2020]:

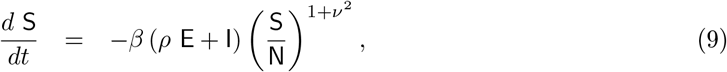

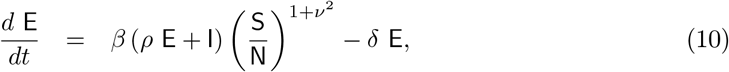

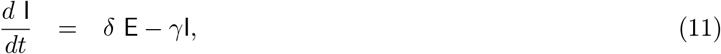

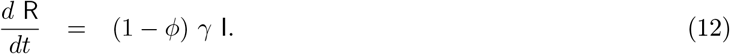

Notice that individuals leave compartment I at a rate *γ* but only a fraction 1 − *ϕ* recovers from infection. The remaining fraction, *ϕ*, die due to COVID-19, *η* days after leaving compartment I. We refer to *ϕ* as the infection fatality ratio (IFR).

Had ℛ_0_ remained constant throughout study duration, the effective reproduction number would have been 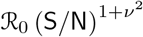 as in [Montalbán *et al*. 2020]. More typical, however, is for ℛ_0_ to vary with seasonal effects [Nickbakhsh *et al*. 2020, Kissler *et al*. 2020, Bacaër and Gomes 2009] and viral evolution [PHE 2021] leading to an effective reproduction number modified as

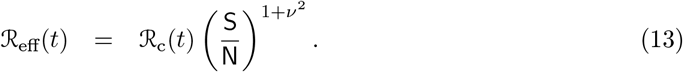

Here, nevertheless, we present HIT estimates elicited by natural infection in reference to the original ℛ_0_, as derived in [Montalbán *et al*. 2020]

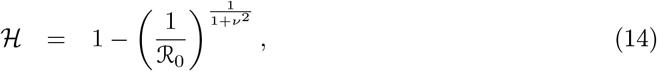

highlighting that these refer to the SARS-CoV-2 of early 2020; higher thresholds will be expected for more transmissible variants. Once reliable estimates become available for evolving transmissibility, HIT estimates can be updated.

As infection spreads, the susceptible compartment, S, is depleted and recovered individuals populate compartment R where they are protected by acquired immunity. In reality, they eventually lose that protection as immunity wanes or is evaded by new viral variants. In addition, the population slowly renews itself through all-cause death of individuals with various levels of acquired immunity and birth of new susceptibles. These processes are omitted in this version of the model given our purpose to analyse data reported over a 1-year period when the frequency of reinfection has been relatively low in our study setting [Hall *et al*. 2021] and population renewal considered negligible. Supplementary Material (Section S1) we formulate an extended model with reinfection and conduct some sensitivity analyses.

### 2.2. Individual variation in exposure to infection

In a directly transmitted infectious disease, such as COVID-19, variation in exposure to infection is primarily governed by patterns of connectivity among individuals. We incorporate this in system (1)-(4) assuming that individuals mix at random [Pastor-Satorras and Vespignani 2001, Miller *et al*. 2012]. In a related study we developed an assortative mixing version of the model [Aguas *et al*. 2020]. With random mixing and heterogeneous connectivity, the force of infection is written

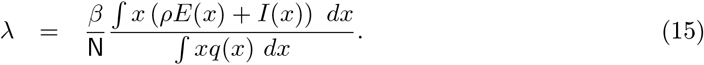

The basic reproduction number is

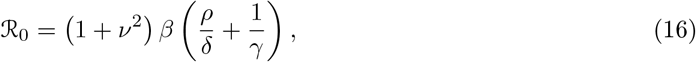

and ℛ_c_(*t*) = *c*(*t*) *·* ℛ_0_ as above.

Model (1)-(4) with variable exposure force of infection (15) can also be reduced to a finite system of ODEs when individual connectivity is gamma distributed, although, in contrast with variable susceptibility, the derivation is approximate in this case [Montalbán *et al*. 2020]:

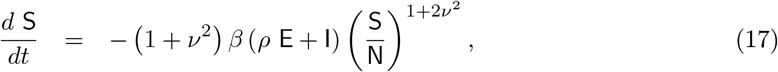

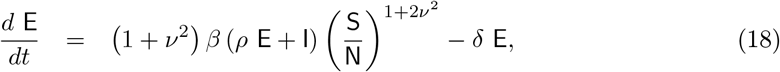

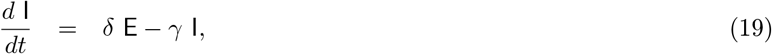

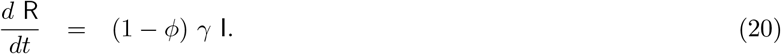

The effective reproduction number is

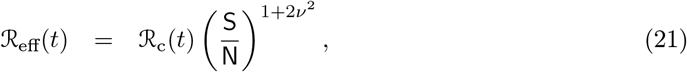

and the natural HIT

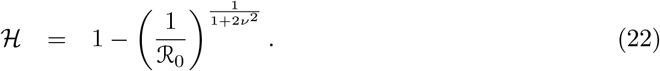

Notice that formulas (14) and (22) refer to herd immunity by natural infection and do not apply to herd immunity induced by vaccines (unless vaccination programmes were designed to mimic the selection exerted by natural infection which is not generally the case [Fine *et al*. 2011]). For a random vaccination programme, the HIT induced by the vaccine would be ℋ_*V*_ = [1*/*(1 − *σ*_*V*_)] *·* (1 − 1*/*ℛ_0_), where *σ*_*V*_ is the susceptibility of a vaccinated individual relative to unvaccinated. As a corollary, the HIT can only be reached by random vaccination if the vaccines in use are more than 1 − 1*/*ℛ_0_ efficacious against infection. For example, achieving the random vaccination HIT requires a vaccine that is at least 67% efficacious if ℛ_0_ = 3, and 80% efficacious if ℛ_0_ = 5, assuming 100% coverage. With lower coverage the demand on vaccine efficacy increases. This paper addresses primarily the natural HIT (ℋ).

## 3. Data

We used publicly available epidemiological data from the UK coronavirus dashboard describing the unfolding of the SARS-CoV-2 epidemic [https://coronavirus.data.gov.uk/] to estimate relevant transmission parameters for the larger nations: England (56 million population) and Scotland (5.5 million). Spefically, we extracted datasets containing daily deaths (deaths within 28 days of positive test by date of death), 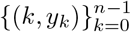, where *k* = 0 is the day when the cumulative rolling average of death numbers exceeded 5 *·* 10^−8^ of the population in both nations (10 March 2020). We also extracted from the same source data on the population considered fully vaccinated (two doses) at the time of this study.

Model outputs would then be fitted to the raw series of daily deaths between 10 March 2020 and 1 February 2021 (329 days in total) or 1 July 2020 (114 days) adopting an IFR of *ϕ* = 0.9% [Ward *et al*. 2021, Chen *et al*. 2021] throughout the fitted period. [Birrel *et al*. 2021] estimate that the IFR fluctuated between about 0.3% and 1.3% prior to vaccination, with a tendency to be low when infection spreads generally in the community and high when spread is predominantly in care homes and hospitals [McKeigue *et al*. 2021]. We assumed constant IFR but conducted analyses of sensitivity to its value (see Supplementary Material (Section S2) for results on *ϕ* = 0.7% and *ϕ* = 1.1%).

Initial conditions were set by solving the linear system of ODEs obtained by neglecting depletion of susceptibles in (9)-(12) (i.e., *d*E*/dt* = *β*(*ρ*E+I) −*δ*E, *d*I*/dt* = *δ*E−*γ*I, *d*R*/dt* = (1 − *ϕ*)*γ*I) for an early *R*_0_ = 2 [Li *et al*. 2020, Kissler *et al*. 2020], to find approximate relationships between variables (conditioning this on higher early transmission potential, such as *R*_0_ = 3, does not significantly change the results but leads to worse fits due to mismatch with early death counts). These approximations concerning the very early days of the epidemic enabled us to constrain the initial conditions as I(−*η*) = *y*_0_*/*0.002, E(−*η*) = I(−*η*)*/*0.136 and S(−*η*) = N − E(−*η*) −I(− *η*), where *η* is the excess duration of a fatal infection relative to non-fatal, and *y*_0_ is the number of deaths in the first day of the study. These numerical values are in the order of I(−*η*) ≈ *y*_0_*/ϕ*[1 −exp(−*γ*)] and E(−*η*) ≈ I(−*η*) */* [1 −exp(−*δ*)], which is expected early in the pandemic given the rates of progression from I to death and from E to I, respectively.

## 4. Model fitting and parameter estimation

In order to preserve identifiability, we made six main simplifying assumptions: (i) reinfection is negligible (see Section S1 for sensitivity analysis to reinfection); (ii) the IFR is constant throughout the fitted period (see Section S2 for sensitivity analysis to IFR) but lower due to vaccination in discussion of projections; (iii) vaccine effects are negligible during the fitted period, which in Section 5.1 ends 1 February 2021 (less than 1% fully vaccinated in the UK) and in Section 5.2 ends 1 July 2020 (no vaccines in use), but represented in projections beyond this phase as appropriate (see also Section 5.6); (iv) natural (seasonality, viral evolution and adaptive behavioural change) and interventional (NPI) modulators of the reproduction number are encapsulated in a single time varying parameter *c*(*t*) as illustrated in Fig. 2; (v) second and third lockdowns impact *c*(*t*) by the same factor; (vi) excess transmission from critically ill stages is negligible.

Parameter estimation was performed with software MATLAB (MathWorks, Natick, MA) using the PESTO (Parameter EStimation TOolbox) package [Stapor *et al*. 2018]. We assumed that numbers of SARS-CoV-2 infections are Poisson distributed.

We tried to reproduce the dynamics of COVID-19 deaths by estimating the set of parameters *θ* that maximises the log-likelihood (LL) of observing the daily numbers of reported deaths *Y* :

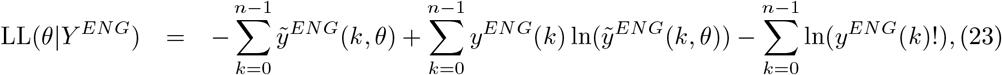

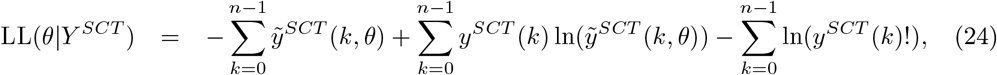

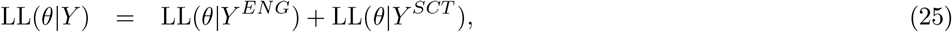

in which 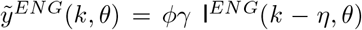 and 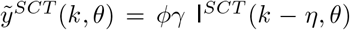 are the model output numbers of COVID-19 deaths at day *k* in England and Scotland for the set of parameters *θ*, 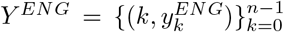 and 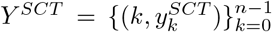 are the numbers of daily reported deaths, and *n* is the total number of days included in the analysis.

The set of parameters to be estimated is

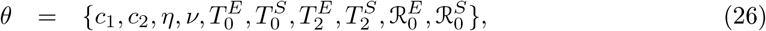

when fitting longer time series (Section 5.1) and

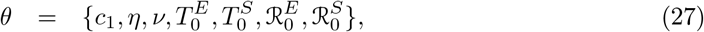

in fittings to shorter versions (Section 5.2).

To ensure that the estimated maximum is a global maximum, we performed 50 multi-start optimisations with initialisation parameters sampled from a Latin-Hypercube. The combination of parameters resulting in the maximal LL were used as a starting point for 100, 000 Markov chain Monte-Carlo (MCMC) iterations. From the resulting posterior distributions, we extracted the median estimates for each parameter and the respective 95% credible intervals. We used uniformly distributed priors with wide ranges.

We applied the outlined fitting procedure using both heterogeneity models (specifically individual variation in susceptibility and individual variation in exposure to infection) as well as a homogeneity model (obtained by setting the coefficient of variation to zero (*ν* = 0) in either model). The Akaike information criterion (AIC) was applied to select best fitting models.

## 5. Results

### 5.1. Estimated parameters and herd immunity thresholds

Variable susceptibility, variable connectivity and homogeneous models were fitted to series of COVID-19 deaths reported in England and Scotland until 1 February 2021. The fits are shown in Figs. 3, 4 and 5 (black solid curves; fitted data points in green), and the estimated parameters in Table 1. Maximum LL are also displayed, as well as AIC scores for model selection. We conclude that variable susceptibility and variable connectivity models are better supported by the data than the homogeneous model (lowest AIC in bold), as found in related studies [Aguas *et al*. 2020, Colombo *et al*. 2020]. Variable connectivity is slightly better supported in this case, although the reality most likely combines the two forms of heterogeneity.

**Figure 3:**
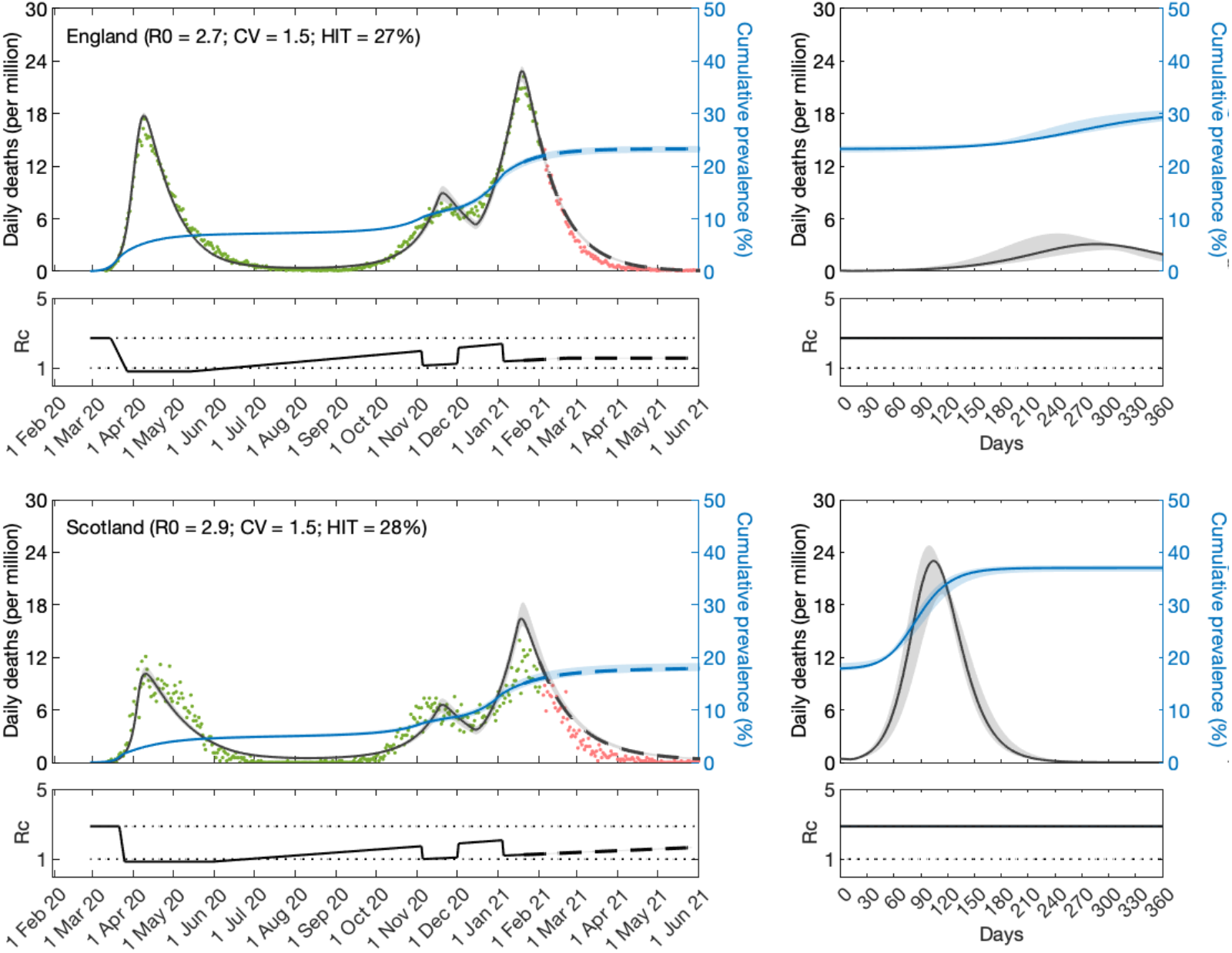
SARS-CoV-2 transmission in England and Scotland with individual variation in susceptibility to infection. Susceptibility factors implemented as gamma distributions (reduced model (9)-(12)). Modelled trajectories of COVID-19 deaths (black) and cumulative percentage infected (blue). Dots are data for daily reported deaths: fitted (green); out-of-sample (red). Basic reproduction numbers under control (ℛ_c_) displayed in shallow panels underneath the main plots. Left panels represent fitted segments as solid curves and projected scenarios as dashed. Right panels prolong those projections further in time assuming ℛ_c_(*t*) = ℛ_0_. Input parameters: progression from E to I (*δ* = 1*/*5.5 per day); recovery (*γ* = 1*/*4 per day); relative infectiousness between E and I stages (*ρ* = 0.5); and IFR (*ϕ* = 0.9%). Initial basic reproduction numbers, coefficients of variation and control parameters estimated by Bayesian inference (estimates in Table 1). Fitted curves represent best fitting trajectories and shades are 95% credible intervals generated from 10, 000 posterior samples.

**Figure 4:**
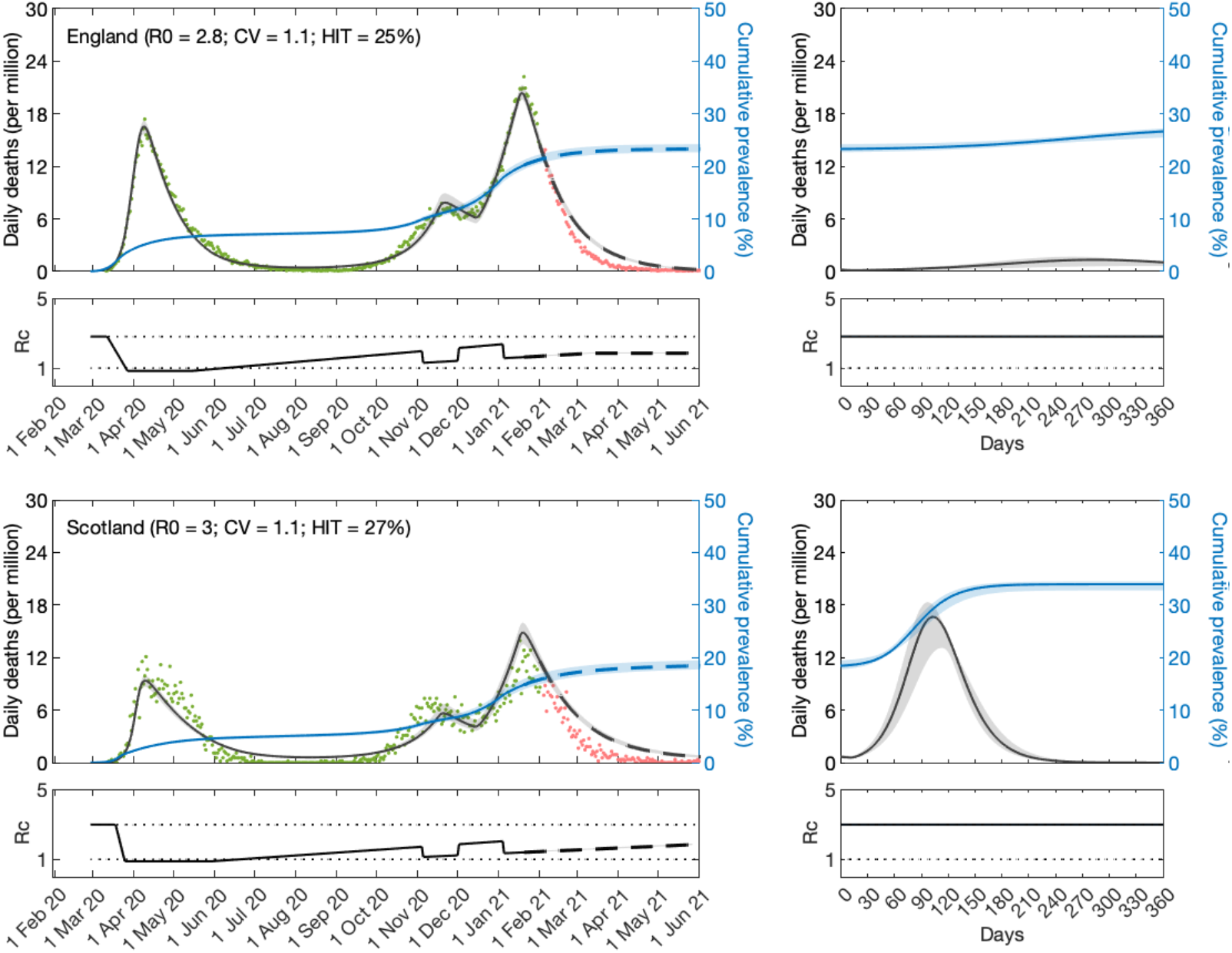
SARS-CoV-2 transmission in England and Scotland with individual variation in exposure to infection. Connectivity factors implemented as gamma distributions (reduced model (17)-(20)). Modelled trajectories of COVID-19 deaths (black) and cumulative percentage infected (blue). Dots are data for daily reported deaths: fitted (green); out-of-sample (red). Basic reproduction numbers under control (ℛ_c_) displayed in shallow panels underneath the main plots. Left panels represent fitted segments as solid curves and projected scenarios as dashed. Right panels prolong those projections further in time assuming ℛ_c_(*t*) = ℛ_0_. Input parameters: progression from E to I (*δ* = 1*/*5.5 per day); recovery (*γ* = 1*/*4 per day); relative infectiousness between E and I stages (*ρ* = 0.5); and IFR (*ϕ* = 0.9%). Initial basic reproduction numbers, coefficients of variation and control parameters estimated by Bayesian inference (estimates in Table 1). Fitted curves represent best fitting trajectories and shades are 95% credible intervals generated from 10, 000 posterior samples.

**Figure 5:**
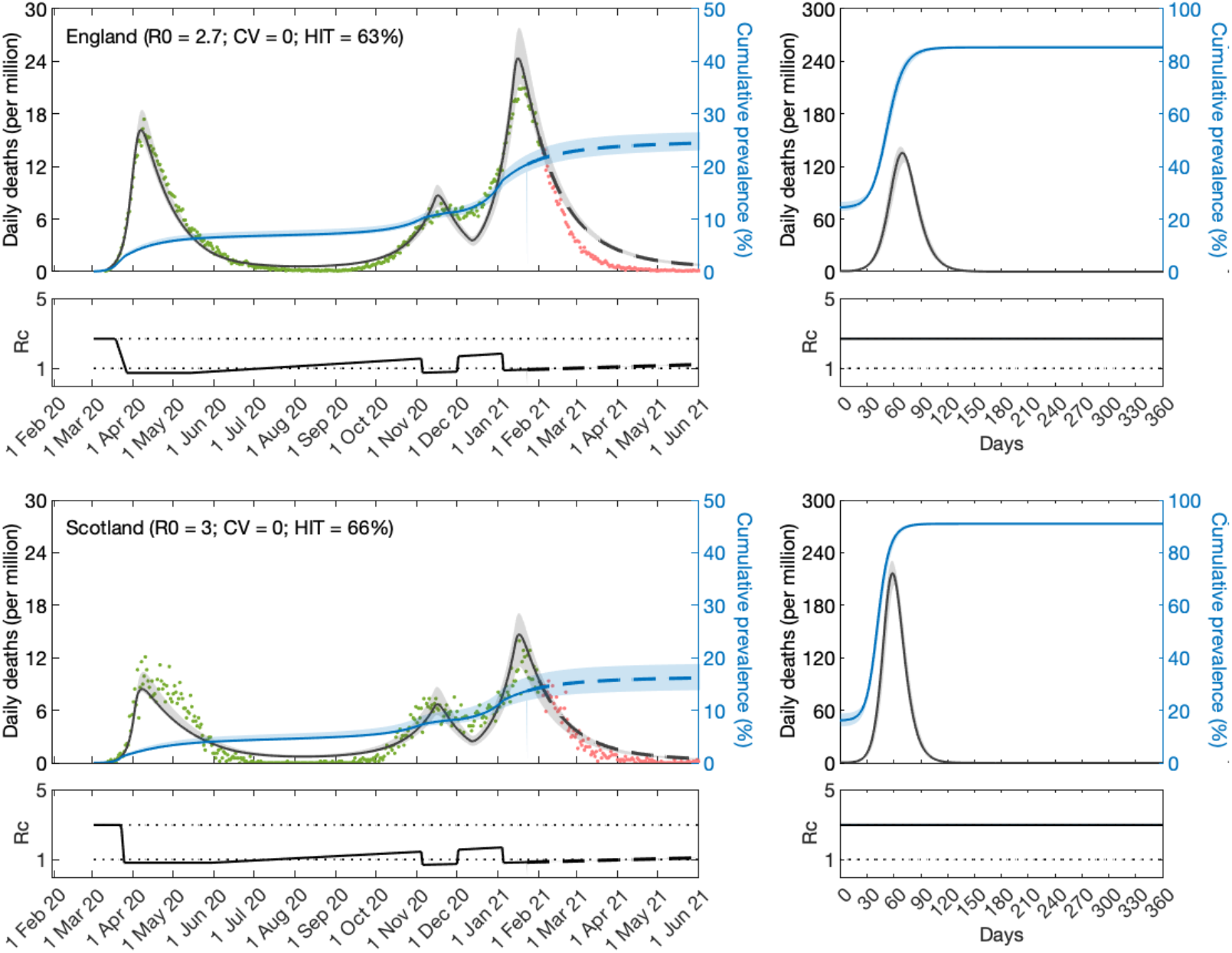
SARS-CoV-2 transmission in England and Scotland assuming homogeneity. Reduced model (9)-(12) or (17)-(20) with *ν* = 0. Modelled trajectories of COVID-19 deaths (black) and cumulative percentage infected (blue). Dots are data for daily reported deaths: fitted (green); out-of-sample (red). Basic reproduction numbers under control (ℛ_c_) displayed in shallow panels underneath the main plots. Left panels represent fitted segments as solid curves and projected scenarios as dashed. Right panels prolong those projections further in time assuming ℛ_c_(*t*) = ℛ_0_. Input parameters: progression from E to I (*δ* = 1*/*5.5 per day); recovery (*γ* = 1*/*4 per day); relative infectiousness between E and I stages (*ρ* = 0.5); and IFR (*ϕ* = 0.9%). Initial basic reproduction numbers and control parameters estimated by Bayesian inference (estimates in Table 1). Fitted curves represent best fitting trajectories and shades are 95% credible intervals generated from 10, 000 posterior samples.

**Table 1:** Model parameters estimated by Bayesian inference based on daily deaths until 1 February 2021 (with reduced models). Model selection based on maximum log-likelihood (LL) and Akaike information criterion (AIC). Best fitting models have lower AIC scores (bold). Infection fatality ratio, *ϕ* = 0.9%. Herd immunity threshold (ℋ) calculated from ℛ_0_ and CV using formulas (14) or (22), as appropriate. *T*_0_ and *T*_2_ parameterise linear reduction and increase in transmissibility, respectively, before and after first lockdown (larger *T* ⇔ lower slope; Fig. 2).

|  | Heterogeneous susceptibility |  | Heterogeneous connectivity |  | Homogeneous |  |
| --- | --- | --- | --- | --- | --- | --- |
|  | Median | 95% CI | Median | 95% CI | Median | 95% CI |
| *Common parameters* |  |  |  |  |  |  |
| $c_1^a$ | 0.2968 | (0.2958, 0.2988) | 0.2993 | (0.2976, 0.3015) | 0.2751 | (0.2740, 0.2772) |
| $c_2^b$ | 0.5761 | (0.5731, 0.5843) | 0.6601 | (0.6535, 0.6661) | 0.4803 | (0.4776, 0.4869) |
| $\eta^c$ | 12 | {12} | 12 | {12} | 9 | {9, 10} |
| $\nu^d$ | 1.475 | (1.470, 1.487) | 1.120 | (1.111, 1.145) | 0 | — |
| *England* |  |  |  |  |  |  |
| $T_0$ | 3.616 | (3.480, 3.759) | 0.6549 | (0.3925, 0.8821) | 7.595 | (6.955, 7.771) |
| $T_2$ | 283.5 | (282.4, 286.0) | 302.2 | (295.8, 303.9) | 416.1 | (408.9, 418.3) |
| $\mathcal{R}_0$ | 2.709 | (2.701, 2.716) | 2.807 | (2.781, 2.827) | 2.691 | (2.683, 2.701) |
| $\mathcal{H}$ | 26.94% | (26.62%, 27.10%) | 25.48% | (24.61%, 25.89%) | 62.84% | (62.72%, 62.98%) |
| *Scotland* |  |  |  |  |  |  |
| $T_0$ | 10.10 | (9.854, 10.72) | 6.934 | (6.460, 7.508) | 11.78 | (11.36, 12.58) |
| $T_2$ | 360.8 | (359.0, 364.4) | 399.9 | (390.2, 406.3) | 540.0 | (524.9, 546.9) |
| $\mathcal{R}_0$ | 2.874 | (2.856, 2.912) | 2.981 | (2.948, 3.053) | 2.978 | (2.954, 2.995) |
| $\mathcal{H}$ | 28.29% | (27.88%, 28.69%) | 26.75% | (25.81%, 27.52%) | 66.41% | (66.14%, 66.61%) |
| *Model selection* |  |  |  |  |  |  |
| LL | −3707 |  | −3684 |  | −5893 |  |
| AIC | 7434 |  | **7387** |  | 11805 |  |

<sup>a</sup>transmissibility reduction due to lockdown 1,
<sup>b</sup>transmissibility reduction due to lockdowns 2 and 3,
<sup>c</sup>difference between mean-time-to-death and mean-time-to-recovery (sampled from a continuous interval and reduced to the nearest integer before entering the model),
<sup>d</sup>coefficient of variation (CV).

Note that while we report CV in all fits, our method is not purposed to estimate levels of heterogeneity solely in susceptibility or connectivity as a goal. Because the two forms of heterogeneity have similar effects on epidemic trajectories, separating relative contributions would pose an identifiability problem that our approach cannot address. Each heterogeneity model we consider assumes that there is no heterogeneity of the other kind. From fitting these two extremes, however, we conclude that they are equally able to fit data and have similar consequences for metrics of interest and projections. Hence a decomposition is not necessary for our purposes.

Given the estimated values for ℛ_0_ and CV (*ν*) we derive the natural HIT through formulas (14) or (22) as appropriate, obtaining ℋ in the range 25 − 27% in England, and 26 − 29% in Scotland. With the homogeneous model, the inferences would be considerably higher, ℋ = 63% in England and in the range 66 − 67% in Scotland, as in early expectations [Kwok *et al*. 2020].

We then prolong model trajectories (dashed curves) for another 4 months (until 1 June 2021) to begin comparisons with data beyond fitted period (red). All models project more deaths than observed, as expected given that UK initiated a mass vaccination programme in late 2020 which would have started impacting the epidemic by February 2021. With reduced models adopted here, implementation of vaccination is not straightforward but in Supplementary Material we replicate the fits with explicit gamma distributions (Section S1) and simulate a crude approximation to the UK vaccination programme (Figs. S1, S2 and S3). In Section 5.6, we focus on England and implement vaccination in more detail according to data on fully vaccinated people in the nation. A relevant question at the time of these results was whether achieved population immunity had been enough to prevent an exit wave as contact restrictions were lifted from 8 March 2021 onwards culminating with the removal of most restrictions from 19 July 2021 (especially in England where this was dubbed “Freedom Day”). To assess how each of the models might answer this question we plotted the cumulative number of estimated infections (blue) to determine the percentage of the population infected by June 2021. We find that both nations remain below the natural herd immunity threshold ℋ (Scotland more than England). Hence, without vaccination, an exit wave might have been expected. To visualise its magnitude we include separate panels on the right where the model is run for approximately 12 months, using as initial conditions the end conditions from left panels and ℛ_c_(*t*) = ℛ_0_. Here this is done crudely but in Section S1 we explore a range of vaccination scenarios and in Section 5.6 we refine the projections in the case of England (the same could be done for Scotland). It is striking that heterogeneous models project exit waves one order of magnitude lower (or less) than when homogeneity is assumed, a feature that we explore more thoroughly in section 5.6.

In Supplementary Material we also show that these results are robust to including reinfection (with a risk of 0.1 relative to the average risk of first infection [Hall *et al*. 2021]) (Section S2), arriving at similar HITs, and changing IFR (to 0.7% or 1.0%) (Section S3). This agrees with the expectation that assuming a lower IFR results in higher HIT, and a higher IFR results in lower HIT. Less intuitive is that when we fit the models with a different IFR, all parameters readjust and exit waves appear relatively conserved (perhaps slightly slower with lower IFR (higher HIT) and higher with higher IFR (lower HIT), illustrating the fragility of taking HITs out of context). Similar conserveness is noticed when reinfection is included. Model selection by AIC continues to favour heterogeneous models by large in all scenarios.

### 5.2. Parameter estimation early in the pandemic

In a pandemic it is important to estimate model parameters early when data series are short. To test suitability of our methods for such task, we apply fitting procedures to series of COVID-19 deaths in England and Scotland until 1 July 2020, as Europe was recovering from the first wave. As an initial exploration we perform the fittings as in Section 5.1, estimating all parameters in (26) except *c*_2_ which had no effect prior to November 2020. The results, displayed in Supplementary Material (Section S4), provide similar estimates to those obtained with longer series in Section 5.1 except for parameter *T*_2_ that determines the slope of contact reactivation following lifting of first lockdown. This preliminary analysis suggests *T*_2_ to be very high (or equivalently, slope of contact reactivation close to zero), presumably due to absence of detectable effects of lifting restrictions in such short timeframe. Technically, the estimation of *T*_2_ becomes unsuitable in this framework since as *T*_2_ → ∞ so does uncertainty about its value. One way to circumvent this problem could be to reparameterise contact reactivation in terms of slope rather than time to reach baseline *c*(*t*) = 1. Meanwhile, here we present two scenarios by constraining *T*_2_ (Fig. 6).

**Figure 6:**
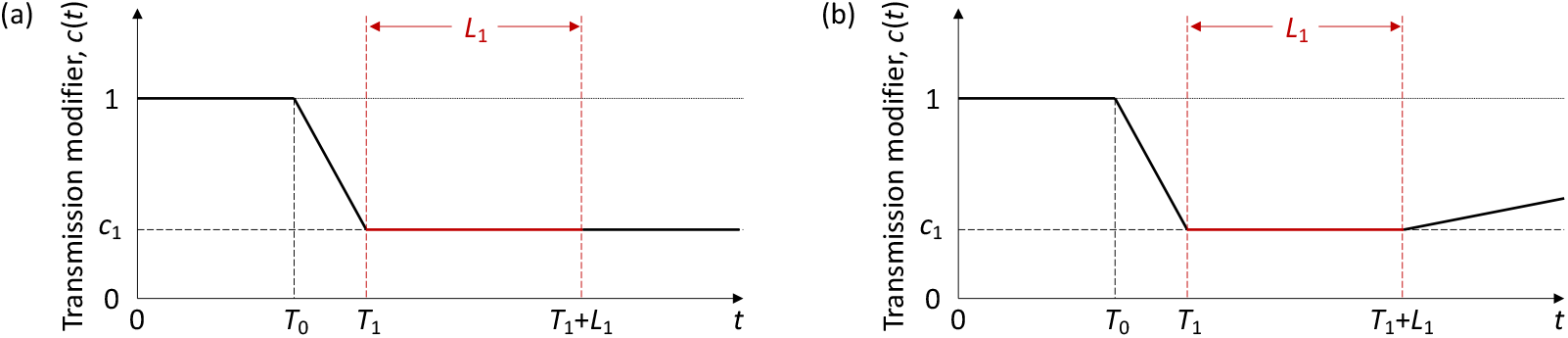
Schematic illustration of the factor *c*(*t*), representing the effect of NPIs and adaptive behavioural changes on transmission. *L*_1_ represents the known duration (and timing) of the first lockdown as imposed by governments. *T*_0_ is the number of days in the series prior to the gradual contact reductions early in the pandemic (estimated). *T*_1_ (*> T*_0_) is the day the first lockdown begins (informed by data). *T*_2_ is the number of days for the ramp of increasing transmission after first lockdown to return to baseline *c*(*t*) = 1 and determines the slope that distinguishes the two panels: (a) *T*_2_ → ∞; (b) *T*_2_ = 120 days.

Informed by preliminary analysis in Section S4, we first run a scenario where *c*(*t*) remains strictly horizontal until end of fitting period (*T*_2_ → ∞, Fig. 7). This assumption appears to align with stringency index, a composite measure based on government response indicators (school closures, workplace closures, travel bans, etc) developed by Oxford Covid-19 Government Response Tracker (OxCGRT) [Hale *et al*. 2020] (blue in ℛ_c_ panels, scaled to match lower and higher bounds in our stylised profiles). Support for heterogeneous models is again very significant according to AIC scores, and optimal parameter values result in similar HITs to when *T*_2_ was estimated: ℋ = 34% (95% CI, 25 − 57%) for heterogeneous models; and ℋ = 70% (95% CI, 68 − 71%) for homogeneous. Compare Tables 2 (*T*_2_ *→ ∞*) and S5 (*T*_2_ estimated). Second, we run a scenario with finite *T*_2_ to inform how estimated parameters might respond to assumptions about contact reactivation. Assuming *T*_2_ = 120 days (crudely motivated by mobility data [Google 2020], blue dots in Fig. 8), we find that support for heterogeneous models persists but estimated coefficients of variation become larger, reflecting in lower HITs (ℋ = 15% (95% CI, 12 − 17%), Table 3). In absence of further information we might have expected true parameter values to be somewhere between these two scenarios. This is supported by results on longer COVID-19 series in Section 5.1 (ℋ in the range 25 − 29%, Table 1), which is inside, but towards the low end, of the range motivated by stringency index.

**Figure 7:**
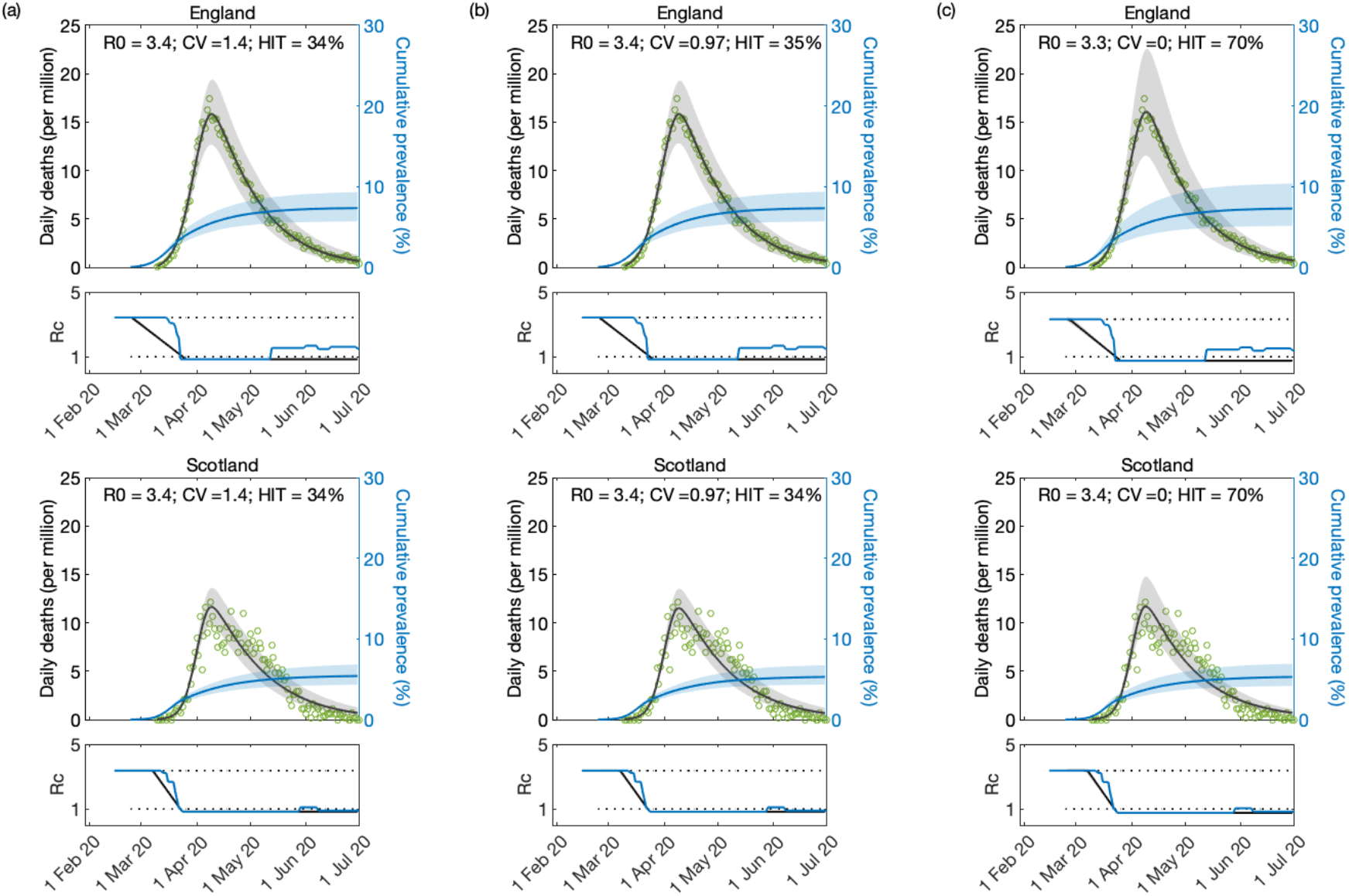
Model fitting to first wave of the SARS-CoV-2 pandemic assuming *T*_2_ → ∞. Modelled trajectories of COVID-19 deaths (black) and cumulative percentage infected (blue). Green dots are data for daily reported deaths. Basic reproduction numbers under control (ℛ_c_) displayed in shallow panels underneath the main plots. Government stringency indices [Hale *et al*. 2020] traced in blue. Input parameters: progression from E to I (*δ* = 1*/*5.5 per day); recovery (*γ* = 1*/*4 per day); relative infectiousness between E and I stages (*ρ* = 0.5); and IFR (*ϕ* = 0.9%). Initial basic reproduction numbers, coefficients of variation and control parameters estimated by Bayesian inference (estimates in Table 2). Fitted curves represent best fitting trajectories and shades are 95% credible intervals generated from 10, 000 posterior samples. (a) Individual variation in susceptibility to infection (model (9)-(12)). (b) Individual variation in exposure to infection ((17)-(20)). (c) Homogeneous susceptibility and exposure (either model with *ν* = 0).

**Table 2:**
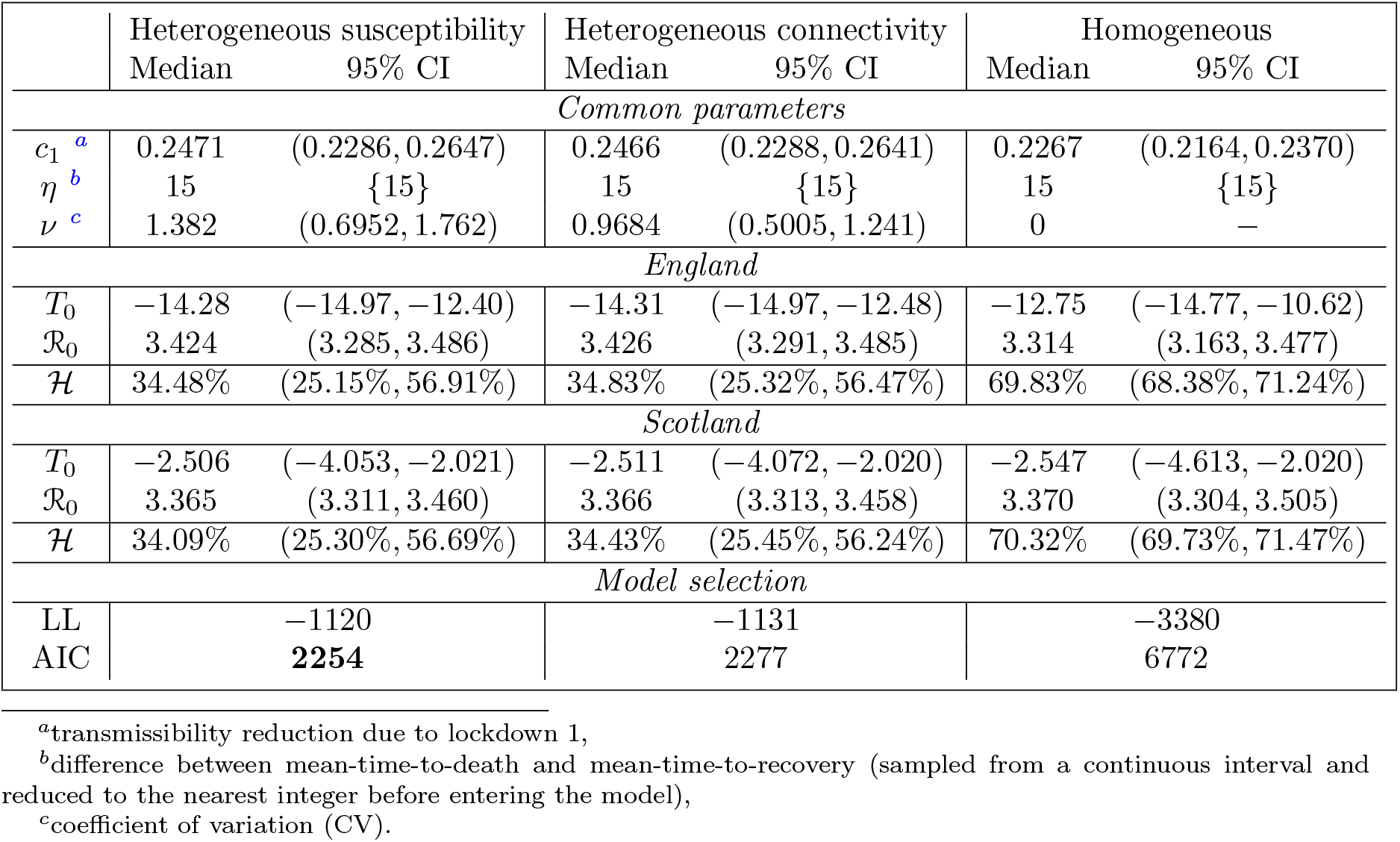
Model parameters estimated by Bayesian inference based on daily deaths until 1 July 2020, assuming constant ℛ_c_(*t*) from first lockdown onwards (i.e., *T*_2_ → ∞). Model selection based on maximum log-likelihood (LL) and Akaike information criterion (AIC). Best fitting models have lower AIC scores (bold). Infection fatality ratio, *ϕ* = 0.9%. Herd immunity threshold (ℋ) calculated from ℛ_0_ and CV using formulas (14) or (22), as appropriate. *T*_0_ and *T*_2_ parameterise linear reduction and increase in transmissibility, respectively, before and after first lockdown (larger *T* ⇔ lower slope; Fig. 6).

|  | Heterogeneous susceptibility |  | Heterogeneous connectivity |  | Homogeneous |  |
| --- | --- | --- | --- | --- | --- | --- |
|  | Median | 95% CI | Median | 95% CI | Median | 95% CI |
| *Common parameters* |  |  |  |  |  |  |
| $c_1^a$ | 0.2471 | (0.2286, 0.2647) | 0.2466 | (0.2288, 0.2641) | 0.2267 | (0.2164, 0.2370) |
| $\eta^b$ | 15 | {15} | 15 | {15} | 15 | {15} |
| $\nu^c$ | 1.382 | (0.6952, 1.762) | 0.9684 | (0.5005, 1.241) | 0 | — |
| *England* |  |  |  |  |  |  |
| $T_0$ | -14.28 | (-14.97, -12.40) | -14.31 | (-14.97, -12.48) | -12.75 | (-14.77, -10.62) |
| $\mathcal{R}_0$ | 3.424 | (3.285, 3.486) | 3.426 | (3.291, 3.485) | 3.314 | (3.163, 3.477) |
| $\mathcal{H}$ | 34.48% | (25.15%, 56.91%) | 34.83% | (25.32%, 56.47%) | 69.83% | (68.38%, 71.24%) |
| *Scotland* |  |  |  |  |  |  |
| $T_0$ | -2.506 | (-4.053, -2.021) | -2.511 | (-4.072, -2.020) | -2.547 | (-4.613, -2.020) |
| $\mathcal{R}_0$ | 3.365 | (3.311, 3.460) | 3.366 | (3.313, 3.458) | 3.370 | (3.304, 3.505) |
| $\mathcal{H}$ | 34.09% | (25.30%, 56.69%) | 34.43% | (25.45%, 56.24%) | 70.32% | (69.73%, 71.47%) |
| *Model selection* |  |  |  |  |  |  |
| LL | -1120 |  | -1131 |  | -3380 |  |
| AIC | **2254** |  | 2277 |  | 6772 |  |

<sup>a</sup>transmissibility reduction due to lockdown 1,
<sup>b</sup>difference between mean-time-to-death and mean-time-to-recovery (sampled from a continuous interval and reduced to the nearest integer before entering the model),
<sup>c</sup>coefficient of variation (CV).

**Figure 8:**
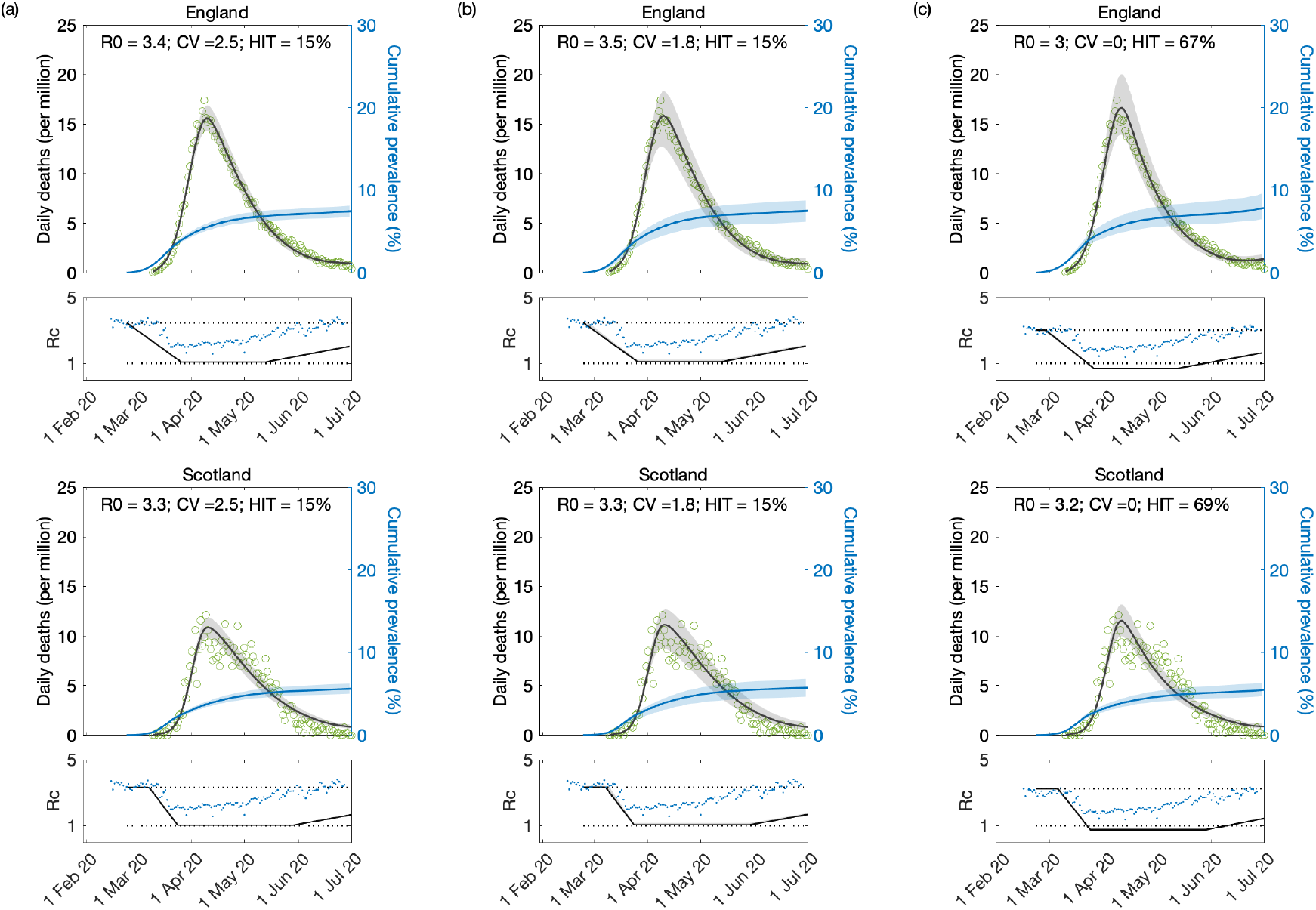
Model fitting to first wave of the SARS-CoV-2 pandemic assuming *T*_2_ = 120 days. Modelled trajectories of COVID-19 deaths (black) and cumulative percentage infected (blue). Green dots are data for daily reported deaths. Basic reproduction numbers under control (ℛ_c_) displayed in shallow panels underneath the main plots. Blue dots represent UK Google mobility index [Google 2020]. Input parameters: progression from E to I (*δ* = 1*/*5.5 per day); recovery (*γ* = 1*/*4 per day); relative infectiousness between E and I stages (*ρ* = 0.5); and IFR (*ϕ* = 0.9%). Initial basic reproduction numbers, coefficients of variation and control parameters estimated by Bayesian inference (estimates in Table 3). Fitted curves represent best fitting trajectories and shades are 95% credible intervals generated from 10, 000 posterior samples. (a) Individual variation in susceptibility to infection (model (9)-(12)). (b) Individual variation in exposure to infection ((17)-(20)). (c) Homogeneous susceptibility and exposure (either model with *ν* = 0).

**Table 3:** Model parameters estimated by Bayesian inference based on daily deaths until 1 July 2020, assuming that after first lockdown ℛ_c_(*t*) begins a gradual return to the baseline ℛ_0_ at a fixed rate (corresponding to *T*_2_ = 120 days in this case). Model selection based on maximum log-likelihood (LL) and Akaike information criterion (AIC). Best fitting models have lower AIC scores (bold). Infection fatality ratio, *ϕ* = 0.9%. Herd immunity threshold (ℋ) calculated from ℛ_0_ and CV using formulas (14) or (22), as appropriate. *T*_0_ and *T*_2_ parameterise linear reduction and increase in transmissibility, respectively, before and after first lockdown (larger *T* ⇔ lower slope; Fig. 6).

|  | Heterogeneous susceptibility |  | Heterogeneous connectivity |  | Homogeneous |  |
| --- | --- | --- | --- | --- | --- | --- |
|  | Median | 95% CI | Median | 95% CI | Median | 95% CI |
| *Common parameters* |  |  |  |  |  |  |
| $c_1^a$ | 0.3133 | (0.2989, 0.3277) | 0.3182 | (0.3002, 0.3419) | 0.2358 | (0.2291, 0.2419) |
| $\eta^b$ | 15 | {15} | 15 | {13, 14, 15} | 17 | {17} |
| $\nu^c$ | 2.521 | (2.369, 2.653) | 1.822 | (1.681, 2.011) | 0 | — |
| *England* |  |  |  |  |  |  |
| $T_0$ | -14.79 | (-14.991, -13.922) | -14.815 | (-14.994, -13.952) | -11.51 | (-13.00, -10.17) |
| $\mathcal{R}_0$ | 3.438 | (3.380, 3.470) | 3.451 | (3.383, 3.533) | 3.017 | (2.940, 3.109) |
| $\mathcal{H}$ | 15.46% | (14.06%, 17.15%) | 14.97% | (12.55%, 17.29%) | 66.85% | (65.99%, 67.84%) |
| *Scotland* |  |  |  |  |  |  |
| $T_0$ | -2.253 | (-3.033, -2.010) | -2.229 | (-2.930, -2.007) | -4.407 | (-5.651, -4.017) |
| $\mathcal{R}_0$ | 3.308 | (3.263, 3.358) | 3.319 | (3.268, 3.388) | 3.234 | (3.194, 3.302) |
| $\mathcal{H}$ | 15.01% | (13.68%, 16.74%) | 14.53% | (12.22%, 16.77%) | 69.08% | (68.69%, 69.71%) |
| *Model selection* |  |  |  |  |  |  |
| LL | -951.7 |  | -960.5 |  | -3308 |  |
| AIC | **1917** |  | 1935 |  | 6627 |  |

<sup>a</sup>transmissibility reduction due to lockdown 1,
<sup>b</sup>difference between mean-time-to-death and mean-time-to-recovery (sampled from a continuous interval and reduced to the nearest integer before entering the model),
<sup>c</sup>coefficient of variation (CV).

### 5.3. Using government response indicators

In this section we use the stringency index, which tracks the strictness of government policies that restricted people’s behaviour during the COVID-19 pandemic [Hale *et al*. 2020], as a main factor capable to modify transmission over time. In Fig. 9 we show stringency indices for England (red) and Scotland (blue), both scaled to be 1 in February 2020 and to match an illustrative *c*_1_ = 0.2 during lockdown 1. We refer to this scaled time-dependent index as *u*(*t*).

**Figure 9:**
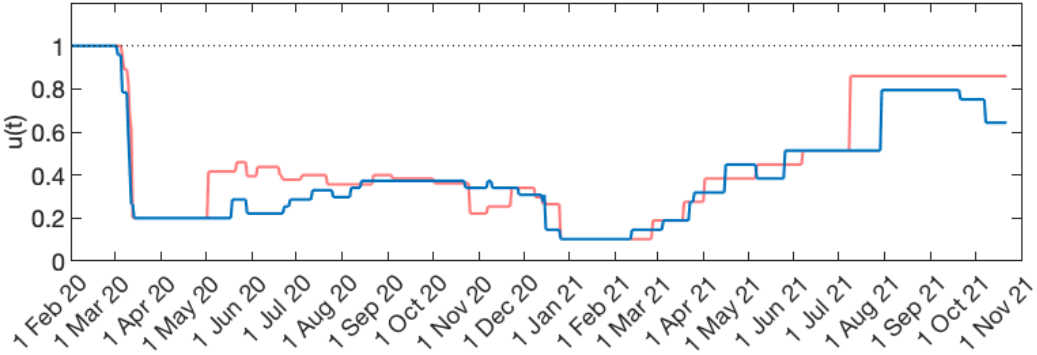
Government stringency index. To facilitate visual comparisons, raw indices [Hale *et al*. 2020] were scaled so they would both be 1 in February 2020 and 0.2 during lockdown 1. The resulting time-dependent *u*(*t*) are shown for England (blue) and Scotland (green).

Here we replicate model fittings to series of COVID-19 deaths until 1 February 2021 using *u*(*t*) to help determine the overall profile *c*(*t*). We assume that until the 1 July 2020 (end of series fitted in Section 5.2) the profile *c*(*t*) is entirely determined by *u*(*t*). From then on we include a multiplicative factor that increases linearly from 1 to reach some value *w* by the last day of the fitted series. Parameters *c*_1_ and *w* are estimated. In the case of Scotland this was not sufficient to allow acceptable fits. Looking at Fig. 9 we notice that the stringency index for Scotland shows no signal for the second lockdown which happened mostly throughout November 2020 (visible in the stringency index for England). Both death series, however, show a clear reversal in epidemic growth around that time. Presumably restrictions imposed by the government in England might have affected Scotland due to mobility across the border and behavioural influences. To enable fittings to the two nations we modify the Scottish stringency to resemble the English from the 1 July 2020 onwards. The results are provided in Fig. 10 and Table 4.

**Figure 10:**
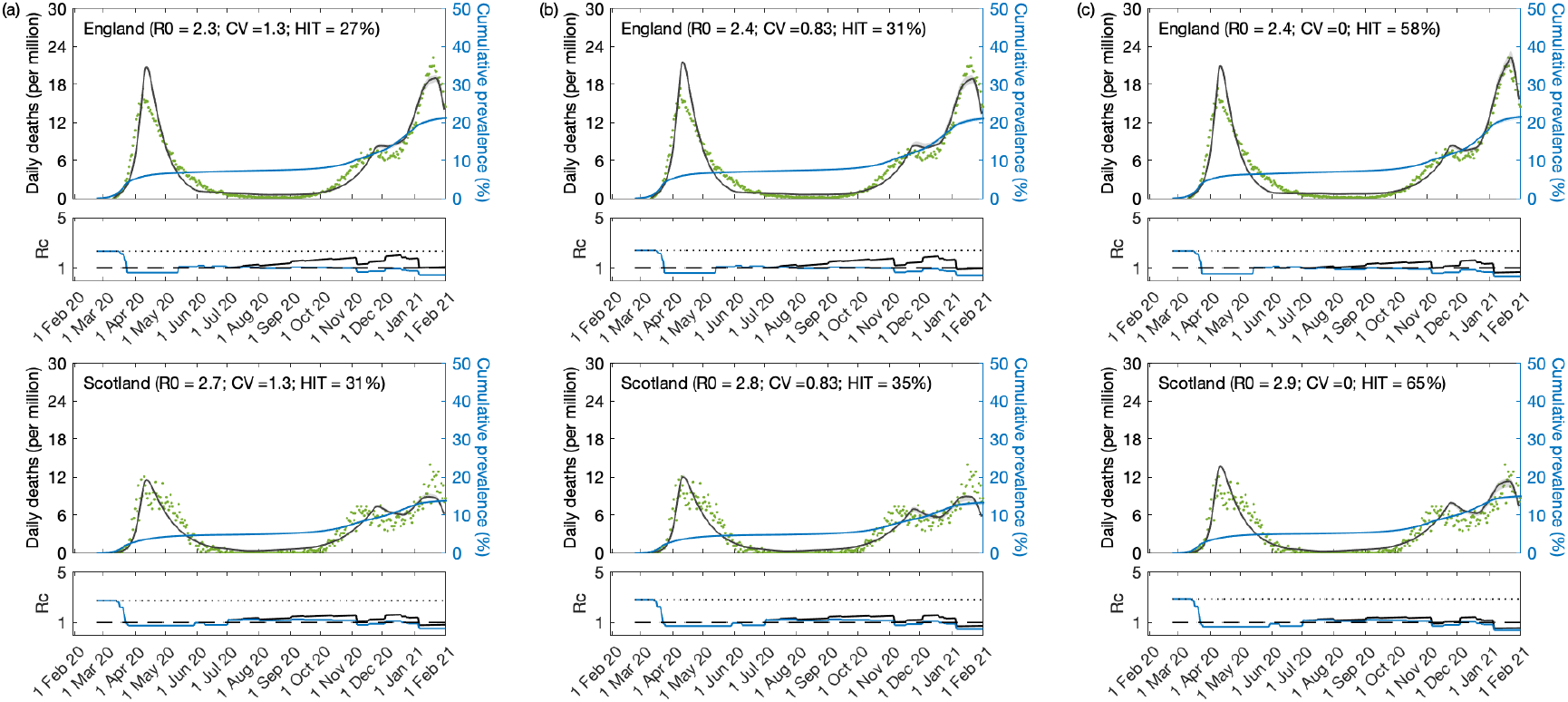
SARS-CoV-2 transmission constrained by government stringency index. (a) individual variation in susceptibility to infection; (b) individual variation in exposure to infection; (c) assuming homogeneity. Modelled trajectories of COVID-19 deaths (black) and cumulative percentage infected (blue). Green dots are data for daily reported deaths. Basic reproduction numbers modified by the stringency index [Hale *et al*. 2020] (ℛ_c_) displayed in shallow panels underneath the main plots. Input parameters: progression from E to I (*δ* = 1*/*5.5 per day); recovery (*γ* = 1*/*4 per day); relative infectiousness between E and I stages (*ρ* = 0.5); and IFR (*ϕ* = 0.9%). Initial basic reproduction numbers, coefficients of variation and control parameters estimated by Bayesian inference (estimates in Table 4). Fitted curves represent best fitting trajectories and shades are 95% credible intervals generated from 10, 000 posterior samples.

**Table 4:** Model parameters estimated by Bayesian inference with transmission constrained by a government stringency index [Hale *et al*. 2020]. Model selection based on maximum log-likelihood (LL) and Akaike information criterion (AIC). Best fitting models have lower AIC scores (bold). Infection fatality ratio, *ϕ* = 0.9%. Herd immunity threshold (ℋ) calculated from ℛ_0_ and CV using formulas (14) or (22), as appropriate. *w* accounts for increases in transmission since the end of lockdown 1 due to factors other than government response.

|  | Heterogeneous susceptibility |  | Heterogeneous connectivity |  | Homogeneous |  |
| --- | --- | --- | --- | --- | --- | --- |
|  | Median | 95% CI | Median | 95% CI | Median | 95% CI |
| *Common parameters* |  |  |  |  |  |  |
| $c_1$ <sup>a</sup> | 0.2808 | (0.2806, 0.2814) | 0.2626 | (0.2624, 0.2634) | 0.2345 | (0.2343, 0.2346) |
| $\eta$ <sup>b</sup> | 17 | {17} | 16 | {16} | 16 | {16} |
| $\nu$ <sup>c</sup> | 1.314 | (1.309, 1.320) | 0.8298 | (0.8272, 0.8447) | 0 | — |
| *England* |  |  |  |  |  |  |
| $w$ | 2.430 | (2.427, 2.434) | 2.339 | (2.338, 2.345) | 2.096 | (2.094, 2.096) |
| $\mathcal{R}_0$ | 2.336 | (2.334, 2.337) | 2.398 | (2.398, 2.400) | 2.356 | (2.354, 2.358) |
| $\mathcal{H}$ | 26.74% | (26.58%, 26.87%) | 30.78% | (30.25%, 30.90%) | 57.56% | (57.52%, 57.59%) |
| *Scotland* |  |  |  |  |  |  |
| $w$ | 1.575 | (1.571, 1.578) | 1.540 | (1.540, 1.542) | 1.430 | (1.429, 1.437) |
| $\mathcal{R}_0$ | 2.750 | (2.748, 2.753) | 2.821 | (2.820, 2.833) | 2.878 | (2.876, 2.879) |
| $\mathcal{H}$ | 31.00% | (30.83%, 31.15%) | 35.36% | (34.76%, 35.58%) | 65.26% | (65.23%, 65.27%) |
| *Model selection* |  |  |  |  |  |  |
| LL | −5801 |  | −5788 |  | −6220 |  |
| AIC | 11616 |  | **11589** |  | 12452 |  |

<sup>a</sup>transmissibility reduction due to lockdown 1,
<sup>b</sup>difference between mean-time-to-death and mean-time-to-recovery (sampled from a continuous interval and reduced to the nearest integer before entering the model),
<sup>c</sup>coefficient of variation (CV).

The quality of these fits is not as good as when we used a stylised transmission modifier *c*(*t*) in Section 5.1, presumably because it does not account as flexibility for behavioural factors other than those resulting from government restrictions. Nevertheless the results remain consistent also in this scenario. Using the stringency index as outlined in this section the heterogeneous models are again better supported by the data according to AIC, and we estimate ℋ in the range 27 − 31% in England and 31 − 36% in Scotland for heterogeneous models, while 58% is obtained for England and 65% for Scotland when homogeneity is assumed.

Overall this analysis suggests that in the eventuality of future pandemics our models, with more or less stylised mitigations, can be used consistently at differently stages as the epidemic unfolds. This study of England and Scotland demonstrates that although applications will always benefit from knowledge of time-dependent effects on transmission, considerable progress can be made by model-based inference before detailed measurements of such factors are available. It is also possible for sensibly designed stylised models to perform better than those constrained by incomplete information. Inferences should be subsequently tested, however, and assumptions improved as more data becomes available.

At this stage, our parameter inferences can be confronted with several types of empirical estimates. First, the basic reproduction numbers estimated here (in the order of ℛ_0_ = 3) align with those from numerous independent studies (e.g., [Flaxman *et al*. 2020, Hilton *et al*. 2020, Kwok *et al*. 2020]). Second, the transmission reduction due to first lockdown, 70 − 80% (i.e., *c*_1_ between 0.2 and 0.3) is in agreement with direct contact measurements [Jarvis *et al*. 2020]. Third, the estimated time from detectable infection to death (1*/γ* + *η*, between 10 and 20 days) agrees with [Verity *et al*. 2020, Wood 2021], more so when heterogeneity models are used. Forth, our time-dependent transmission profiles, *c*(*t*) (parameterised by *T*_0_ and *T*_2_), align well with the stringency index [Hale *et al*. 2020] and might align even better with some hybrid indicator combining government measures with voluntary adaptive behavioural changes (captured by mobility patterns tracked by Google). Finally, the coefficient of variation (*ν*), the main novelty of this study, is assessed in a dedicated section (Section 5.4).

### 5.4. Coefficients of variation from empirical studies

#### 5.4.1. Contact surveys

Contact patterns provide one of the easiest sources of heterogeneity to study directly. One approach is to use large-scale diary experiments to collect self-reported logs of close or physical contact from study participants. In this section we show data from several of these contact-pattern studies, listed in Table 5. In Fig. 11 we show gamma-fits for the contact distribution of each study on a log scale, along with the CV for each empirical distribution. In Supplementary Material we show that lognormal distributions provide considerable worse fits (Fig. S15).

**Table 5:** Contact pattern studies used in this review.

|  | Dataset | citation |
| --- | --- | --- |
| 1 | 2008_Mossong_BE | [Mossong *et al.* 2008] |
| 2 | 2008_Mossong_EU | [Mossong *et al.* 2008] |
| 3 | 2008_Mossong_IT | [Mossong *et al.* 2008] |
| 4 | 2008_Mossong_LU | [Mossong *et al.* 2008] |
| 5 | 2008_Mossong_NL | [Mossong *et al.* 2008] |
| 6 | 2008_Mossong_OT | [Mossong *et al.* 2008] |
| 7 | 2008_Mossong_PL | [Mossong *et al.* 2008] |
| 8 | 2008_Mossong_PT | [Mossong *et al.* 2008] |
| 9 | 2008_Mossong_UK | [Mossong *et al.* 2008] |
| 10 | 2009_Hens_BELGIUM | [Hens *et al.* 2009] |
| 11 | 2010_Willem_BELGIUM | [Willem *et al.* 2012] |
| 12 | 2011_Horby_Vietnam | [Horby *et al.* 2011] |
| 13 | 2015_Beraud_France | [Béraud *et al.* 2015] |
| 14 | 2015_Grijalva_Peru | [Grijalva *et al.* 2015] |
| 15 | 2016_Litvinova_Russia | [Litvinova *et al.* 2019] |
| 16 | 2017_Leung_HongKong | [Leung *et al.* 2017] |
| 17 | 2017_Melegaro_Zimbabwe | [Melegaro *et al.* 2017] |
| 18 | 2019_Zhang_China | [Zhang *et al.* 2020] |
| 19 | 2020_Mahikul_Thailand | [Mahikul *et al.* 2020] |
| 20 | 2015_Dodd_ZambiaAndSA | [Dodd *et al.* 2015] |

**Figure 11:**
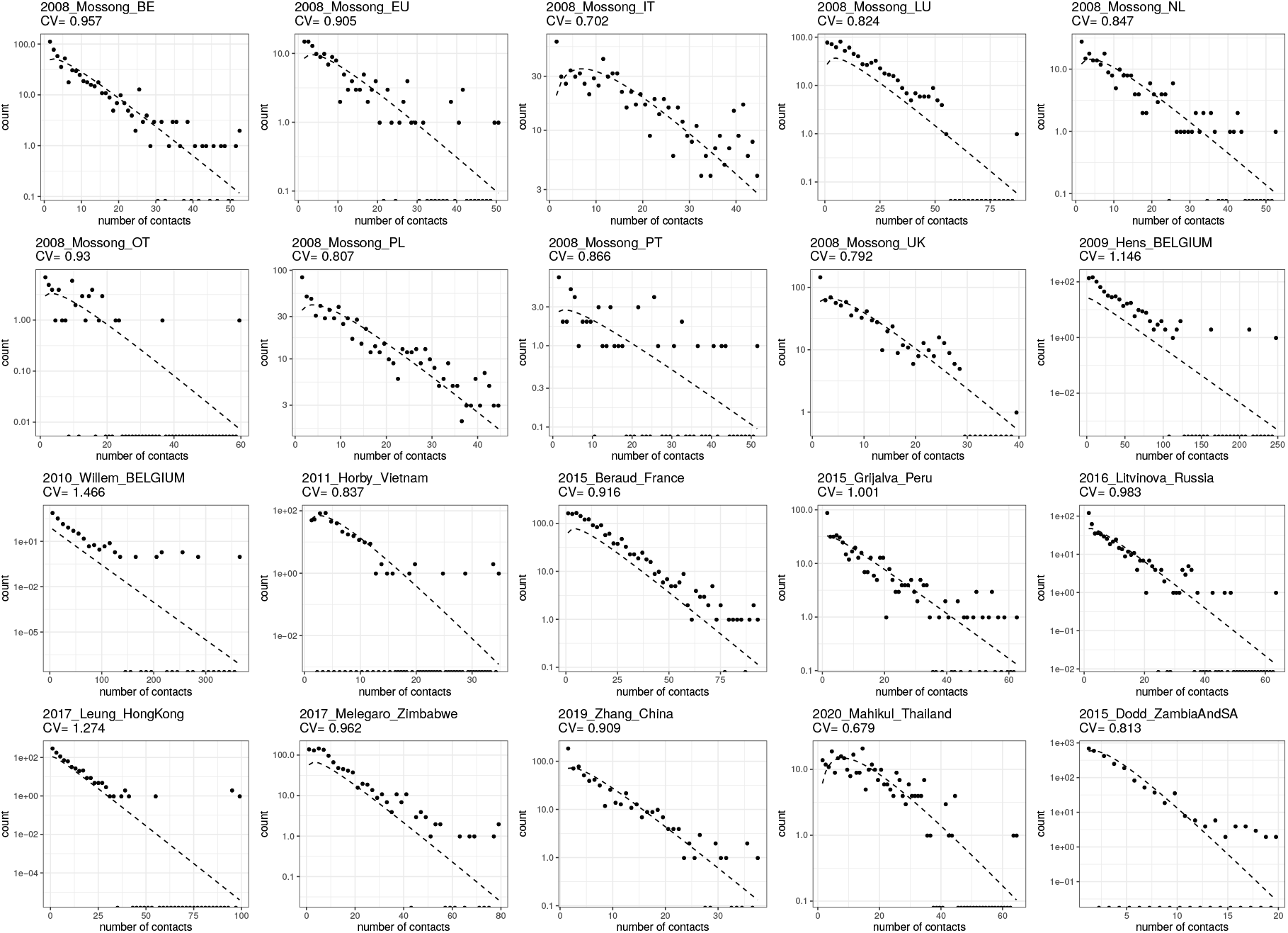
Gamma fits for the included contact surveys. For each dataset in Table 5, we plot the empirical distribution and report its CV, as well as the best-fit gamma distribution.

Irrespective of fits, the empirically measured contact distributions reveal CV between 0.7 and 1.5 (depending on study and setting; mean 0.9, standard deviation 0.2).

In addition to the magnitude of individual variation in connectivity, the time scale of that variation is another important determinant for the effect of selection in accelerating the acquisition of population immunity [Tkachenko *et al*. 2021]. The referenced studies typically report contact patterns for individuals over a very short (e.g., 1-day) period which is insufficient for assessing persistence of the measured variation. One of the studies [Hens *et al*. 2009] made an extra step and measured contact patterns for each individual on two different days (one weekday and one weekend day). In Fig. 12, we show fits for contact patterns by these two days individually, and for the average. The CV for the contact heterogeneity that persists over the two days is approximately 1.1 (in contrast with the larger 1.4 or 1.6 for each day alone).

**Figure 12:**
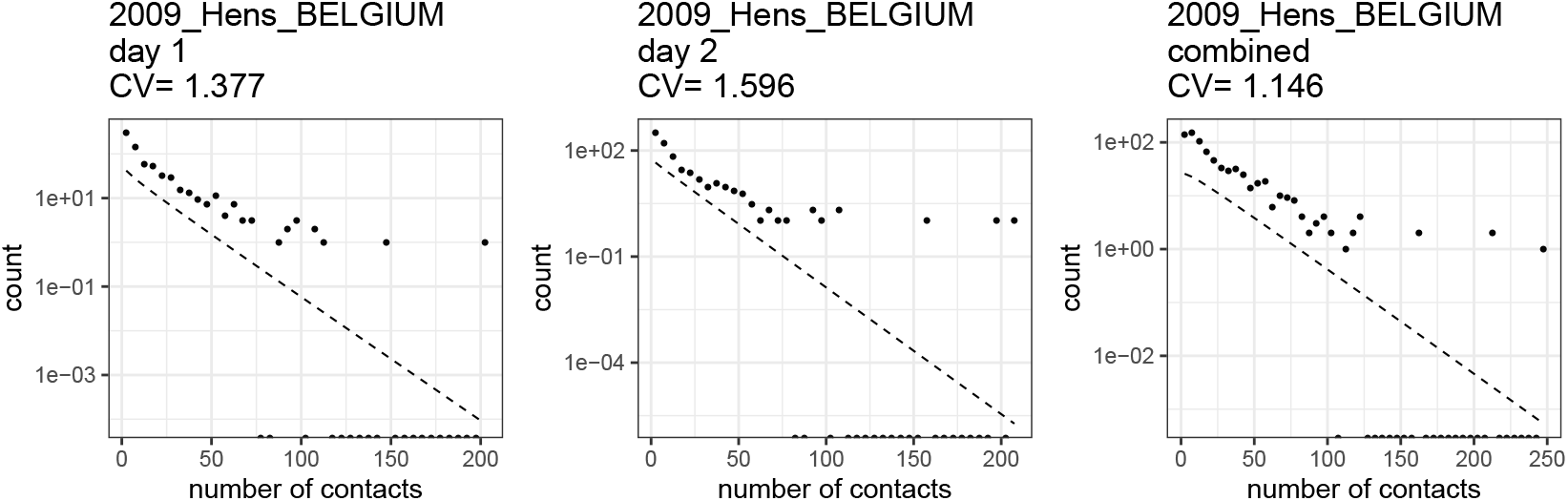
Persistence of contact heterogeneity. For this dataset, contact distributions are available for two different diary days for each participant. When averaging over these two separate diary days, the contact distribution CV is 17 − 28% lower.

For a heterogeneous model of an epidemic unfolding over a timescale of years, local dynamics are driven by assumptions about heterogeneity in short-term (e.g., week-long) averages in contact patterns, while global dynamics depend on assumptions about persistence of heterogeneity in those short-term averages over the timescale of the simulation. However, these long-term averages are frequently not evaluated directly by contact diary experiments.

One way to estimate persistent contact heterogeneity from below is to bin contact data by age groups, assuming for example that heterogeneity in contact patterns which persists in contact data after binning in 5-year age groups represents population-level heterogeneity in contact patterns that is persistent on multi-year time scales. Of course, this approach only captures heterogeneity mediated by age; i.e., it would only capture the full extent of heterogeneity in contact patterns if there was no within-age-group persistent variation in contact patterns. As such we should expect age-binned contact data to underestimate the level of persistent heterogeneity in contact patterns [Britton *et al*. 2020], perhaps quite substantially. In Fig. 13 we show the effect on CV of binning the data from the studies in Table 5 in 1- and 10-year age groups, respectively. Generally, binning by larger periods tends to reduce heterogeneity more, although 1-year binning is sufficient to bring the results of all studies to values below our model-based inference of persistent heterogeneity based on epidemic trajectories (CV = 1.1 for England and Scotland, as in Table 1, heterogeneous connectivity). When CV is artificially reduced by averaging processes such as these, models based on the resulting age-structured matrices are likely to overestimate the size of epidemics.

**Figure 13:**
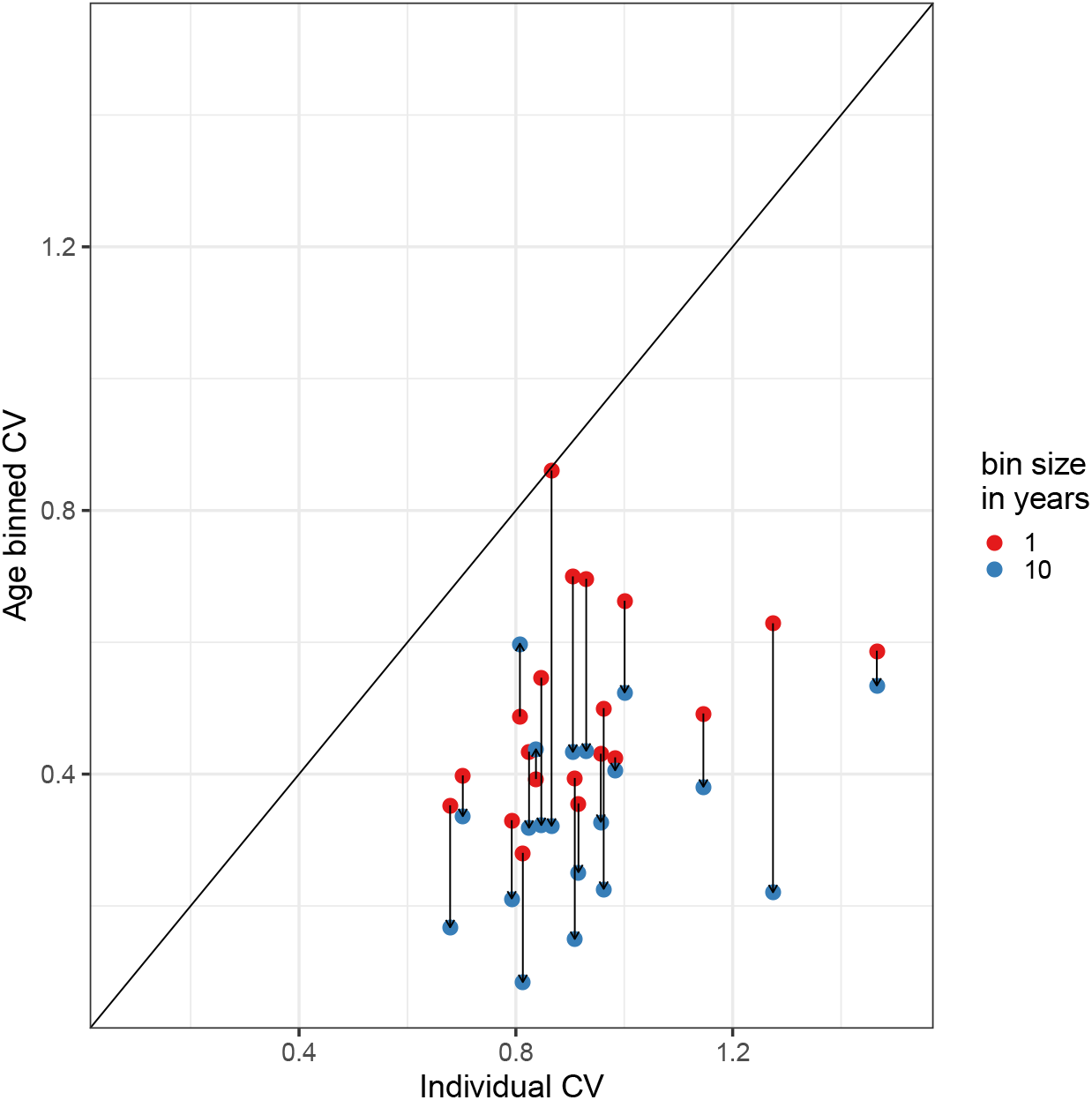
The effect of binning by age. Each linked pair represents one contact dataset; the height of the red dot shows the CV of the contact distribution when ages are binned in 1 year buckets, while the height of the blue dot shows the CV of the contact distribution when binned in 10 year buckets.

#### 5.4.2. Contact tracing

Contact-tracing data can also be used to describe heterogeneity in transmission by measuring directly the number of secondary cases attributed to each infected individual. Distributions of individual infectivity can then be characterised as in [Lloyd-Smith *et al*. 2005] who have analysed several pathogens including SARS-CoV-1. The authors found the measured distributions to be highly skewed around ℛ_0_, with a CV of approximately 2.6 for SARS-CoV-1. [Adam *et al*. 2020] conducted one of the earlier such studies for SARS-CoV-2 (there are now many more but a review of this literature is beyond the scope of this work). The authors estimate parameters that correspond to a CV around 2.1. These studies traditionally assume a negative binomial model and parameterise the resulting distributions by a dispersion parameter *k*. From that we derived the corresponding CVs as the square root of the variance (calculated as ℛ_c_(1 + ℛ_c_*/k*)) divided by the mean ℛ_c_.

Estimates of CV obtained from contact-tracing data are larger (roughly twice) those resulting from contact surveys (Section 5.4.1, Fig. S12), presumably because they capture variation in infectiousness as well as connectivity and, without further information, the two causes are inseparable.

### 5.5. Herd immunity thresholds and epidemic sizes

Fig. 14 shows the expected downward trends in the natural herd immunity threshold ℋ for SARS-CoV-2 as coefficients of variation of the gamma distribution increase (formulas (14) and (22)) considering an approximate basic reproduction number of ℛ_0_ = 3 (black and grey curves). Final sizes of the corresponding unmitigated epidemics (R^*\**^) are shown in blue. In contrast with ℋ, the final size R^*\**^ cannot be derived explicitly but can be formulated implicitly as

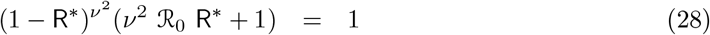

for the variable susceptibility model (9)-(12),

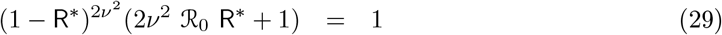

for variable connectivity (17)-(20), and

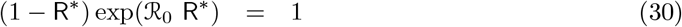

for the homogeneous scenario [Diekmann *et al*. 2013].

**Figure 14:**
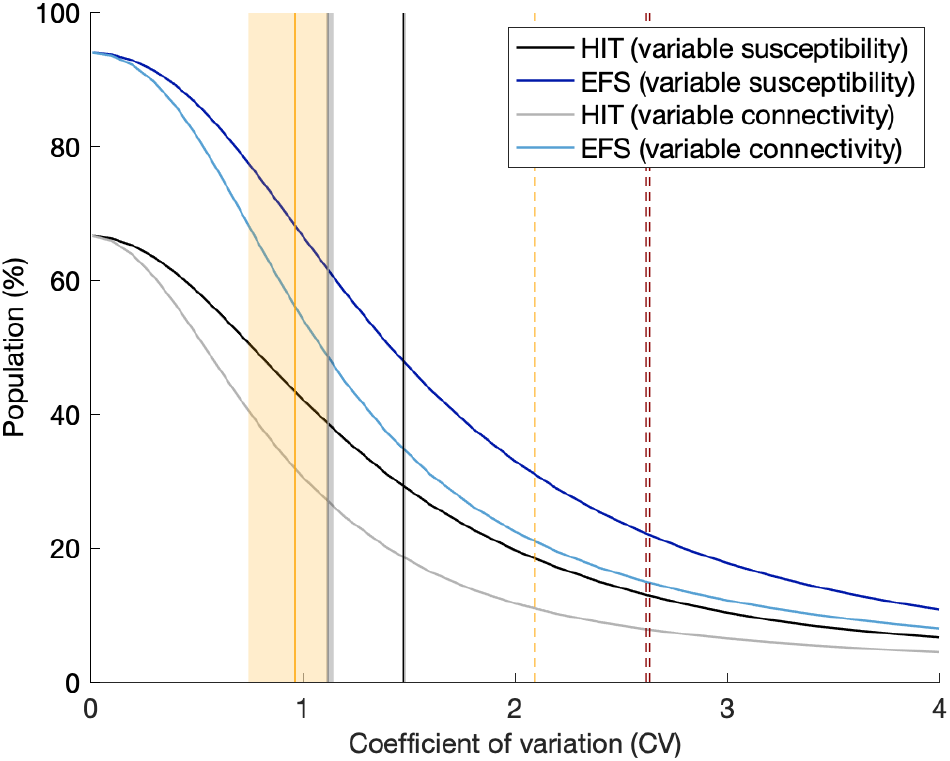
Herd immunity threshold and epidemic final size with gamma distributed susceptibility and exposure to infection. Curves generated using models (9)-(11) (dark tones) and (17)-(19) (light tones) with approximate ℛ_0_ = 3: herd immunity threshold given by (14) and (22) (black and grey, respectively); final size of unmitigated epidemics given implicitly by (28) and (29) (blue). Vertical lines indicate coefficients of individual variation from the literature and this study: susceptibility to SARS-CoV-2 (black) (England and Scotland 1.48, this study Table 1); connectivity for SARS-CoV-2 (grey) (England and Scotland 1.12, this study Table 1); connectivity (solid orange) (mean 0.93, standard deviation 0.19, as reviewed in Section 5.4.1); infectivity for SARS-CoV-2 (dashed orange) (Hong Kong 2.09 [Adam *et al*. 2020]); infectivity for SARS-CoV-1 (dashed red) (Singapore 2.62, Beijing 2.64 [Lloyd-Smith *et al*. 2005]).

Our inferences for CV from fittings to COVID-19 daily deaths until 1 February 2021 (Table 1) are marked as vertical grey lines. We use dark tones for heterogeneous susceptibility and light tones for heterogeneous connectivity.

We searched the literature and found no quantitative estimates of individual variation in susceptibility to infections although the evidence of its existence is documented [Carr *et al*. 2016]. Studies that quantify variation in connectivity are more prolific, as reviewed in Section 5.4 and marked by the vertical coloured lines in Fig. 14. Two types of studies were found: contact surveys (solid orange); and contact tracing (dashed red for SARS-CoV-1, and dashed orange for SARS-CoV-2). Our estimates (light grey) lie between those from these two study types.

The natural HIT resulting from our study (Section 5.1) can be visualised at the intersection between grey vertical lines and grey HIT curves. In both cases we obtain ℋ ≈ 25 − 27% in England, and ℋ ≈ 26 − 29% in Scotland, suggesting conserveness across the two models (variable susceptibility and variable connectivity). A combined formulation should result in intermediate CV estimates (currently 1.5 with variable susceptibility only, and 1.1 with variable connectivity) but similar HIT. We also find that once sufficient selection has occurred in the pandemic, our inferences become robust over different time scales (until 1 February 2021, to capture first and second waves, or until 1 July 2020, to capture the first pandemic wave only) contrasting with results from [Tkachenko *et al*. 2021] on the effects of time scales of individual variation on COVID-19 dynamics.

To explore sensitivity to the type of distribution we show equivalent plots to Fig. 14 with lognormal distributions in Supplementary Material (Section S5, Fig. S16). The curves for variable connectivity remain almost unaltered while the decline with increased CV becomes less steep for variable susceptibility.

### 5.6. Interpretation and counterfactuals

For over a century, mathematical epidemiologists have realised that individual variation in susceptibility and exposure to infection were key to determine the shape of epidemic curves. Models that underrepresent these forms of variation tend to overpredict epidemic sizes and consequently inflate the effects attributed to control measures. Fig. 15(a) depicts the epidemic curves that might have resulted from letting the COVID-19 pandemic run unmitigated in England (results for Scotland would be similar). Using parameters estimated in this study (Section 5.1), the homogeneous model (black) would have predicted almost half million deaths in total (460,000), while accounting for individual variation in susceptibility (red) would have brought these estimates down to less than a half (225,000) (individual variation in exposure leading to a similar result). The maximum number of deaths in a single day would have been around 16,000 by the homogeneous model and 6,000 when variability was considered. The figure also includes the real data for England (green dots), where the number of deaths in a single day has peaked at slightly over 1,200 in January 2021, a reduction attributed to adaptive behavioural changes and NPIs (but see [Wood 2021]). Although, in either case, it seems unimaginable that one would let a pandemic like this go unmitigated, the scenario appears more than twice as dramatic when individual variation is neglected.

**Figure 15:**
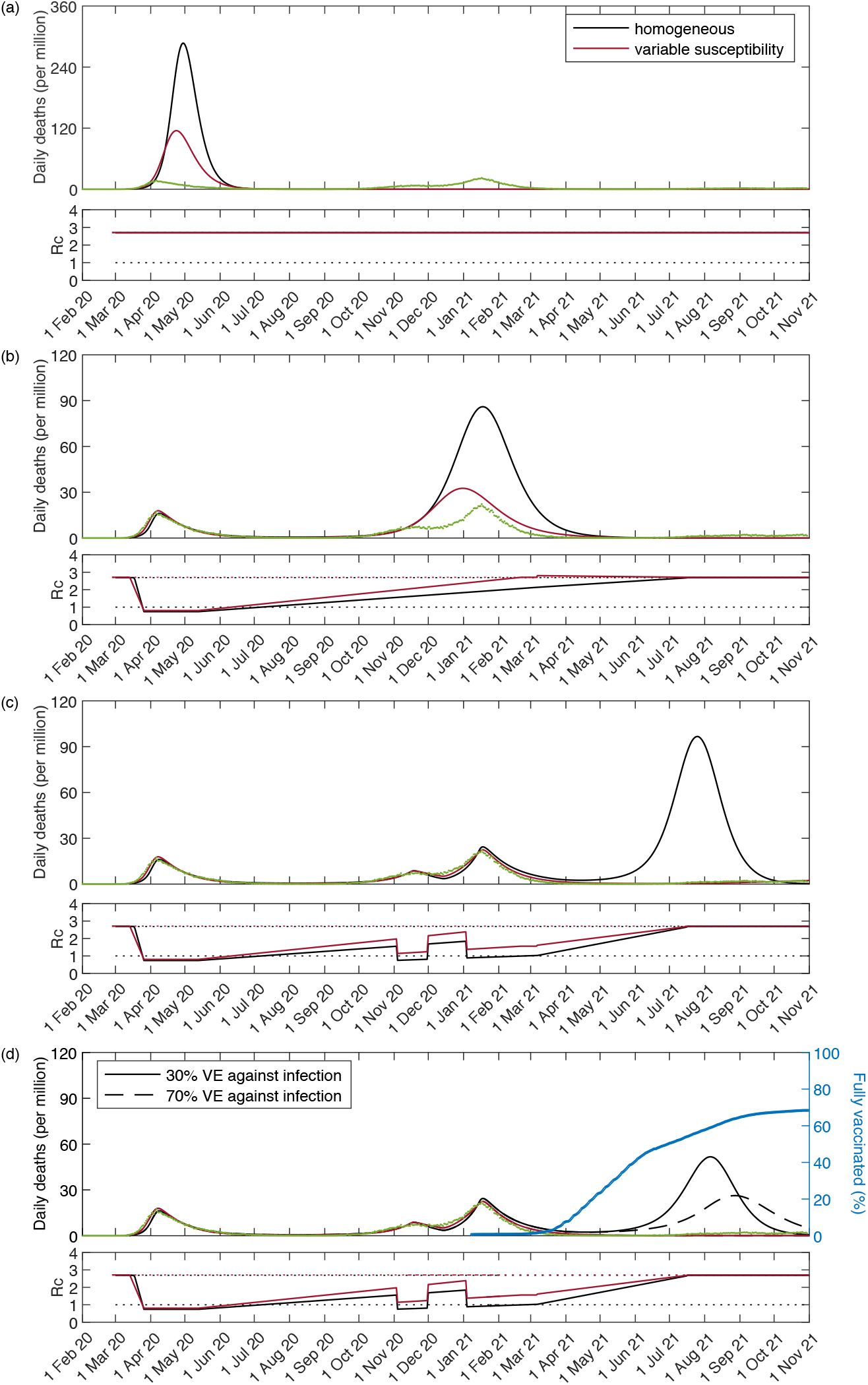
Counterfactuals. Simulations of the COVID-19 pandemic in England with epidemiological parameters estimated in Section 5.1 and interventions implemented incrementally: (a) no interventions (unmitigated epidemic); (b) lockdown 1 only (spring 2020); (c) lockdown 1, as well as 2 and 3 (autumn and winter 2020-2021); (d) lockdowns 1, 2 and 3, as well as vaccination from late 2020 / early 2021 (30% vaccine efficacy (VE) against infection (solid); 70% VE against infection (dashed); 90% VE against death in both cases). Shallow panels show controlled reproduction numbers as introduced in Section 2.1. Cumulative percentage fully vaccinated (two doses) in blue. Curves generated by running model (31)-(39): (red) heterogeneous susceptibility; (black) assuming homogeneity.

Moving down Fig. 15, panel (b) simulates what would have happened according to both models with the first lockdown implemented (as it did in the spring 2020), but not the second and third (in the autumn-winter 2020-2021). In this scenario, the homogeneous model would have predicted around 320,000 deaths over an 8-month period around the peak (September 2020 - April 2021), while individual variation in susceptibility would have brought this projection down to around 135,000. The number of predicted COVID-19 deaths in a single day would have been around 4,800 by the homogeneous model and 1,800 when individual variation was considered. The gap between homogeneous and heterogeneous models appear to widen as the pandemic progresses but, more strikingly, the heterogeneous model (with first lockdown but without second and third) is not far from the real data (which included lockdowns 2 and 3).

Fig. 15(c) simulates all three lockdowns as estimated in Section 5.1, but without vaccines. The third lockdown is assumed to last until 7 March 2021, after which we let the controlled basic reproduction number ℛ_c_ initiate a linear increase towards normalcy (here assumed to reach the original ℛ_0_ by 19 July 2021), when most restrictions were removed in England (but see further discussion below in regards to increased transmission due to viral evolution). In the absence of vaccination, a large exit wave would have been expected under the homogeneous model but only a shallow hump is apparent when individual variation is considered.

Finally, the effects of vaccination are explored in the bottom panel (Fig. 15(d)). We simulate a vaccination programme according to the proportion *v*(*t*) of the population fully vaccinated each day (two doses was the standard full vaccination schedule at the time) as recorded in England (blue):

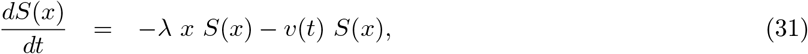

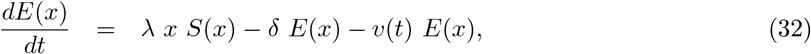

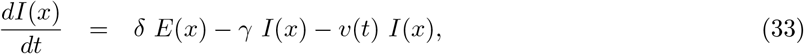

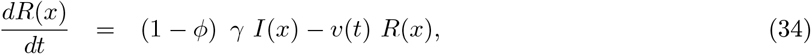

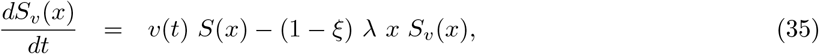

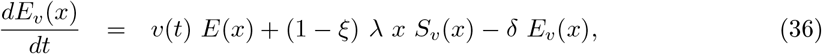

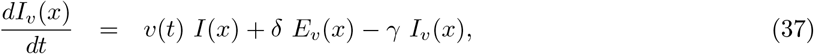

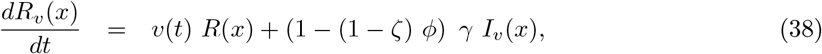

with force of infection

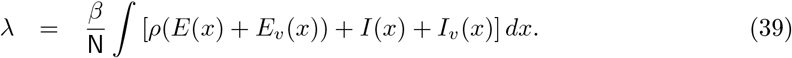

The vaccine is assumed to confer either *ξ* = 30% (solid curves) or *ξ* = 70% (dashed) protection against infection and 1 − (1 − *ξ*) *·* (1 − *ζ*) = 90% against death [Voysey *et al*. 2020, Bernal *et al*. 2021, Sheikh *et al*. 2021]. The implementation of a programme with these characteristics would have reduced the size of the third waves as visualised by comparing Figs. 15(c) and 15(d). According to the homogeneous model the third wave would have nevertheless remained large (black), while under heterogeneity the vaccination programme would have brought model trajectories (red) to the level of the data (green).

To better assess the dynamics governed by the heterogeneous model see Fig. 16(a),(b). We separate the scenarios of 30% and 70% vaccine efficacy against infection in two panels. We also allow ℛ_c_ to reach a value different from baseline after restrictions are lifted on the 19 July 2021. In each vaccine efficacy scenario we calculate the value that ℛ_c_ would have to reach for the number of COVID-19 deaths generated by the model between 1 February 2021 and 1 November 2021 to match the data (26, 129 deaths over the stipulated 9 months). We obtained ℛ_c_ = 1.23 *·* ℛ_0_ in the 30% scenario and ℛ_c_ = 1.41 *·* ℛ_0_ in the 70% scenario. Subtracting the number of deaths under vaccination from the corresponding simulations without vaccination would lead us to conclude that the vaccine would have prevented around 52, 000 and 88, 000 deaths in the 30% and 70% vaccine efficacy scenarios, respectively. This calculation is not possible with the homogeneous model because no vaccination programme in that model can fill the gap between projections in Fig. 15 (c) and the reported deaths (see Fig. 16(c),(d)). Scenarios equivalent to those provided for the heterogeneous model result in epidemic waves one order of magnitude larger when heterogeneity is omitted. Heterogeneous models, which performed better than homogeneous in statistical inferences in this paper (whether based on fittings until July 2020 or February 2021, with or without reinfection, assuming different infection fatality ratios, informed by government stringency indices and mobility data or not), also generate projections in closer agreement with data beyond the fitted period.

**Figure 16:**
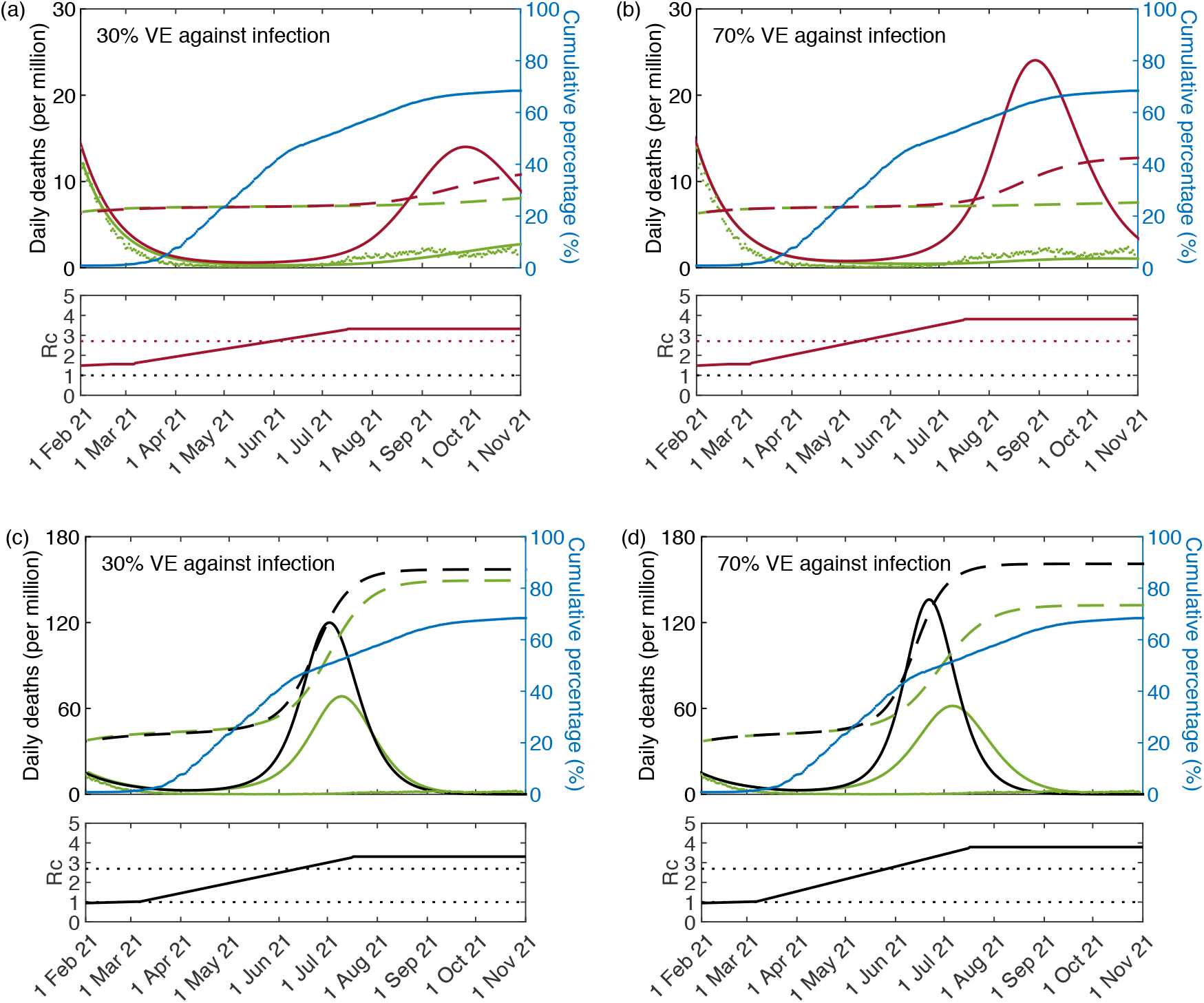
Scenarios allowing transmissibility to increase beyond initial ℛ_0_. Simulations of the COVID-19 pandemic in England with epidemiological parameters estimated in Section 5.1. Larger panels show daily COVID-19 deaths in England while shallow panels underneath show the corresponding controlled reproduction numbers as introduced in Section 2.1. (a,b) heterogeneous susceptibility model: without vaccination (red); and with cumulative vaccination as per blue dots (green curve); (c,d) homogeneous model: without vaccination (black); and with vaccination (green curve). Dashed curves represent cumulative percentage infected in the respective colours. Reported COVID-19 deaths are represented by green dots. Two scenarios are considered for VE against infection: 30% (a,c); and 70% (b,d). VE against death is fixed at 90%. Curves generated by running model (31)-(39).

In the timeframe of this study, SARS-CoV-2 has undergone substantial evolution, with dominant genetic types being replaced by more transmissible variants. Specifically, the larger wave which peaked in January 2021 was caused by the alpha variant (B.1.1.7), estimated as up to 30% more transmissible than early viruses, while the flatter wave in the summer 2021 was mostly due to delta (B.1.617.2), estimated about 40% more transmissible than alpha [PHE 2021, Richard *et al*. 2021, Wang *et al*. 2021] and hence 80% more than early types. The crude analysis in Fig. 16 suggests that ℛ_c_ might have been 23 − 41% higher during the delta wave than in the early months of the pandemic (baseline ℛ_0_). This is consistent with people retaining cautious behaviours which cancel some of the increases in transmissibility, e.g., a virus that is 80% more transmissible than the original (wild type) opposed by 22% reduced transmission due to behavioural change has ℛ_c_ = 1.4 *·* ℛ_0_. In December 2021, delta was replaced by the omicron variant (B.1.1.529) [Elliott *et al*. 2022], assessed as even more transmissible but less pathogenic, a phenomenon that would be interesting to analyse with the models presented here but is beyond scope of the present study.

Throughout the COVID-19 pandemic, our heterogeneity models have systematically projected lower infection rates in the UK (more specifically, England and Scotland) than others conforming to more common approaches [Flaxman *et al*. 2020, Knock *et al*. 2021, Sonabend *et al*. 2021, Davies *et al*. 2020, Davies *et al*. 2021, Hilton *et al*. 2020,Keeling *et al*. 2021a, Keeling *et al*. 2021b, Moore *et al*. 2021]. This was also noticed in similar analyses featuring Spain and Portugal [Aguas *et al*. 2020]. When we disable individual variation in susceptibility and exposure to infection we obtain results similar to those presented elsewhere, suggesting that to be the distinguishing feature that prevents our models from overpredicting infection rates. When mitigations are applied the discrepancy between homogeneous and heterogeneous trajectories increases as the pandemic progresses, eventually causing the rejection of models that lack sufficient heterogeneity. By then, however, such models would have over-attributed impact to interventions, and may have induced governments to systematically make suboptimal decisions by over-weighting benefits in relation to the costs and collateral damages of control measures. More work is need to establish to what extent these might have happened in the COVID-19 pandemic.

## 6. Limitations of the study

We have presented an original approach to modelling pandemics that includes inference of individual variation from its response to selection and enables reliable projections based on series of daily counts of disease outcomes (deaths in this case, but it could have been hospitalisations, reported infections, or a combination of multiple data streams) complemented by minimal information about interventions (Sections 5.1 and 5.2). Future developments might integrate more detailed data about interventions, such as NPIs and vaccines, the emergence and spread of new variants, individual variation in vulnerability to infection due to age, comorbidities or other factors, seasonal dynamics, susceptibility replenishment through waning immunity or birth and deaths, and interactions among these and other processes. Based on explorations initiated here (Sections 5.3 and 5.6) we expect our results to be robust to such extensions given abounding agreements with empirical evidence from independent sources (Section 5.4 and 5.5). Specifying more detail into the models does not necessarily enable more accurate predictions, but there are often other reasons, such as informing the design of targeted interventions, for such developments [Mishra *et al*. 2020]. Although those extensions are beyond the scope of the current paper they are sometimes essential in specific circumstances.

Another aspect where this work may fall short of expectations, and we hope to stimulate further research, is in the treatment of uncertainty. Our inferences result in noticeably narrow credible intervals which may inadvertently suggest overconfidence. Factors that may have contributed to this include the lack of account for uncertainty around some input parameters (such as incubation period, recovery rate and infection fatality ratio) which we assume at fixed values taken from the literature, the absence of stochasticity along individual trajectories (although we consider between individual variability) and the choice of a particular Monte Carlo algorithm [Stapor *et al*. 2018] implemented in Matlab while alternatives are always worth exploring given the rapid development of MCMC methods (although we have replicated a sample of fittings with other algorithms in Stan and obtained similar results). Given the wide discrepancy between heterogeneous and homogeneous model outputs and the consistency across a spectrum of scenarios, however, methodological improvements along these lines are not expected to affect the conclusions of this study. They may nevertheless reduce the confidence on particular metrics being estimated.

## 7. Discussion and conclusion

The conclusions of this work apply much beyond COVID-19. Individual variation in the propensity to acquire infections has many factors. Individuals differ in genetic background, the local environment they live in, and social activity, in ways that affect their susceptibility and exposure to many infectious diseases. Immunological studies identify local environmental conditions as key in shaping the human immune system [Carr *et al*. 2016], while social inequality is probably a main driver [Cevik and Baral 2021]. Studies designed to identify specific factors and characterise their modes of action are key, not necessarily to inform the development of more predictive models but primarily to help focusing interventions on those individuals who need more protection. Notwithstanding these studies being conducted, there will always remain some unobserved variation and, for the sake of model predictability, all variation responding to selection by natural infection must be accounted for to prevent biased outputs. For this reason we took the inverse approach of quantifying selectable variation by fitting fairly generic models to epidemic curves. We used the COVID-19 pandemic in England and Scotland to test the approach. We estimate coefficients of variation for variable connectivity (around 1.1) in agreement with empirical studies of contact patterns, resulting in natural herd immunity thresholds around 25 − 28%. For variable susceptibility we are not aware of available data specific to human COVID-19 to immediately compare our inferences (CV around 1.5, leading to herd immunity thresholds around 27 − 29%).

The natural herd immunity threshold used here (ℋ) describes a ceiling for epidemic growth formulated in terms of the basic reproduction number (ℛ_0_) and coefficients of individual variation in traits that are under selection by natural infection (namely, susceptibility and exposure to infection), filling a gap in mathematical epidemiology. While ℛ_0_ is indicative of the early growth of an epidemic, it says little about the final size and the height of the peak, which are strongly dependent on types of individual variation that decrease transmission over time (see also [Brauer 2018]). ℋ accounts for that variation enabling more accurate predictions of how large an epidemic might be, but it is a theoretical framework to the extent that ℛ_0_ is a theoretical framework. ℋ changes if the parameters that determine its value change. Most notably, natural changes in ℛ_0_ through time, which can happen due to seasonal forces or viral evolution, will manifest in ℋ. In particular, the percentage of the population immune required to prevent sustained epidemic growth may become higher than the initially estimated ℋ if we enter a high-transmission season or if the virus evolves towards higher transmissibility.

The UK experienced a first wave of the wild type virus in the spring 2020 (low-to-moderate transmission season but highly susceptible population) and initiated a second wave in the autumn of the same year (presumably driven by seasonality). The wave was subsiding (due to acquired immunity and contact restrictions) when a more transmissible variant (alpha) emerged, reaccelerating epidemic growth and becoming dominant by January 2021. By the end of January 2021, this wave was retracting again. Levels of infection and disease were especially low throughout April-May 2021 but this was somewhat altered by an even more transmissible variant (delta) that become dominant by the end of May 2021. The delta variant, which ravaged parts of the world (such as India, where it was first detected), maintained moderate levels of infection and disease in the UK throughout the summer and autumn 2021 (immunity due to previous infection and vaccination having a key role in this outcome). By the time this study was completed (November 2021), a new variant was being detected (omicron, associated with even higher transmissibility, lower incubation period and lower pathogenicity) but this is beyond the scope of this paper.

Although our models have not been developed to estimate variant-specific parameters (except for the wild type), they can give an indication of which parameter ranges are compatible with the transmission dynamics in each nation at the time a particular variant was dominant. In England, assuming an infection fatality ratio of 0.9%, our heterogeneous models estimate ℋ for wild type in the range 25 − 27% (ℛ_0_ around 2.7). Assuming alpha 30% more transmissible, and delta 40% more transmissible than alpha [PHE 2021], we obtain ℋ in the range 30 − 33% (ℛ_0_ around 3.6) for alpha, and 36 − 40% (ℛ_0_ around 5.0) for delta. The estimated cumulative infected percentage is close to 25% by November 2021, suggesting that the nation was then entering the range estimated for the natural herd immunity threshold for wild type SARS-CoV-2 (assuming acquired immunity to be effective across variants), but remained 5% and 11% below threshold for alpha and delta variants, respectively. Adding that almost 70% of the population had been fully vaccinated by then, it might have been reasonable to expect herd immunity to be achieved soon for the variants as well. This was happening just prior to the rapid rise of the omicron variant by December 2021.

In Scotland, we estimate ℋ in the range 26 − 29% (ℛ_0_ around 2.9) for the wild type, 31 − 34% (ℛ_0_ around 3.8) for alpha, and 37 − 41% (ℛ_0_ around 5.3) for delta, where the same infection fatality ratio of 0.9% would suggest a cumulative infected percentage close to 20% by November 2021. In this case, the nation remained 6% below the natural herd immunity threshold for the wild type, 11% for alpha, and 17% for the delta variant. As in England, though, it is plausible the addition of vaccination coverage would have brought the nation to the herd immunity threshold for the three SARS-CoV-2 types, but more research is needed to determine this with satisfactory confidence.

These figures are for the scenario where the IFR is 0.9% and should be adapted if different assumptions are made about this metric, but explorations reported in Supplementary Material suggest that general tendencies will not be affected. Also worth noting that although neither the wild type nor the alpha variant remained in circulation by November 2021, it is of both theoretical and practical interest to detail the mechanisms that resulted in their elimination. In particular, our analyses suggest that variant-specific naturally acquired immunity had a much larger role than what might have been expected according to models that take less account of individual variation in susceptibility and exposure to infection (recall that models that assume homogeneity inflate herd immunity thresholds to 60% and higher). More generally, heterogeneity affects competition mechanisms and measures of relative fitness between variants [Gomes *et al*. 2019], and efficacy of vaccines over time [Gomes *et al*. 2016, Gomes 2020], in ways that are yet to be considered for COVID-19 [Bushman *et al*. 2021, Althaus *et al*. 2021, Thomas *et al*. 2021].

Finally, the maintenance of herd immunity is a dynamic process [Gomes 2021]. Immune individuals lose their immunity or die while new susceptibles are born. Further infections and vaccinations replenish population immunity maintaining infection rates at an endemic equilibrium - the herd immunity threshold. This equilibrium is precarious in the sense that it is sensitive to viral evolution and demographic change, and may be temporarily eroded by seasonality [Bacaër and Gomes 2009] in a pattern exhibited by many common air-borne or vector-borne infectious diseases.

## Data Availability

All data referred to in the manuscript are publicly available.

## Acknowledgements

We thank Jorge Carneiro, José Ferreira Machado, Kate Langwig, Robert MacKay, Paul McKeigue, Antonio Montalbán, Joe Schoneman and Laurette Tuckerman for valuable discussions throughout this study. At the University of Strathclyde, Matthew Burns, Zhichun Jiang, Naithan McNeil, Lauren Schofield and Aidan West conducted their final year BSc projects on Communicating Mathematics and Statistics, supervised by M.G.M.G., on topics related to this study while this paper was being written. This has contributed clarity to our presentation. The models presented here were first submitted to *medRxiv* on 27 April 2020 and posted soon after. Applications to the COVID-19 pandemic and subsequent updates followed. We are grateful to the preprint server for making our work available to interested readers in real time. M.U.F. received funding from Conselho Nacional de Desenvolvimento Científico e Tecnológico, Brazil.

## References

Aalen, O. O. (1988) Heterogeneity in survival analysis. Stat. Med., 7, 1121–1137.

Aalen, O. O., Valberg, M., Grotmol, T. and Tretli, S. (2015) Understanding variation in disease risk: the elusive concept of frailty. Int. J. Epidemiol., 4, 1408–1421.

Adam, D. C., Wu, P., Wong, J. Y., Lau, E. H. Y., Tsang, T. K., Cauchemez, S., et al. (2020) Clustering and superspreading potential of SARS-CoV-2 infections in Hong Kong. Nat. Med., 26, 1714–1719.

Aguas, R., Corder, R. M., King, J. G., Gonçalves, G., Ferreira, M. U. and Gomes, M. G. M. (2020) Herd immunity thresholds for SARS-CoV-2 estimated from unfolding epidemics. Preprint medRxiv, 10.1101/2020.07.23.20160762.

Althaus, C. L., Baggio, S., Reichmuth, M. L., Hodcroft, E. B., Riou, J., Neher, R. A., et. al. (2021) A tale of two variants: Spread of SARS-CoV-2 variants Alpha in Geneva, Switzerland, and Beta in South Africa. Preprint medRxiv, 10.1101/2021.06.10.21258468.

Arons, M. M., Hatfield, K. M., Reddy, S. C., Kimball, A., James, A., Jacobs, et al. (2020) Public Health–Seattle and King County and CDC COVID-19 Investigation Team. Presymptomatic SARS-CoV-2 infections and transmission in a skilled nursing facility. N. Engl. J. Med., 382, 2081–2090.

Bacaër, N. and Gomes, M. G. M. (2009) On the final size of epidemics with seasoanality. Bull. Math. Biol., 71, 1954–1966.

Ball, F. (1985) Deterministic and stochastic epidemic models with several kinds of susceptibles. Adv. Appl. Probab., 17, 1–22.

Béraud, G., Kazmercziak, S., Beutels, P., Levy-Bruhl, D., Lenne, X., Mielcarek, N., et al. (2015) The French connection: the first large population-based contact survey in France relevant for the spread of infectious diseases. PLOS One, 10, e0133203.

Bernal, J. L., Andrews, N., Gower, C., Gallagher, E., Simmons, R., Thelwall, S., et al. (2021) Effectiveness of Covid-19 Vaccines against the B.1.617.2 (Delta) Variant. N. Engl. J. Med., 385, 585–594.

Birrel, P., Blake, J., van Leeuwen, E., MRC Biostatistics Unit COVID-19 Working Group and De Angelis, D. (2021) MRC Biostatistics Unit Report on Nowcasting and Forecasting of the COVID-19 Pandemic - 29th October 2021. https://www.mrc-bsu.cam.ac.uk/now-casting/nowcasting-and-forecasting-29th-october-2021/.

Brauer, F. (2018) Early estimates of epidemic final sizes. J. Biol. Dyn., 13, 23–30.

Britton, T., Ball, F. and Trapman, P. (2020) A mathematical model reveals the influence of population heterogeneity on herd immunity to SARS-CoV-2. Science, 369, 846–849.

Bushman, M., Kahn, R., Taylor, B. P., Lipsitch, M. and Hanage, W. P. (2021) Population impact of SARS-CoV-2 variants with enhanced transmissibility and/or partial immune escape. Cell, 184, 6229–6242.

Carr, E. J., Dooley, J., Garcia-Perez, J. E., Lagou, V., Lee, J. C., Wouters, C., et al. (2016) The cellular composition of the human immune system is shaped by age and cohabitation. Nat. Immunol., 17, 461–468.

Cevik, M. and Baral, S. D. (2021) Networks of SARS-CoV-2 transmission. Science, 373, 162–163.

Chen, S., Flegg, J. A., White, L. J. and Aguas, R. (2021) Levels of SARS-CoV-2 population exposure are considerably higher than suggested by seroprevalence surveys. PLOS Comput. Biol., 17, e1009436.

Colombo, M., Mellor, J., Colhoun, H. M., Gomes, M. G. M. and McK-eigue, P. M. (2020) Trajectory of COVID-19 epidemic in Europe. Preprint medRxiv, 10.1101/2020.09.26.20202267.

Coutinho, F. A. B., Massad, E., Lopez, L. F., Burattini, M. N., Struchiner, C. J. and Azevedo-Neto, R. S. (1999) Modelling heterogeneities in individual frailties in epidemic models. Math. Comput. Model., 30, 97–115.

Davies, N. G., Abbott, S., Barnard, R. C., Jarvis, C. I., Kucharski, A. J., Munday, J. D., et al. (2021) Estimated transmissibility and impact of SARS-CoV-2 lineage B.1.1.7 in England. Science, 372, eabg3055.

Davies, N. G., Kucharski, A. J., Eggo, R. M., Gimma, A., and Edmunds W. J. (2020) Effects of non-pharmaceutical interventions on COVID-19 cases, deaths, and demand for hospital services in the UK: a modelling study. Lancet Public Health, 398, 1825–1835.

Diekmann, O., Heesterbeek, H. and Britton, T. (2013) Mathematical Tools for Understanding Infectious Disease Dynamics. Princeton University Press, Princeton, New Jersey.

Dodd, P. J., Looker, C., Plumb, I. D., Bond, V., Schaap, A., Shanaube, K., et al. (2015) Age-and sex-specific social contact patterns and incidence of Mycobacterium tuberculosis infection. Am. J. Epidemiol., 2, 156–166.

Dwyer, G., Elkinton, J. S. and Buonaccorsi, J. P. (1997) Host heterogeneity in susceptibility and disease dynamics: Tests of a mathematical model. Am. Nat., 150, 685–707.

Elliott, P., Bodinier, B., Eales, O., Wang, H., Haw, D., Elliott, J., et al. (2022) Rapid increase in Omicron infections in England during December 2021: REACT-1 study. Science, 10.1126/science.abn8347.

Fine, P., Eames, K. and Heymann, D. L. (2011) “Herd immunity”: a rough guide. Clin. Infect. Dis., 52, 911–916.

Finkenstädt, B. F. and Grenfell, B. T. (2000) Time series modelling of childhood diseases: a dynamical systems approach. Appl. Statist., 49, 187–205.

Flaxman, S., Mishra, S., Gandy, A., Unwin, H. J. T., Mellan, T. A., Coupland, H., et al. (2020) Estimating the effects of non-pharmaceutical interventions on COVID-19 in Europe. Nature, 584, 257–261.

Google (2020) COVID-19 Community Mobility Reports.

Gart, J. J. (1968) The mathematical analysis of an epidemic with two kinds of susceptibles. Biometrics, 24, 557–566.

Gart, J. J. (1971) The statistical analysis of chain-binomial epidemic models with several kinds of susceptibles. Biometrics, 28, 921–930.

Gomes, M. G. M., Gordon, S. B. and Lalloo, D. G. (2016) Clinical trials: The mathematics of falling vaccine efficacy with rising disease incidence. Vaccine, 34, 3007.

Gomes, M. G. M., King, J. G., Nunes, A., Colegrave, N. and Hoffmann, A. (2019) The effects of individual nonheritable variation on fitness estimation and coexistence. Ecol. Evol., 9, 8995–9004.

Gomes, M. G. M. (2020) A pragmatic approach to account for individual risks to optimise health policy. Preprint arXiv, 2009.01354.

Gomes, M. G. M. (2021) Timeliness and obsolescence of herd immunity threshold estimates in the COVID-19 pandemic. Public Health, 10.1016/j.puhe.2021.09.036.

Grijalva, C. G., Goeyvaerts, N., Verastegui, H., Edwards, K. M., Gil, A. I., Lanata, C.F ., et al. (2015) A household-based study of contact networks relevant for the spread of infectious diseases in the highlands of Peru. PLOS One, 10, e0118457.

Hale, T., Webster, S., Petherick, A., Phillips, T. and Kira, B. Oxford covid-19 government response tracker (2020) Blavatnik School of Government, 25.

Hall, V. J., Foulkes, S., Charlett, A., Atti, A., Monk, E. J., Simmons, R., et al. (2021) SARS-CoV-2 infection rates of antibody-positive compared with antibody-negative health-care workers in England: a large, multicentre, prospective cohort study (SIREN). Lancet, 397, 1459–1469.

He, X., Lau, E. H. Y., Wu, P., Deng, X., Wang, J., Hao, X., et al. (2020) Temporal dynamics in viral shedding and transmissibility of COVID-19. Nat. Med., 26, 672–675.

Hens, N., Goeyvaerts, N., Aerts, M., Shkedy, Z., Van Damme, P. and Beutels, P. (2009) Mining social mixing patterns for infectious disease models based on a two-day population survey in Belgium. BMJ Infect. Dis., 9, 1–18.

Hilton, J. and Keeling, M. (2020) Estimation of country-level basic reproductive ratios for novel Coronavirus (SARS-CoV-2/COVID-19) using synthetic contact matrices. PLOS Comput. Biol., 16, e1008031.

Horby, P., Thai, P. Q., Hens, N., Yen, N. T. T., Thoang, D. D., Linh, N. M., et al. (2011) Social contact patterns in Vietnam and implications for the control of infectious diseases. PLOS One, 6, e16965.

Hougaard, P. (1984) Life tables methods for heterogeneous populations. Biometrika, 71, 75–83.

Jarvis, C. I., Van Zandvoort, K., Gimma, A., Prem, K., CMMID COVID-19 working group, Klepac, P., et al. (2020) Quantifying the impact of physical distance measures on the transmission of COVID-19 in the UK. BMC Med., 18, 124.

Katriel, G. (2012) The size of epidemics in populations with heterogeneous susceptibility. J. Math. Biol., 65, 237–262.

Keeling, M., Hill, E. M., Gorsich, E. E., Penman, B., Guyver-Fletcher, G., Holmes, A., et al. (2021) Predictions of COVID-19 dynamics in the UK: Short-term forecasting and analysis of potential exit strategies. PLOS Comput. Biol., 17, e1008619.

Keeling, M. J., Guyver-Fletcher, G., Dyson, L., Tildesley, M. J., Hill, E. M. and Medley, G. F. (2021) Precautionary breaks: Planned, limited duration circuit breaks to control the prevalence of SARS-CoV2 and the burden of COVID-19 disease. Epidemics, 37, 100526.

Kermack, W. O. and McKendrick, A. G. (1927) A contribution to the mathematical theory of epidemics. Proc. R. Soc. Lond. A, 115, 700–721.

Kissler, S. M., Tedijanto, C., Goldstein, E., Grad, Y. H. and Lipsitch, M. (2020) Projecting the transmission dynamics of SARS-CoV-2 through the postpandemic period. Science, 368, 860–868.

Knock, E. S., Whittles, L. K., Lees, J. A., Perez-Guzman, P. N., Verity, R., FitzJohn, R. G., et al. (2021) Key epidemiological drivers and impact of interventions in the 2020 SARS-CoV-2 epidemic in England. Sci. Transl. Med., 13, eabg4262.

Kwok, K. O., Lai, F., Wei, W. I., Wong, S. Y. S. and Tang, J. (2020) Herd immunity – estimating the level required to halt the COVID-19 epidemics in affected countries. J. Infect., 80, e32–e33.

Lauer, S. A., Grantz, K. H., Bi, Q., Jones, F. K., Zheng, Q., Meredith, H. R., Azman, A. S., et al. (2020) The incubation period of coronavirus disease 2019 (COVID-19) from publicly reported confirmed cases: estimation and application. Ann. Intern. Med., 172, 577–582.

Leung, K., Jit, M., Lau, E. H. Y. and Wu, J. Y. (2017) Social contact patterns relevant to the spread of respiratory infectious diseases in Hong Kong. Sci. Rep, 7, 1–12.

Li, Q., Guan, X., Wu, P., Wang, X., Zhou, L., Tong, Y., et al. (2020) Early transmission dynamics in Wuhan, China, of novel coronavirus-infected pneumonia. N. Engl. J. Med., 382, 1199–1207.

Litvinova, M., Liu, Q.-H., Kulikov, E. S., Evgeny, S. and Ajelli., M. (2019) Reactive school closure weakens the network of social interactions and reduces the spread of influenza. Proc. Natl. Acad. Sci. U.S.A., 27, 13174–13181.

Lloyd-Smith, J. O., Schreiber, S. J., Kopp, P. E. and Getz., W. M. (2005) Superspreading and the effect of individual variation on disease emergence. Nature, 438, 355–359.

McKendrick, A. G. (1939) The dynamics of crowd infection. Edinb. Med. J., 47, 117–136.

McKeigue, P. M., McAllister, D. A., Caldwell, D., Gribben, C., Bishop, J., McGurnaghan, S., (2021) Relation of severe COVID-19 in Scotland to transmission-related factors and risk conditions eligible for shielding support: REACT-SCOT case-control study. BMC Med., 19, 149.

Mahikul, W., Kripattanapong, S., Hanvoravongchai, P., Meeyai, A., Iamsirithaworn, S., Auewarakul, P., et al. (2020) Contact mixing patterns and population movement among migrant workers in an urban setting in Thailand. Int. J. Environ. Res. Public Health, 17, 2237.

McAloon, C., Collins, A., Hunt, K., Barber, A., Byrne, A. W., Butler, F., et al. (2020) Incubation period of COVID-19: a rapid systematic review and meta-analysis of observational research. BMJ Open, 10, e039652.

Melegaro, A., Del Fava, E., Poletti, P., Merler, S., Nyamukapa, C., Williams, J., et al. (2017) Social contact structures and time use patterns in the Manicaland Province of Zimbabwe. PLOS One, 12, e0170459.

Miller, J. C., Slim, A. C. and Volz, E. M. (2012) Edge-based compartmental modelling for infectious disease spread. J. R. Soc. Interface, 9, 890–906.

Mishra, S., Kwong, J. C., Chan, A. K., and Barral, S. D. (2020) Understanding heterogeneity to inform the public health response to COVID-19 in Canada. Can. Med. Assoc. J., 192, E684–E685.

Montalbán, A., Corder, R. M. and Gomes, M. G. M. (2020) Herd immunity under individual variation and reinfection. Preprint arXiv, 2008.00098v2.

Moore, S., Hill, E. M., Tildesley, M. J., Dyson, L. and Keeling, M. J. (2021) Vaccination and non-pharmaceutical interventions for COVID-19: a mathematical modelling study. PLOS Comput. Biol., 17, e1008619.

Mossong, J., Hens, N., Jit, M., Beutels, P., Auranen, K., Mikolajczyk, R., et al. (2008) Social contacts and mixing patterns relevant to the spread of infectious diseases. PLOS Med., 5, e74.

Nickbakhsh, S., Ho, A., Marques, D. F. P., McMenamin, J., Gunson, R. N. and Murcia P. R. (2020) Epidemiology of seasonal coronaviruses: Establishing the context for the emergence of coronavirus disease 2019. J. Infect. Dis., 222, 17–25.

Nishiura, H., Linton, N. M. and Akhmetzhanov, A. R. (2020) Serial interval of novel coronavirus (COVID-19) infections. Int. J. Infect. Dis., 93, 284–286.

Novozhilov, A. S. (2008) On the spread of epidemics in a closed heterogeneous population. Math. Biosci., 215, 177–185.

Pastor-Satorras, R. and Vespignani, A. (2001) Epidemic dynamics and endemic states in complex networks. Phys. Rev. E, 63, 066117.

Public Health England (2021) SARS-CoV-2 variants of concern under investigation in England. Technical briefing 15.

Richard, D., Shaw, L. P., Lanfear, R., Acman, M., Owen, C.J., Tan, C. C. S., et al. (2021) A phylogeny-based metric for estimating changes in transmissibility from recurrent mutations in SARS-CoV-2. Preprint bioRxiv, 10.1101/2021.05.06.442903.

Sheikh, A., Robertson, C. and Taylor, B. (2021) BNT162b2 and ChAdOx1 nCoV-19 Vaccine Effectiveness against Death from the Delta Variant. N. Engl. J. Med., 385, 2195–2197.

Sonabend, R., Whittles, L. K., Imai, N., Perez-Guzman, P. N., Knock, E. S., Rawson, T., et a. (2021) Non-pharmaceutical interventions, vaccination, and the SARSCoV-2 delta variant in England: a mathematical modelling study. Lancet, 398, 1825–1835.

Stapor, P., Weindl, D., Ballnus, B., Hug, S., Loos, C., Fiedler, A., et al. (2018) PESTO: Parameter EStimation TOolbox. Bioinformatics, 34, 705–707.

Thomas, S. J., Moreira, E. D. Jr., Kitchin, N., Absalon, J., Gurtman, A., Lockhart, S., et al. (2021) Safety and Efficacy of the BNT162b2 mRNA Covid-19 Vaccine through 6 Months. New Engl. J. Med., 385, 1761–1773.

Tkachenko, A. V., Maslov, S., Elbanna, A., Wong, G. N., Weiner, Z. J. and Goldenfeld, N. (2021) Time-dependent heterogeneity leads to transient suppression of the COVID-19 epidemic, not herd immunity. Proc. Natl. Acad. Sci. U. S. A., 118, e2015972118.

To, K. K., Tsang, O. T., Leung, W. S., Tam, A. R., Wu, T. C., Lung, D. C., et al. (2020) Temporal profiles of viral load in posterior oropharyngeal saliva samples and serum antibody responses during infection by SARS-CoV-2: an observational cohort study. Lancet Infect. Dis., 20, 565–574.

Vaupel, J. H., Manton, K. G. and Stallard. E. (1979) The impact of heterogeneity in individual frailty in the dynamics of mortality. Demography, 16, 439–454.

Verity R., Okell, L. C., Dorigatti, I., Winskill, P., Whittaker, C., Imai, N., et al. (2020) Estimates of the severity of coronavirus disease 2019: a model-based analysis. Lancet Infect. Dis., 20, 669–677.

Voysey, J. L., Clemens, S. A. C., Madhi, S. A., Weckx, L. Y., Folegatti, P. M., Aley, P. K., et al. (2020) Safety and efficacy of the ChAdOx1 nCoV-19 vaccine (AZD1222) against SARS-CoV-2: an interim analysis of four randomised controlled trials in Brazil, South Africa, and the UK. Lancet, 397, 99–111.

Wang, L., Didelot, X., Bi, Y. and Gao, G. F. (2021) Comparative evaluation of the transmissibility of SARS-CoV-2 variants of concern. Preprint medRxiv, 10.1101/2021.06.25.21259565.

Ward, H., Atchison, C., Whitaker, M., Ainslie, K. E. C., Elliott, J., Okell, L., et al. (2021) SARS-CoV-2 antibody prevalence in England following the first peak of the pandemic. Nat. Commun., 12, 905.

Wei, W. E., Li, Z., Chiew, C. J., Yong, S. E., Toh, M. P. and Lee, V. J. (2020) Presymptomatic transmission of SARS-CoV-2 - Singapore, January 23-March 16, 2020. MMWR Morb. Mortal. Wkly. Rep., 69, 411–415.

Willem, L., Van Kerckhove, K., Chao, D.L., Hens, N. and Beutels, P. (2012) A nice day for an infection? Weather conditions and social contact patterns relevant to influenza transmission. PLOS One, 7, e48695.

Wood, S. N. (2021) Inferring UK COVID-19 fatal infection trajectories from daily mortality data: Were infections already in decline before the UK lockdowns? Biometrics, 10.1111/biom.13462.

Zhang, J., Litvinova, M., Wang, W., Wang, Y., Deng, X., Chen, X., et al. (2020) Evolving epidemiology and transmission dynamics of coronavirus disease 2019 outside Hubei province, China: a descriptive and modelling study. Lancet Infect. Dis., 20, 793–802.

