## Supplementary Material for "Individual variation in susceptibility or exposure to SARS-CoV-2 lowers the herd immunity threshold"

M. Gabriela M. Gomes
Marcelo U. Ferreira
Rodrigo M. Corder
Jessica G. King
Caetano Souto-Maior
Carlos Penha-Gonçalves
Guilherme Gonçalves
Maria Chikina
Wesley Pegden
Ricardo Aguas

### Contents

**S1 Sensitivity to model reductions** **S2**

**S2 Sensitivity to reinfection** **S8**

**S3 Sensitivity to infection fatality ratio** **S14**

S3.1 IFR = 0.7% . . . . . . . . . . . . . . . . . . . . . . . . . . . . . . . . . . . . . . . S14

S3.2 IFR = 1.0% . . . . . . . . . . . . . . . . . . . . . . . . . . . . . . . . . . . . . . . S19

**S4 Preliminary analysis of the first wave** **S23**

**S5 Sensitivity to distribution type** **S25**

#### S1 Sensitivity to model reductions

In the main text we present an infinite system of ODEs (1)-(4) to model the dynamics of an epidemic with individual variation in susceptibility to infection (variable susceptibility, with force of infection (5)) or individual variation in exposure to infection (variable connectivity, with force of infection (15)). We then introduce alternative finite versions, which were derived exactly in the case of variable susceptibility and approximately in the case of variable connectivity [Montalbán *et al.* 2020]. Results in the main text were obtained with reduced systems. Here we replicate the main results (Section 5.1) by runing the original infinite systems which account explicitly for the continuous distribution of the desired trait $x$ (susceptibility or connectivity):

$$\frac{dS(x)}{dt} = -\lambda \; x \; S(x), \tag{S1}$$

$$\frac{dE(x)}{dt} = \lambda \; x \; S(x) - \delta \; E(x), \tag{S2}$$

$$\frac{dI(x)}{dt} = \delta \; E(x) - \gamma \; I(x), \tag{S3}$$

$$\frac{dR(x)}{dt} = (1-\phi) \; \gamma \; I(x), \tag{S4}$$

where the force of infection $\lambda$ is written as

$$\lambda = \frac{\beta}{\mathsf{N}} \int (\rho E(x) + I(x)) \; dx, \tag{S5}$$

when variation is in susceptibility, and

$$\lambda = \frac{\beta}{\mathsf{N}} \frac{\int x\,(\rho E(x) + I(x)) \; dx}{\int xq(x) \; dx}, \tag{S6}$$

when it is connectivity that varies between individuals. There is a transmission coefficient $\beta$ to represent the average contact rate among individuals in the population, and infection history parameters $\delta$ and $\gamma$ representing the average rates of progression from exposure ($E$) to maximal infectiousness ($I$), and recovery from infection, respectively. The infection fatality ratio (IFR) is represented by parameter $\phi$.

Basic reproduction number $\mathcal{R}_0$ and herd immunity threshold $\mathcal{H}$ are as in the main text (Section 2), specifically,

$$\mathcal{R}_0 = \beta \left( \frac{\rho}{\delta} + \frac{1}{\gamma} \right) \qquad \text{and} \qquad \mathcal{H} = 1 - \left( \frac{1}{\mathcal{R}_0} \right)^{\frac{1}{1+\nu^2}} \tag{S7}$$

for variable susceptibility, and

$$\mathcal{R}_0 = \left(1+\nu^2\right) \beta \left( \frac{\rho}{\delta} + \frac{1}{\gamma} \right) \qquad \text{and} \qquad \mathcal{H} = 1 - \left( \frac{1}{\mathcal{R}_0} \right)^{\frac{1}{1+2\nu^2}} \tag{S8}$$

for variable connectivity [Montalbán *et al.* 2020].

Equivalent results to those in main text Figs. 3 and 4, with the explicit system (S1)-(S4) and gamma distributed traits, are shown in Fig. S1 (for variable susceptibility, force of infection (S5)) and Fig.S2 (for variable connectivity, force of infection (S6)). Homogeneous model results are the same as in Section 2 (Fig. 5) but nevertheless replicated here for convenience (Figure S3). Estimated parameters are in Table S1 (for comparison with Table 1).

Given the estimated values for $\mathcal{R}_0$ and CV ($\nu$) we derive the natural HIT using (S7) or (S8) as appropriate, obtaining $\mathcal{H}$ in the range $26-27\%$ in England, and $27-28\%$ in Scotland. With the homogeneous model, the inferences are considerably higher, $\mathcal{H}=63\%$ in England and in the range $66-67\%$ in Scotland.

We then prolong model trajectories (black dashed curves) for another 4 months (until 1 June 2021) to begin comparisons with data beyond the fitted period (red). All models project more deaths than observed, as expected given that the UK initiated a mass vaccination programme in late 2020 which would have started impacting the epidemic by February 2021. To mimic this we simulate the effective immunisation of 8% of the susceptible population per month from February onwards as a crude approximation to the UK programme by transferring that percentage of individuals to the immune compartment ($\mathsf{R}$) and depict the result by the red dashed curve in the figures. The rate of 8% per month was derived by noting that the UK reached 70% of its population fully vaccinated (two doses) by November 2021, mostly by the ChAdOx1 nCoV-19 (AstraZeneca) vaccine whose efficacy against symptomatic infection has been estimated around 70% [Voysey *et al.* 2020, Bernal *et al.* 2021, Sheikh *et al.* 2021], resulting in the effective immunisation of $0.7 \times 70\% \approx 50\%$ of the population. In order to reach 50% effective population protection at a constant rate over 9 months, approximately 8% would be vaccinated per month. Agreement with data (red dots in left panels) can be visually assessed in this exploration.

As in Section 5.1, we include separate panels on the right where the model is run for an additional 12 months to assess the prospects for an exit wave as contact restrictions were lifted from 8 March 2021 onwards culminating with the removal of most restrictions from 19 July 2021. We use as initial conditions the end conditions from left panels and $\mathcal{R}_c(t) = \mathcal{R}_0$. This is done without vaccination (heavy black) and with vaccination (effective protection of 8% per month; heavy red). We explore additional vaccination scenarios (thin curves), from top to bottom (in % of susceptible population per month): 1, 2, 3, 4, 5, 6, 7 (black); 9, 10, 11, 12, 13, 14, 15, 16 (red). In the main text (Section 5.6) we implement a more realistic vaccination programme.

The reduced models used in the main text are convenient for mathematical, numerical and statistical analyses given their low dimension but have little flexibility for extensions, such as including reinfection, vaccination or viral evolution. However, the agreement of the results presented here with those in Section 5.1 is reassuring that we can use one or the other formalism interchangeably, especially in the case of variable susceptibility where model reduction is exact (approximate in the case of variable connectivity). The realistic vaccination programme simulated in Section 5.6 was implemented by inputing parameters estimated with the reduced model (Section 5.1) in explicit distribution formalism.

Table S1: Model parameters estimated by Bayesian inference based on daily deaths until 1 February 2021 (with explicit gamma distributed traits). Model selection based on maximum log-likelihood (LL) and Akaike information criterion (AIC). Best fitting models have lower AIC scores (bold). Infection fatality ratio, $\phi = 0.9\%$. Herd immunity threshold ($\mathcal{H}$) calculated from $\mathcal{R}_0$ and $\nu$ using formulas in (S7) or (S8) as appropriate. $T_0$ and $T_2$ parameterise linear reduction and increase in transmissibility, respectively, before and after first lockdown (larger $T \Leftrightarrow$ lower slope; main text and Fig. S13).

| | Heterogeneous susceptibility | | Heterogeneous connectivity | | Homogeneous | |
|---|---|---|---|---|---|---|
| | Median | 95% CI | Median | 95% CI | Median | 95% CI |
| *Common parameters* | | | | | | |
| $c_1$ [a] | 0.2806 | $(0.2795, 0.2816)$ | 0.2386 | $(0.2355, 0.2411)$ | 0.2751 | $(0.2740, 0.2772)$ |
| $c_2$ [b] | 0.6637 | $(0.6608, 0.6655)$ | 0.6479 | $(0.6415, 0.6590)$ | 0.4803 | $(0.4776, 0.4869)$ |
| $\eta$ [c] | 11 | $\{11\}$ | 12 | $\{12\}$ | 9 | $\{9, 10\}$ |
| $\nu$ [d] | 1.638 | $(1.627, 1.648)$ | 1.227 | $(1.224, 1.236)$ | 0 | – |
| *England* | | | | | | |
| $T_0$ | −0.5678 | $(-0.7241, -0.4243)$ | −3.321 | $(-3.689, -2.813)$ | 7.595 | $(6.955, 7.771)$ |
| $T_2$ | 325.5 | $(323.3, 327.4)$ | 381.2 | $(373.2, 386.7)$ | 416.1 | $(408.9, 418.3)$ |
| $\mathcal{R}_0$ | 3.001 | $(2.991, 3.011)$ | 3.545 | $(3.513, 3.586)$ | 2.691 | $(2.683, 2.701)$ |
| $\mathcal{H}$ | 25.80% | $(25.53\%, 26.09\%)$ | 27.05% | $(26.66\%, 27.34\%)$ | 62.84% | $(62.72\%, 62.98\%)$ |
| *Scotland* | | | | | | |
| $T_0$ | 3.467 | $(3.149, 3.913)$ | 3.790 | $(2.818, 4.394)$ | 11.78 | $(11.36, 12.58)$ |
| $T_2$ | 463.6 | $(456.6, 467.2)$ | 458.1 | $(448.9, 471.2)$ | 540.0 | $(524.9, 546.9)$ |
| $\mathcal{R}_0$ | 3.242 | $(3.228, 3.255)$ | 3.718 | $(3.680, 3.783)$ | 2.978 | $(2.954, 2.995)$ |
| $\mathcal{H}$ | 27.34% | $(27.04\%, 27.65\%)$ | 27.91% | $(27.49\%, 28.31\%)$ | 66.41% | $(66.14\%, 66.61\%)$ |
| *Model selection* | | | | | | |
| LL | −3745 | | −3494 | | −5893 | |
| AIC | 7509 | | **7008** | | 11805 | |

[a]transmissibility reduction due to lockdown 1,
[b]transmissibility reduction due to lockdowns 2 and 3,
[c]difference between mean-time-to-death and mean-time-to-recovery (sampled from a continuous interval and reduced to the nearest integer before entering the model),
[d]coefficient of variation (CV).

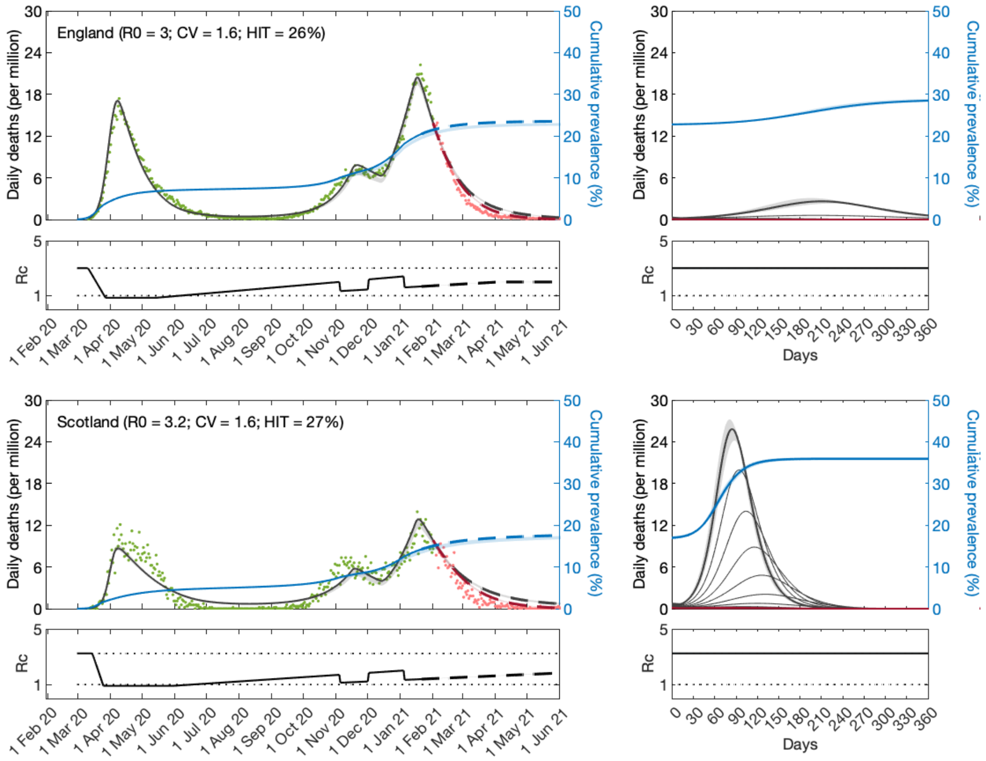


Figure S1: **SARS-CoV-2 transmission in England and Scotland with individual variation in susceptibility to infection.** Susceptibility factors implemented explicitly as gamma distributions (model (S1)-(S4) with force of infection (S5)). Modelled trajectories of COVID-19 deaths (black and red curves) and cumulative percentage infected (blue). Dots are data for daily reported deaths: fitted (green); out-of-sample (red). Basic reproduction numbers under control ($\mathcal{R}_c$) displayed in shallow panels underneath the main plots. Left panels represent fitted segments as solid curves and projected scenarios as dashed: without vaccination (black); with a vaccination programme that effectively immunises 8% of the susceptible population per month from February 2121 onwards (red). Right panels prolong those projections further in time assuming $\mathcal{R}_c(t) = \mathcal{R}_0$ (heavier curves) and explore additional vaccination scenarios (thin curves), from top to bottom (in % of susceptible population per month): 1, 2, 3, 4, 5, 6, 7 (black); 9, 10, 11, 12, 13, 14, 15, 16 (red). Input parameters: progression from $E$ to $I$ ($\delta = 1/5.5$ per day); recovery ($\gamma = 1/4$ per day); relative infectiousness between $E$ and $I$ stages ($\rho = 0.5$); and IFR ($\phi = 0.9\%$). Initial basic reproduction numbers, coefficients of variation and control parameters estimated by Bayesian inference (estimates in Table S1). Fitted curves represent best fitting trajectories and shades are 95% credible intervals generated from 10,000 posterior samples.

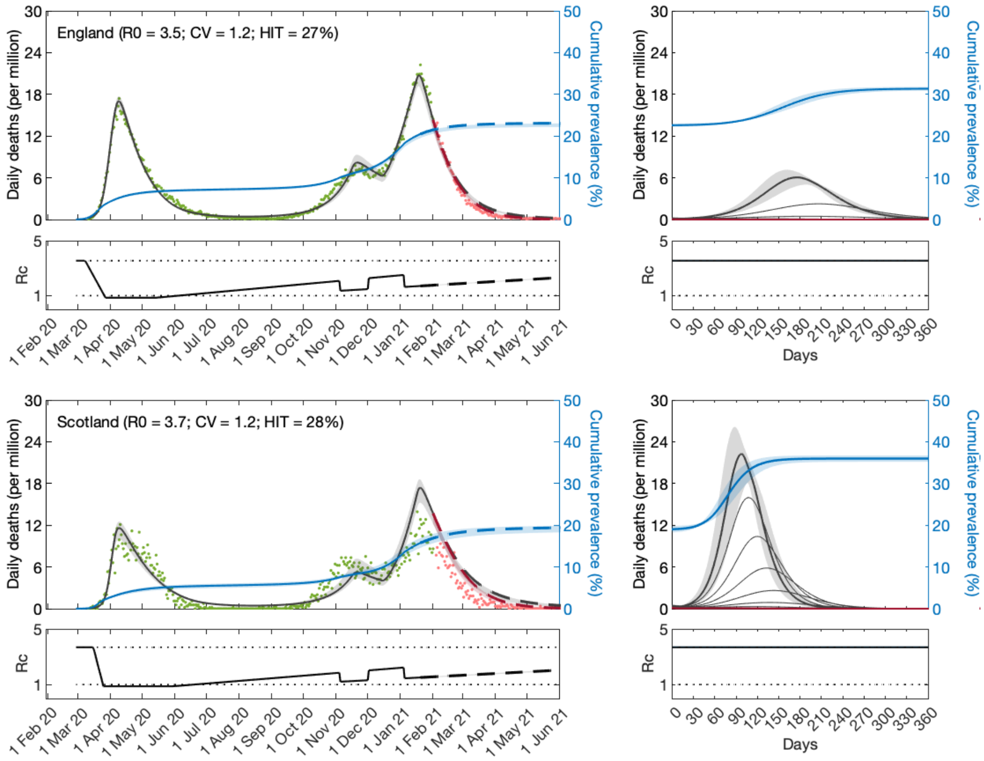


Figure S2: **SARS-CoV-2 transmission in England and Scotland with individual variation in exposure to infection.** Connectivity factors implemented explicitly as gamma distributions (model (S1)-(S4) with force of infection (S6)). Modelled trajectories of COVID-19 deaths (black and red curves) and cumulative percentage infected (blue). Dots are data for daily reported deaths: fitted (green); out-of-sample (red). Basic reproduction numbers under control ($\mathcal{R}_c$) displayed in shallow panels underneath the main plots. Left panels represent fitted segments as solid curves and projected scenarios as dashed: without vaccination (black); with a vaccination programme that effectively immunises 8% of the susceptible population per month from February 2121 onwards (red). Right panels prolong those projections further in time assuming $\mathcal{R}_c(t) = \mathcal{R}_0$ (heavier curves) and explore additional vaccination scenarios (thin curves), from top to bottom (in % of susceptible population per month): 1, 2, 3, 4, 5, 6, 7 (black); 9, 10, 11, 12, 13, 14, 15, 16 (red). Input parameters: progression from $E$ to $I$ ($\delta = 1/5.5$ per day); recovery ($\gamma = 1/4$ per day); relative infectiousness between $E$ and $I$ stages ($\rho = 0.5$); and IFR ($\phi = 0.9\%$). Initial basic reproduction numbers, coefficients of variation and control parameters estimated by Bayesian inference (estimates in Table S1). Fitted curves represent best fitting trajectories and shades are 95% credible intervals generated from $10,000$ posterior samples.

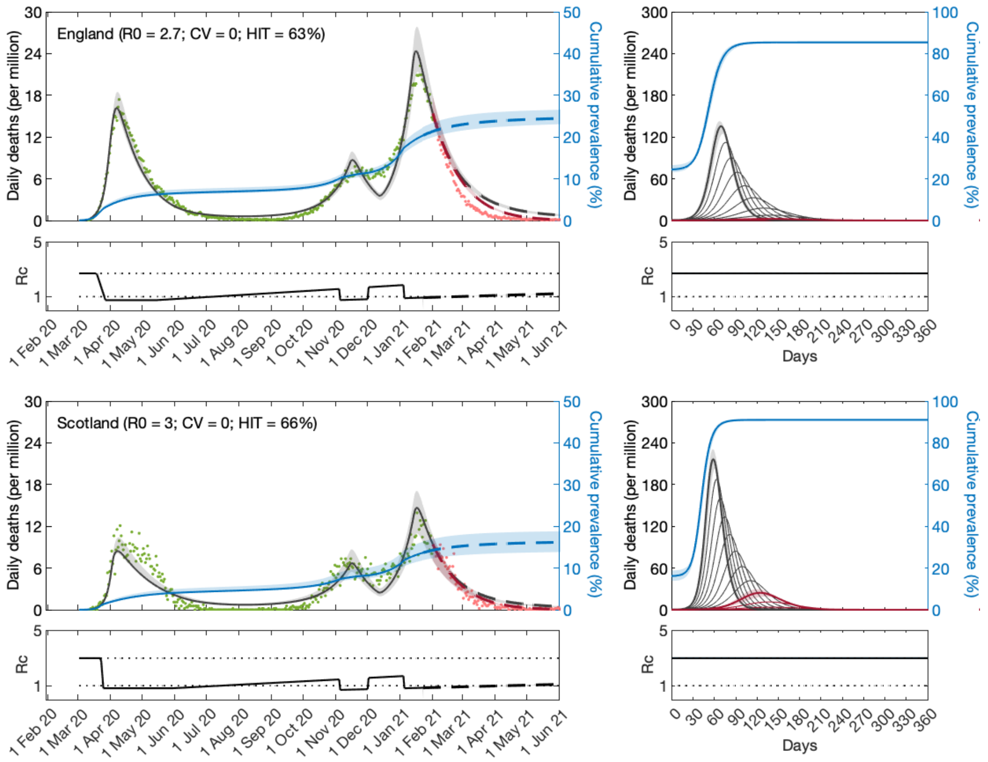


Figure S3: **SARS-CoV-2 transmission in England and Scotland assuming homogeneity.** Model (S1)-(S4) with force of infection (S6) or (S6) for $\nu = 0$. Modelled trajectories of COVID-19 deaths (black and red curves) and cumulative percentage infected (blue). Dots are data for daily reported deaths: fitted (green); out-of-sample (red). Basic reproduction numbers under control ($\mathcal{R}_c$) displayed in shallow panels underneath the main plots. Left panels represent fitted segments as solid curves and projected scenarios as dashed: without vaccination (black); with a vaccination programme that effectively immunises 8% of the susceptible population per month from February 2121 onwards (red). Right panels prolong those projections further in time assuming $\mathcal{R}_c(t) = \mathcal{R}_0$ (heavier curves) and explore additional vaccination scenarios (thin curves), from top to bottom (in % of susceptible population per month): 1, 2, 3, 4, 5, 6, 7 (black); 9, 10, 11, 12, 13, 14, 15, 16 (red). Input parameters: progression from $E$ to $I$ ($\delta = 1/5.5$ per day); recovery ($\gamma = 1/4$ per day); relative infectiousness between $E$ and $I$ stages ($\rho = 0.5$); and IFR ($\phi = 0.9\%$). Initial basic reproduction numbers and control parameters estimated by Bayesian inference (estimates in Table S1). Fitted curves represent best fitting trajectories and shades are 95% credible intervals generated from 10,000 posterior samples.

#### S2 Sensitivity to reinfection

[Montalbán *et al.* 2020] extended and analysed system (S1)-(S4) to include unlimited repeated infections:

$$\frac{dS(x)}{dt} = -\lambda \; x \; S(x), \tag{S9}$$

$$\frac{dE(x)}{dt} = \lambda \; x \; (S(x) + \sigma \; R(x)) - \delta \; E(x), \tag{S10}$$

$$\frac{dI(x)}{dt} = \delta \; E(x) - \gamma \; I(x), \tag{S11}$$

$$\frac{dR(x)}{dt} = (1-\phi) \; \gamma \; I(x) - \sigma \; \lambda \; x \; R(x), \tag{S12}$$

with force of infection written again given by (S5) for variable susceptibility and (S6) for variable connectivity. Infection history parameters $\delta$ and $\gamma$ are as in Section S1, and $\sigma$ is the risk of reinfection relative to that of first infection. Basic reproduction numbers are given by the same expressions as in the absence of reinfection (Section S1) while herd immunity thresholds are reformulated to account for reinfection. [Montalbán *et al.* 2020] derive expressions for the herd immunity threshold when the reinfection factor is below the reinfection threshold (i.e., when $\sigma > 1/\mathcal{R}_0$ [Gomes *et al.* 2004, Gomes *et al.* 2016]), specifically,

$$\mathcal{H}_\sigma = 1 - \left( \frac{1 - \sigma \mathcal{R}_0}{(1-\sigma)\mathcal{R}_0} \right)^{\frac{1}{1+\nu^2}} \tag{S13}$$

for variable susceptibility, and

$$\mathcal{H}_\sigma = 1 - \left( \frac{1 - \sigma \mathcal{R}_0}{(1-\sigma)\mathcal{R}_0} \right)^{\frac{1}{1+2\nu^2}} \tag{S14}$$

for variable connectivity. The authors also show that system (S1)-(S4) has no herd immunity threshold when above the reinfection threshold, with the entire population expected to be infected repeatedly in that scenario (i.e., $\mathcal{H}_\sigma = 1$).

Here we consider an intermediate version which contemplates the possibility of a second infection but no further reinfections within the study period (between 1 and 2 years):

$$\frac{dS(x)}{dt} = -\lambda \; x \; S(x), \tag{S15}$$

$$\frac{dE(x)}{dt} = \lambda \; x \; S(x) - \delta \; E(x), \tag{S16}$$

$$\frac{dI(x)}{dt} = \delta \; E(x) - \gamma \; I(x), \tag{S17}$$

$$\frac{dR(x)}{dt} = (1-\phi) \; \gamma \; I(x) - \sigma \; \lambda \; x \; R(x), \tag{S18}$$

$$\frac{dE'(x)}{dt} = \sigma \; \lambda \; x \; R(x) - \delta \; E'(x), \tag{S19}$$

$$\frac{dI'(x)}{dt} = \delta \; E'(x) - \gamma \; I'(x), \tag{S20}$$

$$\frac{dR'(x)}{dt} = (1-\phi) \; \gamma \; I'(x), \tag{S21}$$

with force of infection

$$\lambda \quad = \quad \frac{\beta}{\mathsf{N}} \int \left[\rho(E(x) + E'(x)) + I(x) + I'(x)\right] \, dx, \tag{S22}$$

when variation is in susceptibility, and

$$\lambda \quad = \quad \frac{\beta}{\mathsf{N}} \frac{\int x \left[\rho(E(x) + E'(x)) + I(x) + I'(x)\right] \, dx}{\int xq(x) \, dx}, \tag{S23}$$

when it is connectivity that varies between individuals. Infection history parameters $\delta$ and $\gamma$ are as above and $\sigma$ is the relative risk of second infection relative to the first. We have no expression for the herd immunity threshold in this case but know it lies between $\mathcal{H}$ and $\mathcal{H}_\sigma$.

For concreteness, in the analyses conducted in this section we set $\sigma = 0.1$ and assume that reinfection does not lead to death. We adopt a slightly conservative estimate for $\sigma$ in relation to, for example [Hall *et al.* 2021] (0.16 [95% CI, 0.13-0.19]), because direct measurements are likely to be amplified by the same process of selection on individual variation that is the focus of this work (see [Rodrigues *et al.* 2016], for instance). Studies like [Hall *et al.* 2021] measure relative risk of reinfection with reference to a susceptible pool that has been depleted of highly susceptible individuals, which is higher than relative risk of reinfection with reference to the whole population ($\sigma$ in our model).

Results are shown in Fig. S4 for individual variation in susceptibility and Fig. S5 for individual variation in connectivity. The homogeneous model is obtained by setting $\nu = 0$ in either model (Fig. S6). Estimated parameters are provided (with 95% credible intervals) in Table S2.

In agreement with the scenario $\sigma = 0$ presented in the main text (Section 5.1) and in Section S1, we find that heterogeneous models are significantly better supported by the data (lower AIC scores). These models also indicate substantially lower HITs $((\mathcal{H}, \mathcal{H}_\sigma)$ around $(22-24\%, 25-30\%)$ than homogeneous $(62-67\%, 69-75\%)$. Finally, when epidemic trajectories are projected beyond the fitted period we find similar dynamics (compare the right panels in Figs, S4, S5 and S6, with those in main text Figs. 3, 4 and 5 (or Figs. S1, S2 and S3), respectively).

Table S2: Model parameters estimated by Bayesian inference based on daily deaths until 1 February 2021 (with reinfection). Model selection based on maximum log-likelihood (LL) and Akaike information criterion (AIC). Best fitting models have lower AIC scores (bold). Infection fatality ratio, $\phi = 0.9\%$, and reinfection factor, $\sigma = 0.1$. Lower bound for herd immunity threshold ($\mathcal{H}$) calculated from $\mathcal{R}_0$ and $\nu$ using formulas in (S7) or (S8) as appropriate. Upper bound ($\mathcal{H}_\sigma$) calculated using formulas (S13) or (S14). $T_0$ and $T_2$ parameterise linear reduction and increase in transmissibility, respectively, before and after first lockdown (larger $T \Leftrightarrow$ lower slope; main text and Fig. S13).

| | Heterogeneous susceptibility | | Heterogeneous connectivity | | Homogeneous | |
|---|---|---|---|---|---|---|
| | Median | 95% CI | Median | 95% CI | Median | 95% CI |
| *Common parameters* | | | | | | |
| $c_1$ [a] | 0.3250 | $(0.3231, 0.3272)$ | 0.2484 | $(0.2474, 0.2507)$ | 0.2751 | $(0.2736, 0.2777)$ |
| $c_2$ [b] | 0.5916 | $(0.5877, 0.6006)$ | 0.6916 | $(0.6830, 0.6956)$ | 0.5029 | $(0.4996, 0.5086)$ |
| $\eta$ [c] | 13 | $\{13\}$ | 11 | $\{11\}$ | 8 | $\{8\}$ |
| $\nu$ [d] | 1.615 | $(1.610, 1.637)$ | 1.381 | $(1.365, 1.389)$ | 0 | – |
| *England* | | | | | | |
| $T_0$ | 8.695 | $(8.163, 9.012)$ | 1.180 | $(1.006, 1.524)$ | 12.27 | $(11.89, 12.44)$ |
| $T_2$ | 243.3 | $(242.7, 245.0)$ | 362.6 | $(361.6, 364.1)$ | 402.7 | $(401.5, 406.2)$ |
| $\mathcal{R}_0$ | 2.462 | $(2.453, 2.482)$ | 3.478 | $(3.433, 3.490)$ | 2.629 | $(2.620, 2.637)$ |
| $\mathcal{H}$ | 22.11% | $(21.65\%, 22.35\%)$ | 22.80% | $(22.42\%, 23.23\%)$ | 61.96% | $(61.83\%, 62.08\%)$ |
| $\mathcal{H}_\sigma$ | 25.83% | $(25.30\%, 26.15\%)$ | 27.80% | $(27.29\%, 28.32\%)$ | 68.85% | $(68.70\%, 68.98\%)$ |
| *Scotland* | | | | | | |
| $T_0$ | 12.90 | $(12.34, 12.99)$ | 7.632 | $(7.169, 7.772)$ | 12.36 | $(11.74, 12.99)$ |
| $T_2$ | 333.7 | $(331.2, 338.4)$ | 457.6 | $(455.1, 463.2)$ | 589.1 | $(572.5, 594.2)$ |
| $\mathcal{R}_0$ | 2.702 | $(2.685, 2.713)$ | 3.644 | $(3.629, 3.672)$ | 3.020 | $(2.996, 3.035)$ |
| $\mathcal{H}$ | 24.08% | $(23.55\%, 24.25\%)$ | 23.55% | $(23.30\%, 24.06\%)$ | 66.88% | $(66.62\%, 67.06\%)$ |
| $\mathcal{H}_\sigma$ | 28.36% | $(27.73\%, 28.58\%)$ | 28.88% | $(28.58\%, 29.51\%)$ | 74.32% | $(74.02\%, 74.50\%)$ |
| *Model selection* | | | | | | |
| LL | −3950 | | −3556 | | −5775 | |
| AIC | 7921 | | **7132** | | 11567 | |

[a]transmissibility reduction due to lockdown 1,
[b]transmissibility reduction due to lockdowns 2 and 3,
[c]difference between mean-time-to-death and mean-time-to-recovery (sampled from a continuous interval and reduced to the nearest integer before entering the model),
[d]coefficient of variation (CV).

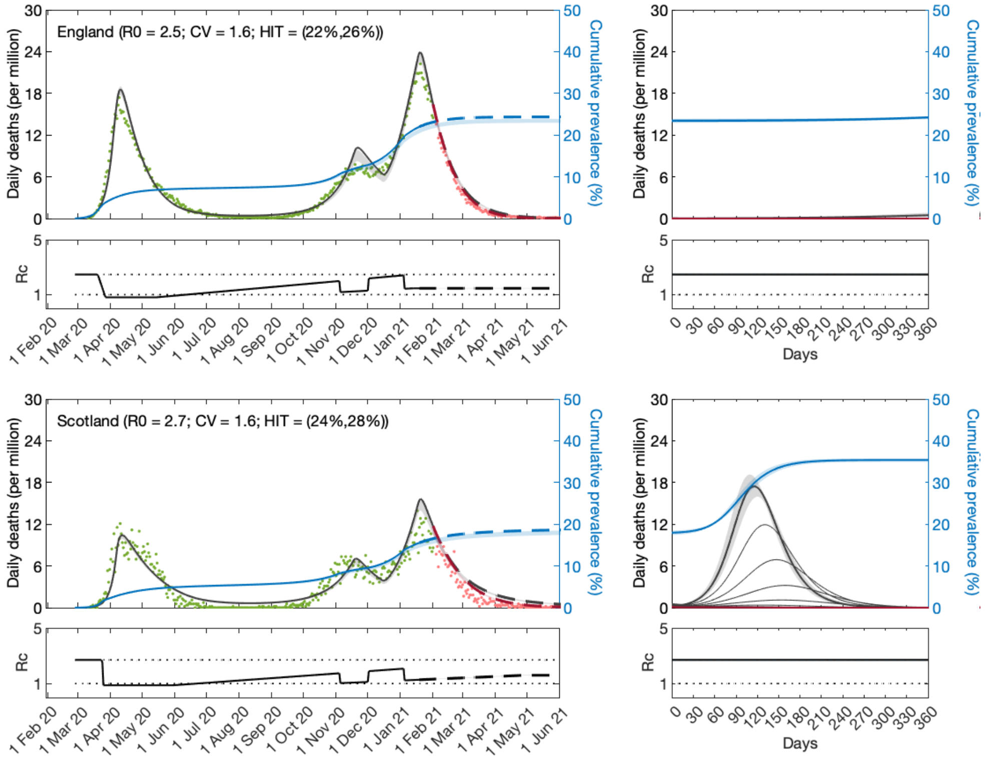


Figure S4: **SARS-CoV-2 transmission in England and Scotland with reinfection and individual variation in susceptibility infection.** Susceptibility factors implemented explicitly as gamma distributions (model (S15)-(S21) with force of infection (S22)). Modelled trajectories of COVID-19 deaths (black and red curves) and cumulative percentage infected (blue). Dots are data for daily reported deaths: fitted (green); out-of-sample (red). Basic reproduction numbers under control ($\mathcal{R}_c$) displayed in shallow panels underneath the main plots. Left panels represent fitted segments as solid curves and projected scenarios as dashed: without vaccination (black); with a vaccination programme that effectively immunises 8% of the susceptible population per month from February 2121 onwards (red). Right panels prolong those projections further in time assuming $\mathcal{R}_c(t) = \mathcal{R}_0$ (heavier curves) and explore additional vaccination scenarios (thin curves), from top to bottom (in % of susceptible population per month): 1, 2, 3, 4, 5, 6, 7 (black); 9, 10, 11, 12, 13, 14, 15, 16 (red). Input parameters: progression from $E$ to $I$ ($\delta = 1/5.5$ per day); recovery ($\gamma = 1/4$ per day); relative infectiousness between $E$ and $I$ stages ($\rho = 0.5$); IFR ($\phi = 0.9\%$); and reinfection factor ($\sigma = 0.1$). Initial basic reproduction numbers, coefficients of variation and control parameters estimated by Bayesian inference (estimates in Table S2). Fitted curves represent best fitting trajectories and shades are 95% credible intervals generated from $10,000$ posterior samples.

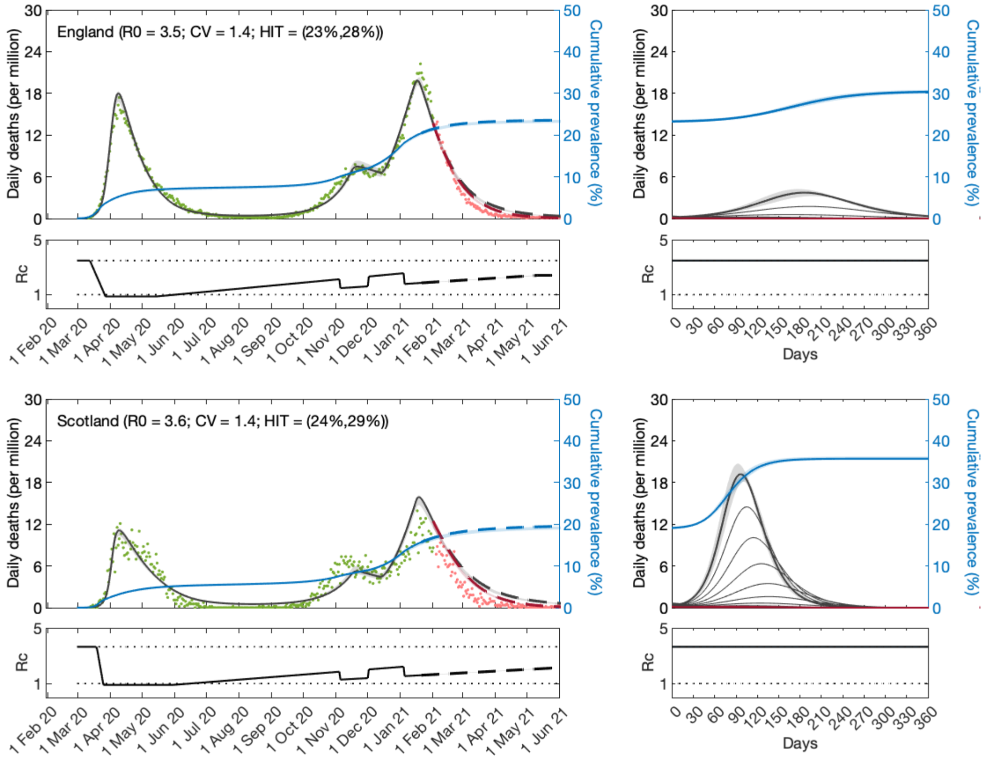


Figure S5: **SARS-CoV-2 transmission in England and Scotland with reinfection and individual variation in exposure to infection.** Connectivity factors implemented explicitly as gamma distributions (model (S15)-(S21) with force of infection (S23)). Modelled trajectories of COVID-19 deaths (black and red curves) and cumulative percentage infected (blue). Dots are data for daily reported deaths: fitted (green); out-of-sample (red). Basic reproduction numbers under control ($\mathcal{R}_c$) displayed in shallow panels underneath the main plots. Left panels represent fitted segments as solid curves and projected scenarios as dashed: without vaccination (black); with a vaccination programme that effectively immunises 8% of the susceptible population per month from February 2121 onwards (red). Right panels prolong those projections further in time assuming $\mathcal{R}_c(t) = \mathcal{R}_0$ (heavier curves) and explore additional vaccination scenarios (thin curves), from top to bottom (in % of susceptible population per month): 1, 2, 3, 4, 5, 6, 7 (black); 9, 10, 11, 12, 13, 14, 15, 16 (red). Input parameters: progression from $E$ to $I$ ($\delta = 1/5.5$ per day); recovery ($\gamma = 1/4$ per day); relative infectiousness between $E$ and $I$ stages ($\rho = 0.5$); IFR ($\phi = 0.9\%$); and reinfection factor ($\sigma = 0.1$). Initial basic reproduction numbers, coefficients of variation and control parameters estimated by Bayesian inference (estimates in Table S2). Fitted curves represent best fitting trajectories and shades are 95% credible intervals generated from $10,000$ posterior samples.

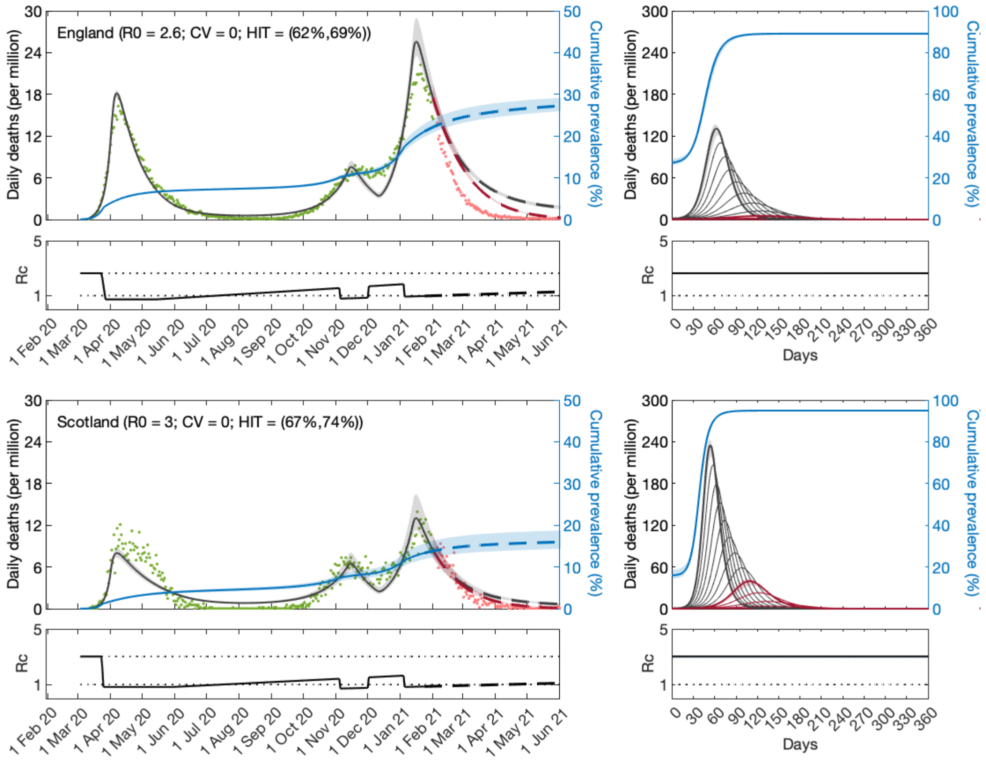


Figure S6: **SARS-CoV-2 transmission in England and Scotland with reinfection and homogeneity.** Model (S15)-(S21) with force of infection (S23) or (S23) for $\nu = 0$. Modelled trajectories of COVID-19 deaths (black and red curves) and cumulative percentage infected (blue). Dots are data for daily reported deaths: fitted (green); out-of-sample (red). Basic reproduction numbers under control ($\mathcal{R}_c$) displayed in shallow panels underneath the main plots. Left panels represent fitted segments as solid curves and projected scenarios as dashed: without vaccination (black); with a vaccination programme that effectively immunises 8% of the susceptible population per month from February 2121 onwards (red). Right panels prolong those projections further in time assuming $\mathcal{R}_c(t) = \mathcal{R}_0$ (heavier curves) and explore additional vaccination scenarios (thin curves), from top to bottom (in % of susceptible population per month): 1, 2, 3, 4, 5, 6, 7 (black); 9, 10, 11, 12, 13, 14, 15, 16 (red). Input parameters: progression from $E$ to $I$ ($\delta = 1/5.5$ per day); recovery ($\gamma = 1/4$ per day); relative infectiousness between $E$ and $I$ stages ($\rho = 0.5$); IFR ($\phi = 0.9\%$); and reinfection factor ($\sigma = 0.1$). Initial basic reproduction numbers and control parameters estimated by Bayesian inference (estimates in Table S2). Fitted curves represent best fitting trajectories and shades are 95% credible intervals generated from 10,000 posterior samples.

#### S3 Sensitivity to infection fatality ratio

The analyses in the main text assume infection fatality ratio (IFR) $\phi = 0.9\%$ [Ward *et al.* 2021, Chen *et al.* 2021]. Here we explore impacts of lower or higher IFR on estimated model parameters and projected epidemic dynamics. In this section we use the reduced models introduced in the main text: (9)-(12) for variable susceptibility; (17)-(20) for variable connectivity; and either formulation with $\nu = 0$ for homogeneity assumptions.

##### S3.1 IFR $= 0.7\%$

When IFR is assumed to take a lower value, each death represents more infections. Since we are estimating model parameters by fitting series of daily deaths, lower IFR is expected to result in higher HIT estimates. In this scenario we estimate $\mathcal{H}$ around $28 - 36\%$ with heterogeneous models and $69 - 70\%$ with homogeneous. Epidemics projected beyond the fitted period resemble those in the main analysis (compare the right panels in Figs, S7, S8 and S9, with those in main text Figs. 3, 4 and 5, respectively) but tend to be slightly smaller despite the higher HITs, illustrating the fragility of taking HITs out of context.

Table S3: Model parameters estimated by Bayesian inference based on daily deaths until 1 February 2021 (with lower IFR). Model selection based on maximum log-likelihood (LL) and Akaike information criterion (AIC). Best fitting models have lower AIC scores (bold). Infection fatality ratio, $\phi = 0.7\%$. Herd immunity threshold ($\mathcal{H}$) calculated from $\mathcal{R}_0$ and $\nu$ using main text formulas (14) or (22) as appropriate. $T_0$ and $T_2$ parameterise linear reduction and increase in transmissibility, respectively, before and after first lockdown (larger $T \Leftrightarrow$ lower slope; main text and Fig. S13).

| | Heterogeneous susceptibility | | Heterogeneous connectivity | | Homogeneous | |
|---|---|---|---|---|---|---|
| | Median | 95% CI | Median | 95% CI | Median | 95% CI |
| *Common parameters* | | | | | | |
| $c_1$ [a] | 0.2854 | $(0.2841, 0.2894)$ | 0.3122 | $(0.3097, 0.3130)$ | 0.2374 | $(0.2359, 0.2383)$ |
| $c_2$ [b] | 0.6515 | $(0.6413, 0.6602)$ | 0.7001 | $(0.6906, 0.7012)$ | 0.5689 | $(0.5677, 0.5708)$ |
| $\eta$ [c] | 13 | $\{13\}$ | 12 | $\{12\}$ | 9 | $\{9\}$ |
| $\nu$ [d] | 1.259 | $(1.227, 1.296)$ | 0.9935 | $(0.9866, 0.9979)$ | 0 | – |
| *England* | | | | | | |
| $T_0$ | $-3.563$ | $(-3.831, -3.144)$ | 3.267 | $(2.982, 3.325)$ | $-1.500$ | $(-1.655, -1.441)$ |
| $T_2$ | 322.0 | $(317.1, 325.2)$ | 269.6 | $(268.8, 270.8)$ | 544.1 | $(542.4, 547.1)$ |
| $\mathcal{R}_0$ | 2.931 | $(2.902, 2.946)$ | 2.719 | $(2.711, 2.735)$ | 3.261 | $(3.246, 3.269)$ |
| $\mathcal{H}$ | 34.04% | $(32.80\%, 35.03\%)$ | 28.56% | $(28.35\%, 28.93\%)$ | 69.33% | $(69.19\%, 69.41\%)$ |
| *Scotland* | | | | | | |
| $T_0$ | 7.418 | $(6.996, 7.828)$ | 7.133 | $(6.843, 7.486)$ | 8.262 | $(7.838, 9.040)$ |
| $T_2$ | 363.4 | $(356.7, 373.4)$ | 386.8 | $(383.7, 390.2)$ | 553.2 | $(549.3, 556.4)$ |
| $\mathcal{R}_0$ | 2.983 | $(2.962, 3.050)$ | 2.981 | $(2.961, 2.992)$ | 3.287 | $(3.278, 3.305)$ |
| $\mathcal{H}$ | 34.49% | $(33.32\%, 35.92\%)$ | 30.73% | $(30.43\%, 31.06\%)$ | 69.58% | $(69.50\%, 69.75\%)$ |
| *Model selection* | | | | | | |
| LL | $-3747$ | | $-3584$ | | $-5601$ | |
| AIC | 7514 | | **7188** | | 11219 | |

[a] transmissibility reduction due to lockdown 1,

[b] transmissibility reduction due to lockdowns 2 and 3,

[c] difference between mean-time-to-death and mean-time-to-recovery (sampled from a continuous interval and reduced to the nearest integer before entering the model),

[d] coefficient of variation (CV).

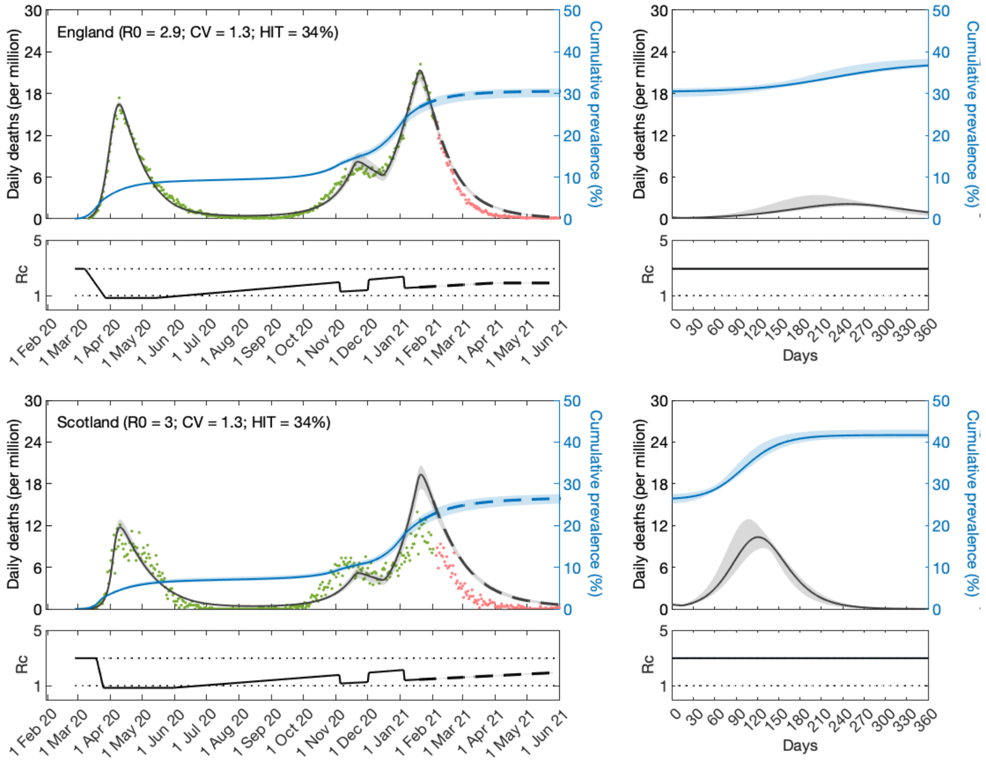


Figure S7: **SARS-CoV-2 transmission in England and Scotland with individual variation in susceptibility to infection and $\phi = 0.7\%$ (IFR).** Susceptibility factors implemented as gamma distributions (reduced model as in main text (9)-(12)). Modelled trajectories of COVID-19 deaths (black) and cumulative percentage infected (blue). Dots are data for daily reported deaths: fitted (green); out-of-sample (red). Basic reproduction numbers under control ($\mathcal{R}_c$) displayed in shallow panels underneath the main plots. Left panels represent fitted segments as solid curves and projected scenarios as dashed. Right panels prolong those projections further in time assuming $\mathcal{R}_c(t) = \mathcal{R}_0$. Input parameters: progression from $E$ to $I$ ($\delta = 1/5.5$ per day); recovery ($\gamma = 1/4$ per day); and relative infectiousness between $E$ and $I$ stages ($\rho = 0.5$). Initial basic reproduction numbers, coefficients of variation and control parameters estimated by Bayesian inference (Table S3). Fitted curves represent best fitting trajectories and shades are 95% credible intervals generated from 10,000 posterior samples.

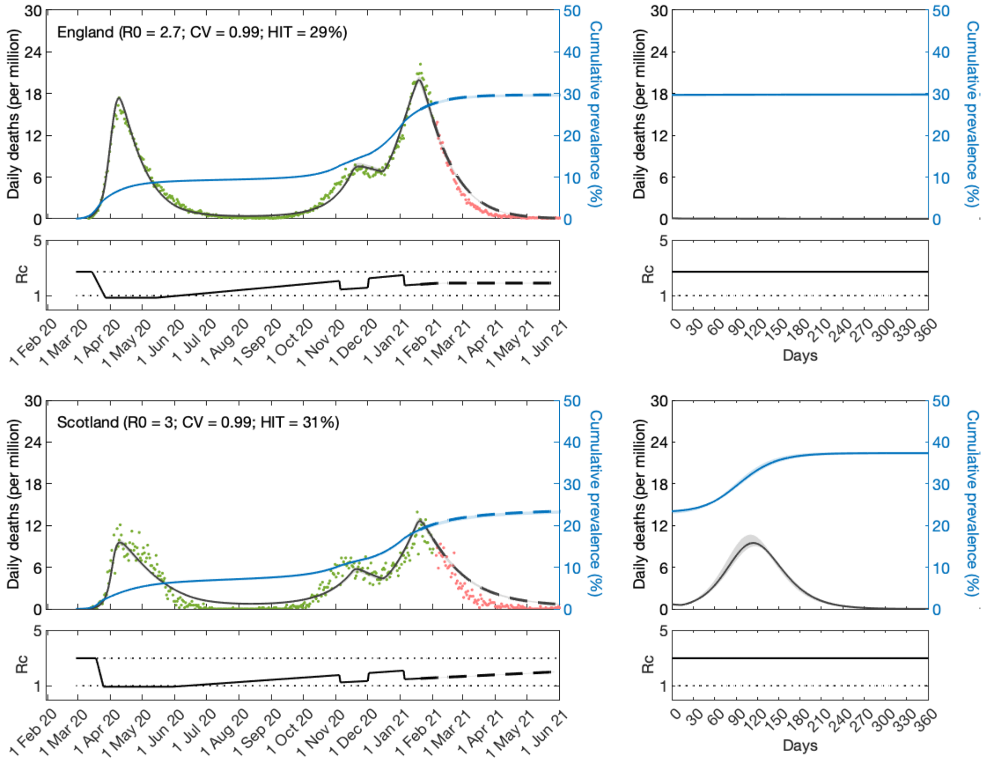


Figure S8: **SARS-CoV-2 transmission in England and Scotland with individual variation in exposure to infection and $\phi = 0.7\%$ (IFR).** Connectivity factors implemented as gamma distributions (reduced model as in main text (17)-(20)). Modelled trajectories of COVID-19 deaths (black) and cumulative percentage infected (blue). Dots are data for daily reported deaths: fitted (green); out-of-sample (red). Basic reproduction numbers under control ($\mathcal{R}_c$) displayed in shallow panels underneath the main plots. Left panels represent fitted segments as solid curves and projected scenarios as dashed. Right panels prolong those projections further in time assuming $\mathcal{R}_c(t) = \mathcal{R}_0$. Input parameters: progression from $E$ to $I$ ($\delta = 1/5.5$ per day); recovery ($\gamma = 1/4$ per day); and relative infectiousness between $E$ and $I$ stages ($\rho = 0.5$). Initial basic reproduction numbers, coefficients of variation and control parameters estimated by Bayesian inference (Table S3). Fitted curves represent best fitting trajectories and shades are 95% credible intervals generated from $10,000$ posterior samples.

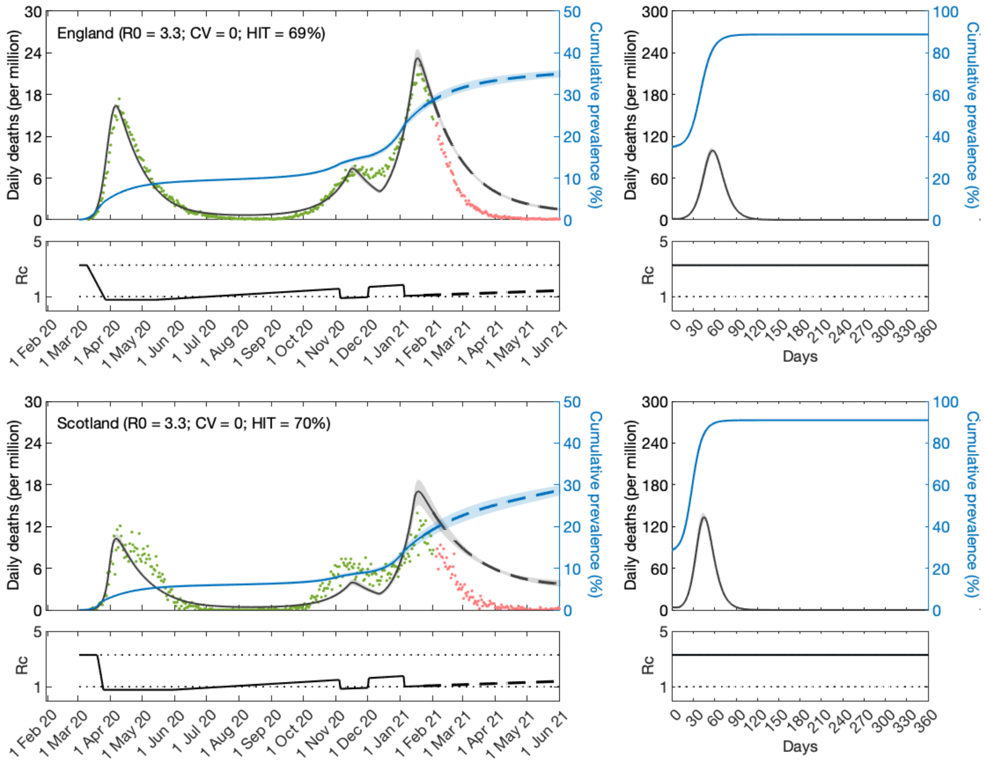


Figure S9: **SARS-CoV-2 transmission in England and Scotland with homogeneity and $\phi = 0.7\%$ (IFR).** Reduced model as in main text (7)-(12) or (17)-(20) with $\nu = 0$. Modelled trajectories of COVID-19 deaths (black) and cumulative percentage infected (blue). Dots are data for daily reported deaths: fitted (green); out-of-sample (red). Basic reproduction numbers under control ($\mathcal{R}_c$) displayed in shallow panels underneath the main plots. Left panels represent fitted segments as solid curves and projected scenarios as dashed. Right panels prolong those projections further in time assuming $\mathcal{R}_c(t) = \mathcal{R}_0$. Input parameters: progression from $E$ to $I$ ($\delta = 1/5.5$ per day); recovery ($\gamma = 1/4$ per day); and relative infectiousness between $E$ and $I$ stages ($\rho = 0.5$). Initial basic reproduction numbers and control parameters estimated by Bayesian inference (Table S3). Fitted curves represent best fitting trajectories and shades are 95% credible intervals generated from 10,000 posterior samples.

#### S3.2 IFR = 1.0%

Higher IFR is expected to result in lower HIT estimates. In this scenario we estimate $\mathcal{H}$ around 22 − 27% with heterogeneous models and 62 − 66% with homogeneous. Epidemics projected beyond the fitted period resemble those in the main analysis (compare the right panels in Figs, S10, S11 and S12, with those in main text Figs. 3, 4 and 5, respectively) but tend to be slightly larger despite the lower HITs, reinforcing again the fragility of taking HITs out of context.

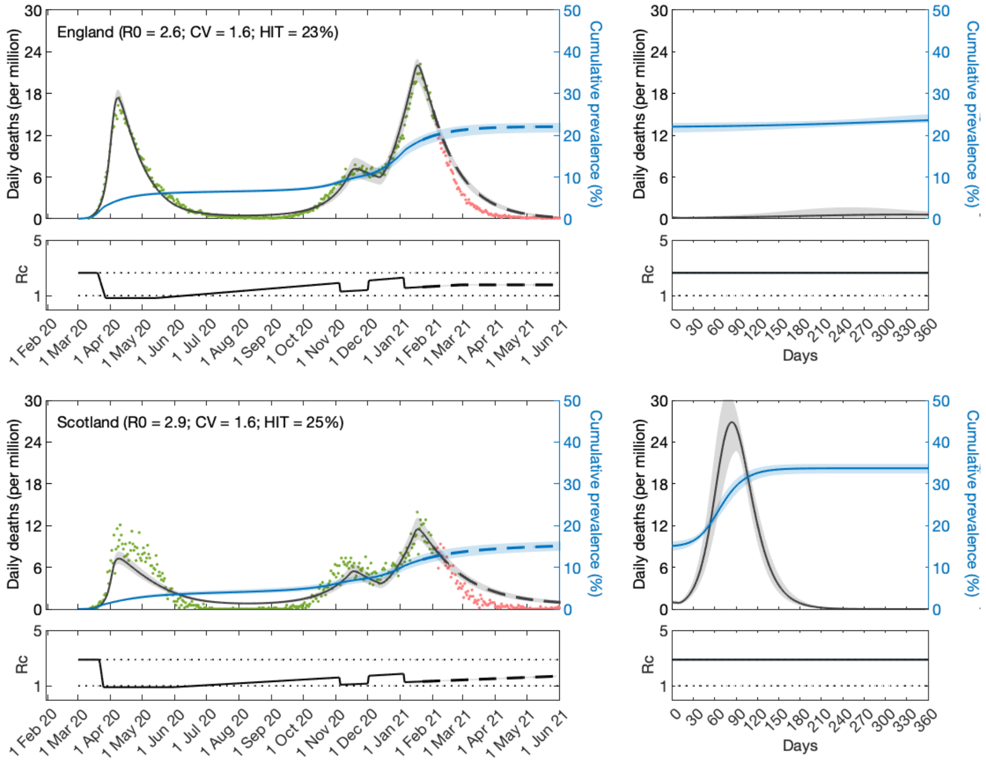


Figure S10: **SARS-CoV-2 transmission in England and Scotland with individual variation in susceptibility to infection and $\phi = 1.0\%$ (IFR).** Susceptibility factors implemented as gamma distributions (reduced model as in main text (9)-(12)). Modelled trajectories of COVID-19 deaths (black) and cumulative percentage infected (blue). Dots are data for daily reported deaths: fitted (green); out-of-sample (red). Basic reproduction numbers under control ($\mathcal{R}_c$) displayed in shallow panels underneath the main plots. Left panels represent fitted segments as solid curves and projected scenarios as dashed. Right panels prolong those projections further in time assuming $\mathcal{R}_c(t) = \mathcal{R}_0$. Input parameters: progression from $E$ to $I$ ($\delta = 1/5.5$ per day); recovery ($\gamma = 1/4$ per day); and relative infectiousness between $E$ and $I$ stages ($\rho = 0.5$). Initial basic reproduction numbers, coefficients of variation and control parameters estimated by Bayesian inference (Table S4). Fitted curves represent best fitting trajectories and shades are 95% credible intervals generated from 10,000 posterior samples.

Table S4: Model parameters estimated by Bayesian inference based on daily deaths until 1 February 2021 (with higher IFR). Model selection based on maximum log-likelihood (LL) and Akaike information criterion (AIC). Best fitting models have lower AIC scores (bold). Infection fatality ratio, $\phi = 1.0\%$. Herd immunity threshold ($\mathcal{H}$) calculated from $\mathcal{R}_0$ and $\nu$ using main text formulas (14) or (22) as appropriate. $T_0$ and $T_2$ parameterise linear reduction and increase in transmissibility, respectively, before and after first lockdown (larger $T \Leftrightarrow$ lower slope; main text and Fig. S13).

| | Heterogeneous susceptibility | | Heterogeneous connectivity | | Homogeneous | |
|---|---|---|---|---|---|---|
| | Median | 95% CI | Median | 95% CI | Median | 95% CI |
| *Common parameters* | | | | | | |
| $c_1$ [a] | 0.3142 | $(0.308, 0.3162)$ | 0.3168 | $(0.3150, 0.3182)$ | 0.2780 | $(0.2772, 0.2787)$ |
| $c_2$ [b] | 0.6735 | $(0.6538, 0.6867)$ | 0.6781 | $(0.6699, 0.6796)$ | 0.5072 | $(0.5027, 0.5083)$ |
| $\eta$ [c] | 10 | $\{10\}$ | 10 | $\{10\}$ | 9 | $\{9\}$ |
| $\nu$ [d] | 1.627 | $(1.583, 1.681)$ | 1.175 | $(1.151, 1.188)$ | 0 | – |
| *England* | | | | | | |
| $T_0$ | 8.625 | $(8.222, 8.912)$ | 8.882 | $(8.683, 9.000)$ | 8.974 | $(8.722, 9.001)$ |
| $T_2$ | 289.9 | $(283.2, 295.5)$ | 284.4 | $(279.4, 288.2)$ | 416.9 | $(415.6, 418.8)$ |
| $\mathcal{R}_0$ | 2.638 | $(2.627, 2.680)$ | 2.634 | $(2.619, 2.645)$ | 2.650 | $(2.639, 2.657)$ |
| $\mathcal{H}$ | 23.36% | $(22.31\%, 24.50\%)$ | 22.70% | $(22.26\%, 23.41\%)$ | 62.26% | $(62.10\%, 62.36\%)$ |
| *Scotland* | | | | | | |
| $T_0$ | 10.22 | $(9.238, 10.93)$ | 11.02 | $(10.48, 12.20)$ | 12.73 | $(12.43, 12.99)$ |
| $T_2$ | 449.1 | $(435.1, 459.9)$ | 434.5 | $(412.0, 442.2)$ | 543.4 | $(539.7, 552.0)$ |
| $\mathcal{R}_0$ | 2.885 | $(2.849, 2.953)$ | 2.888 | $(2.842, 2.905)$ | 2.947 | $(2.938, 2.956)$ |
| $\mathcal{H}$ | 25.22% | $(23.94\%, 26.56\%)$ | 24.57% | $(23.91\%, 25.35\%)$ | 66.07% | $(65.96\%, 66.17\%)$ |
| *Model selection* | | | | | | |
| LL | −3937 | | −3818 | | −5893 | |
| AIC | 7893 | | **7655** | | 11805 | |

[a] transmissibility reduction due to lockdown 1,
[b] transmissibility reduction due to lockdowns 2 and 3,
[c] difference between mean-time-to-death and mean-time-to-recovery (sampled from a continuous interval and reduced to the nearest integer before entering the model),
[d] coefficient of variation (CV).

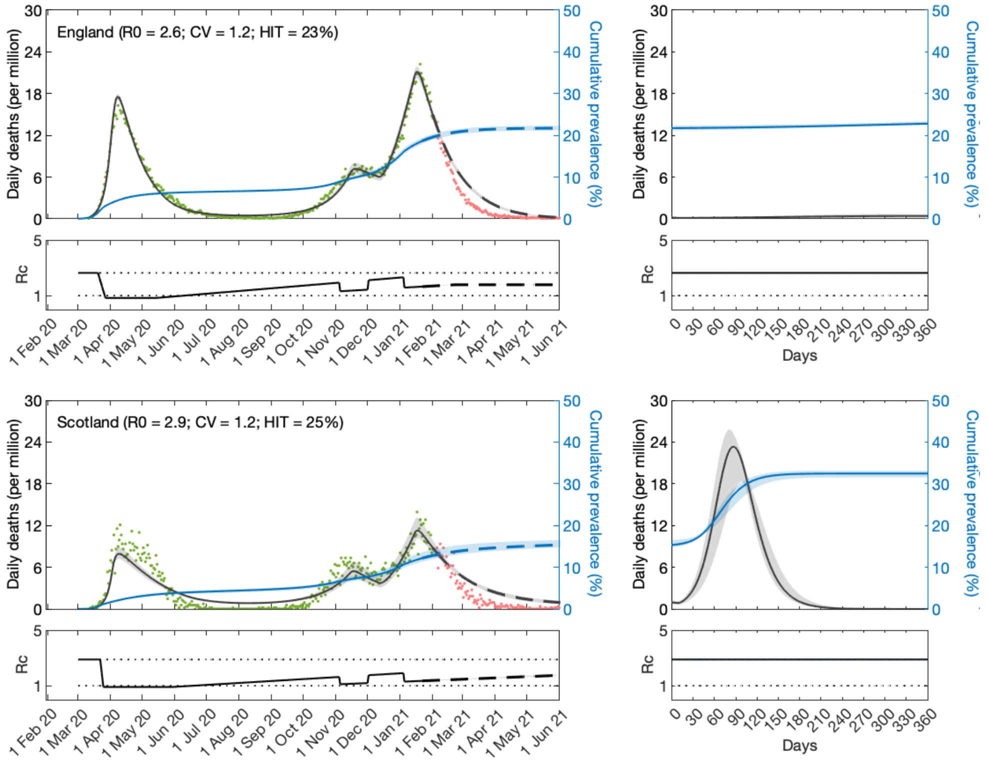


Figure S11: **SARS-CoV-2 transmission in England and Scotland with individual variation in exposure to infection and $\phi = 1.0\%$ (IFR).** Connectivity factors implemented as gamma distributions (reduced model as in main text (17)-(20)). Modelled trajectories of COVID-19 deaths (black) and cumulative percentage infected (blue). Dots are data for daily reported deaths: fitted (green); out-of-sample (red). Basic reproduction numbers under control ($\mathcal{R}_c$) displayed in shallow panels underneath the main plots. Left panels represent fitted segments as solid curves and projected scenarios as dashed. Right panels prolong those projections further in time assuming $\mathcal{R}_c(t) = \mathcal{R}_0$. Input parameters: progression from $E$ to $I$ ($\delta = 1/5.5$ per day); recovery ($\gamma = 1/4$ per day); and relative infectiousness between $E$ and $I$ stages ($\rho = 0.5$). Initial basic reproduction numbers, coefficients of variation and control parameters estimated by Bayesian inference (Table S4). Fitted curves represent best fitting trajectories and shades are 95% credible intervals generated from 10,000 posterior samples.

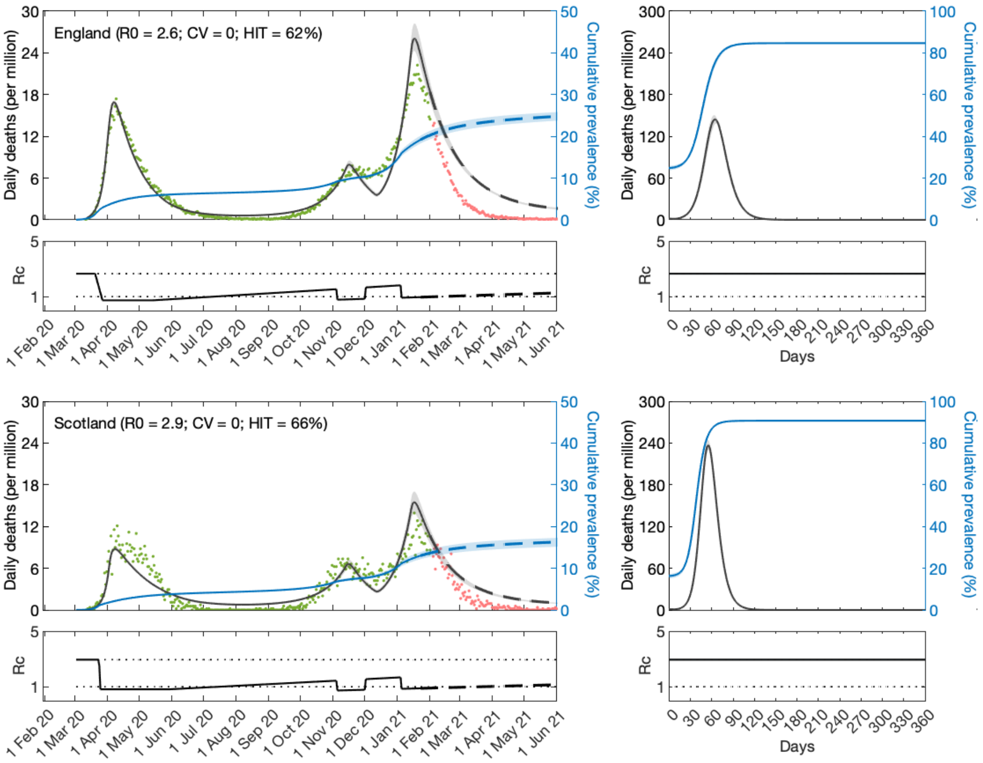


Figure S12: **SARS-CoV-2 transmission in England and Scotland with homogeneity and $\phi = 1.0\%$ (IFR).** Reduced model as in main text (7)-(12) or (17)-(20) with $\nu = 0$. Modelled trajectories of COVID-19 deaths (black) and cumulative percentage infected (blue). Dots are data for daily reported deaths: fitted (green); out-of-sample (red). Basic reproduction numbers under control ($\mathcal{R}_c$) displayed in shallow panels underneath the main plots. Left panels represent fitted segments as solid curves and projected scenarios as dashed. Right panels prolong those projections further in time assuming $\mathcal{R}_c(t) = \mathcal{R}_0$. Input parameters: progression from $E$ to $I$ ($\delta = 1/5.5$ per day); recovery ($\gamma = 1/4$ per day); and relative infectiousness between $E$ and $I$ stages ($\rho = 0.5$). Initial basic reproduction numbers and control parameters estimated by Bayesian inference (Table S4). Fitted curves represent best fitting trajectories and shades are 95% credible intervals generated from 10,000 posterior samples.

#### S4 Preliminary analysis of the first wave

Fitting the models to the first pandemic wave only, requires less constraints on the factor $c(t)$ that modifies the basic reproduction number relative to the original $\mathcal{R}_0$. Here we present an exploratory analysis of daily COVID-19 deaths in England and Scotland until 1 July 2020. We fitted the data with a shorter version of $c(t)$ introduced in the main text. Given the shorter timespan, autumn and winter constraints due to lockdowns 2 and 3 do not influence the results in this case. The $c(t)$ profile adopted here is depicted in Fig. S13.

Model fittings are shown in Fig. S14 and estimated parameters in Table S5. Notice the large values estimated for $T_2$ suggesting that lifting lockdown 1 (since 12 May 2020 in England, and 28 May 2020 in Scotland) had not impacted transmission significantly over the period fitted here. Large values of $T_2$ imply wide confidence intervals for this parameter and large uncertainly. These results led us to repeat the fittings for two particular cases: (a) $c(t)$ remains at lockdown level until the end of the fitting period (i.e., $T_2 \to \infty$, motivated by these preliminary results and by government stringency indices [Hale *et al.* 2020]); or (b) the slope of contact reactivation is such that $c(t)$ is back to baseline ($c(t) = 1$) in 120 days (inspired in Google mobility patterns [Google 2020]). These scenarios are shown in the main text. Although the analysis presented here suggests that (a) is closer to reality we also include (b) to allow assessment of how the estimation of other parameters might depend on assumptions about $T_2$.

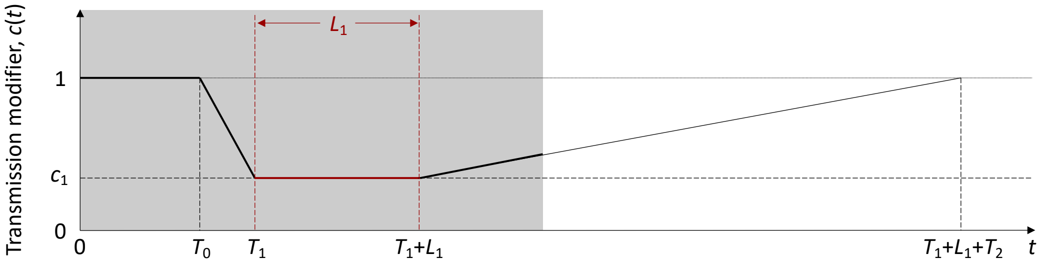


Figure S13: Schematic illustration of the factor $c(t)$, representing the effect of NPIs and adaptive behavioural changes on transmission. $L_1$ represents the known duration (and timing) of the first lockdown as imposed by the governments. The grey area illustrates the time period included in the analysis. $T_0$ is the number of days in the series prior to the gradual contact reductions early in the pandemic (estimated). $T_1 > T_0$ is the day the first lockdown begins. $T_2$ is the number of days for the ramp of increasing transmission after first lockdown to reach $c(t) = 1$ (technically this is estimated and used to define the slope of the linear increase rather than to imply it will continue to follow the trend beyond the study period).

Table S5: Model parameters estimated by Bayesian inference based on daily deaths until 1 July 2020. Model selection based on maximum log-likelihood (LL) and Akaike information criterion (AIC). Best fitting models have lower AIC scores. Infection fatality ratio $\phi = 0.9\%$. Herd immunity threshold ($\mathcal{H}$) calculated from $\mathcal{R}_0$ and $\nu$ using main text formulas (14) or (22) as appropriate. $T_0$ and $T_2$ parameterise linear reduction and increase in transmissibility, respectively, before and after first lockdown (larger $T \Leftrightarrow$ lower slope; main text and Fig. S13).

| | Heterogeneous susceptibility | | Heterogeneous connectivity | | Homogeneous | |
|---|---|---|---|---|---|---|
| | Median | 95% CI | Median | 95% CI | Median | 95% CI |
| *Common parameters* | | | | | | |
| $c_1$ [a] | 0.25 | $(0.23, 0.27)$ | 0.25 | $(0.23, 0.27)$ | 0.23 | $(0.22, 0.24)$ |
| $\eta$ [b] | 15 | $\{15\}$ | 15 | $\{15\}$ | 15 | $\{15\}$ |
| $\nu$ [c] | 1.42 | $(0.96, 1.81)$ | 0.99 | $(0.43, 1.28)$ | 0 | – |
| *England* | | | | | | |
| $T_0$ | 0.65 | $(0.03, 2.30)$ | 0.61 | $(0.03, 2.77)$ | 2.27 | $(0.23, 4.45)$ |
| $T_2$ | $1.99 \cdot 10^5$ | $(1.19 \cdot 10^3, 7.00 \cdot 10^7)$ | $7.42 \cdot 10^5$ | $(1.63 \cdot 10^3, 7.95 \cdot 10^7)$ | $9.78 \cdot 10^5$ | $(4.45 \cdot 10^3, 8.08 \cdot 10^7)$ |
| $\mathcal{R}_0$ | 3.43 | $(3.31, 3.49)$ | 3.43 | $(3.28, 3.49)$ | 3.31 | $(3.16, 3.48)$ |
| $\mathcal{H}$ | 34% | $(24\%, 48\%)$ | 34% | $(24\%, 60\%)$ | 70% | $(68\%, 71\%)$ |
| *Scotland* | | | | | | |
| $T_0$ | 12.50 | $(10.87, 12.98)$ | 12.48 | $(10.78, 12.98)$ | 12.45 | $(10.34, 12.98)$ |
| $T_2$ | $1.28 \cdot 10^5$ | $(3.55 \cdot 10^2, 6.35 \cdot 10^7)$ | $4.35 \cdot 10^5$ | $(4.09 \cdot 10^2, 6.84 \cdot 10^7)$ | $3.82 \cdot 10^5$ | $(5.83 \cdot 10^2, 7.25 \cdot 10^7)$ |
| $\mathcal{R}_0$ | 3.37 | $(3.31, 3.46)$ | 3.37 | $(3.31, 3.46)$ | 3.37 | $(3.30, 3.51)$ |
| $\mathcal{H}$ | 33% | $(25\%, 47\%)$ | 34% | $(24\%, 60\%)$ | 70% | $(70\%, 71\%)$ |
| *Model selection* | | | | | | |
| LL | $-900.4$ | | $-899.6$ | | $-909.9$ | |
| AIC | 1819 | | **1817** | | 1836 | |

[a] transmissibility reduction due to lockdown 1,

[b] difference between mean-time-to-death and mean-time-to-recovery (sampled from a continuous interval and reduced to the nearest integer before entering the model),

[c] coefficient of variation (CV).

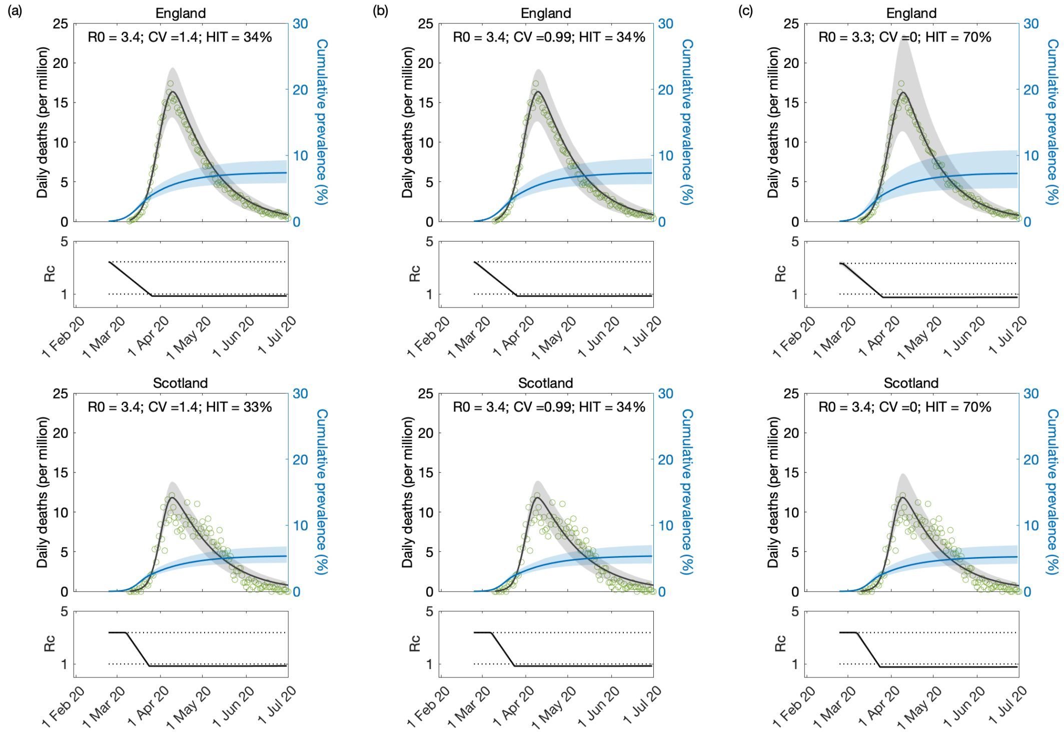


Figure S14: **Model fitting to first wave of the SARS-CoV-2 pandemic.** Modelled trajectories of COVID-19 deaths (black) and cumulative percentage infected (blue). Dots are data for daily reported deaths. Basic reproduction numbers under control ($\mathcal{R}_c$) displayed in shallow panels underneath the main plots. Input parameters: progression from $E$ to $I$ ($\delta = 1/5.5$ per day); recovery ($\gamma = 1/4$ per day); relative infectiousness between $E$ and $I$ stages ($\rho = 0.5$); and IFR ($\phi = 0.9\%$). Initial basic reproduction numbers, coefficients of variation and control parameters estimated by Bayesian inference (estimates in Table S5). Fitted curves represent best fitting trajectories and shades are 95% credible intervals generated from 10,000 posterior samples. (a) Individual variation in susceptibility to infection (main text model (9)-(12)). (b) Individual variation in exposure to infection (main text model (17)-(20)). (c) Homogeneous model (either model with $\nu = 0$).

### S5 Sensitivity to distribution type

To assess sensitivity to the type of distribution, in Fig. S15 we show lognormal fits to the contact survey data reviewed in the main text (Section 5.4.1). These fits are notably worse than those with gamma distributions (main text Fig. 11).

Despite the fits to survey data being worse, for completeness we include a Fig. S16 with the HIT (grey) and epidemic final size (EFS, in blue) adopting the general model (Eqs. (1)-(5) in main text, or (S1)-(S4) in Section S1) with lognormal distributions. These curves were approximated numerically, from the infinite system of ODEs, as the percentage of the population no longer susceptible when infection peaks (i.e., when $d(E+I)/dt = 0$) for HIT, and when infection rates are down to approximately zero for EFS. In the figure we also represent empirical

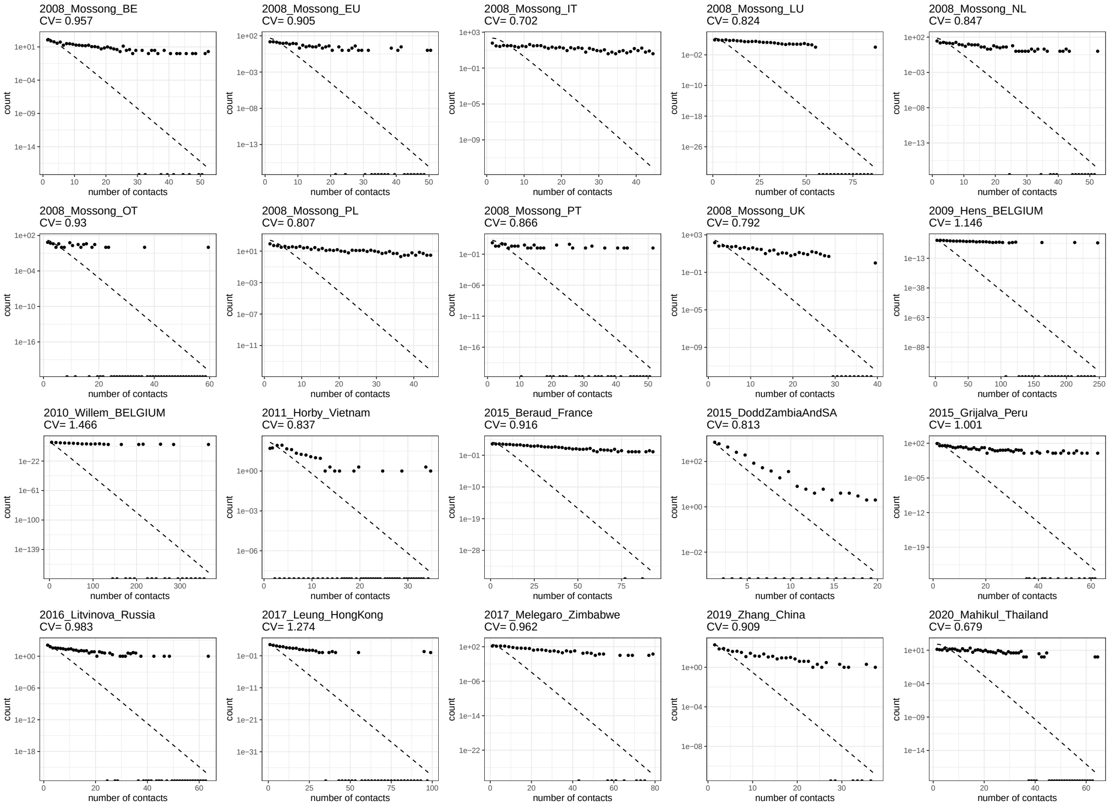


Figure S15: **Lognormal fits for the included contact surveys**. For each dataset in main text Table 5, we plot the empirical distribution and report its CV, as well as the best-fit lognormal distribution.

estimates of CV, obtained from the contact surveys reviewed in Section 5.4.1 (raw data used in Figs. 9 and S15) (solid orange vertical lines) and contact tracing (dashed vertical lines, SARS-CoV-1 in red and SARS-CoV-2 in orange) as described in Section 5.4.2. Comparing this figure with the gamma distribution version (main text, Fig. 14), we note that HIT and EFS curves are basically identical with variable connectivity while the decline of both metrics with CV becomes less steep with variable susceptibility. Model-based estimates for CV assuming lognormal distributions have not been conducted and hence their omission in Fig. S16.

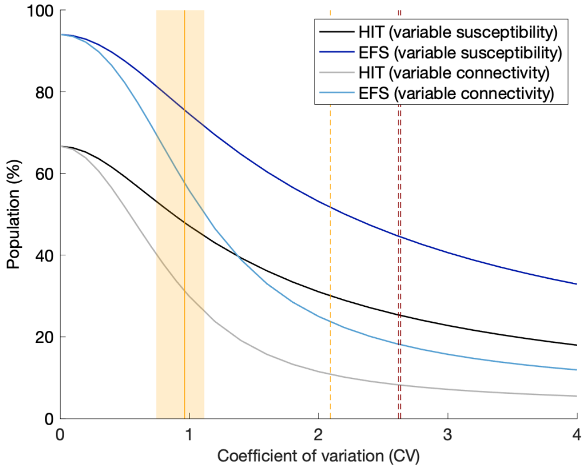


Figure S16: **Herd immunity threshold and epidemic final size with lognormal distributed susceptibility and exposure to infection.** Curves generated using model (1)-(5) in main text ((S1)-(S5) in Section S1) with approximate $\mathcal{R}_0 = 3$: herd immunity threshold (black and grey); final size of unmitigated epidemics (blue). Vertical lines indicate coefficients of individual variation from the literature: connectivity (solid orange) (mean 0.93, standard deviation 0.19, as reviewed in main text Section 5.4.1); infectivity for SARS-CoV-2 (dashed orange) (Hong Kong 2.09 [Adam *et al.* 2020]); infectivity for SARS-CoV-1 (dashed red) (Singapore 2.62, Beijing 2.64 [Lloyd-Smith *et al.* 2005]).

### References

[Adam *et al.* 2020] Adam, D. C., Wu, P., Wong, J. Y., Lau, E. H. Y., Tsang, T. K., Cauchemez, S., et al. (2020) Clustering and superspreading potential of SARS-CoV-2 infections in Hong Kong. *Nat. Med.*, **26**, 1714-1719.

[Bernal *et al.* 2021] Bernal, J. L., Andrews, N., Gower, C., Gallagher, E., Simmons, R., Thelwall, S., et al. (2021) Effectiveness of Covid-19 Vaccines against the B.1.617.2 (Delta) Variant. *N. Engl. J. Med.*, **385**, 585-594.

[Chen *et al.* 2021] Chen, S., Flegg, J. A., White, L. J. and Aguas, R. (2021) Levels of SARS-CoV-2 population exposure are considerably higher than suggested by seroprevalence surveys. *PLOS Comput. Biol.*, **17**, e1009436.

[Gomes *et al.* 2016] Gomes, M. G. M., Gjini, E., Lopes, J. S., Souto-Maior, C. and Rebelo, C. (2016) A theoretical framework to identify invariant thresholds in infectious disease epidemiology. *J. Theor. Biol.*, **395**, 97-102.

[Gomes *et al.* 2004] ]Gomes, M. G. M., White, L. J. and Medley, G. F. (2004) Infection, reinfection, and vaccination under suboptimal immune protection: Epidemiological perspectives. *J. Theor. Biol.*, **228**, 539-549.

[Google 2020] Google (2020) COVID-19 Community Mobility Reports.

[Hale *et al.* 2020] Hale, T., Webster, S., Petherick, A., Phillips, T. and Kira, B. Oxford covid-19 government response tracker (2020) Blavatnik School of Government, 25.

[Hall *et al.* 2021] Hall, V. J., Foulkes, S., Charlett, A., Atti, A., Monk, E. J., Simmons, R., et al. (2021) SARS-CoV-2 infection rates of antibody-positive compared with antibody-negative health-care workers in England: a large, multicentre, prospective cohort study (SIREN). *Lancet*, **397**, 1459-1469.

[Lloyd-Smith *et al.* 2005] Lloyd-Smith, J. O., Schreiber, S. J., Kopp, P. E. and Getz., W. M. (2005) Superspreading and the effect of individual variation on disease emergence. *Nature*, **438**, 355-359.

[Montalbán *et al.* 2020] Montalbán, A., Corder, R. M. and Gomes, M. G. M. (2020) Herd immunity under individual variation and reinfection. *Preprint arXiv:2008.00098v2.*

[Rodrigues *et al.* 2016] Rodrigues, P., Margheri, A., Rebelo, C. and Gomes, M. G. M. (2016) Heterogeneity in susceptibility to infection can explain high reinfection rates. *J. Theor. Biol.*, **259**, 280-290.

[Sheikh *et al.* 2021] Sheikh, A., Robertson, C. and Taylor, B. (2021) BNT162b2 and ChAdOx1 nCoV-19 Vaccine Effectiveness against Death from the Delta Variant. *N. Engl. J. Med.*, **385**, 2195-2197.

[Voysey *et al.* 2020] Voysey, J. L., Clemens, S. A. C., Madhi, S. A., Weckx, L. Y., Folegatti, P. M., Aley, P. K., et al. (2020) Safety and efficacy of the ChAdOx1 nCoV-19 vaccine (AZD1222) against SARS-CoV-2: an interim analysis of four randomised controlled trials in Brazil, South Africa, and the UK. *Lancet*, **397**, 99-111.

[Ward *et al.* 2021] Ward, H., Atchison, C., Whitaker, M., Ainslie, K. E. C., Elliott, J., Okell, L., et al. (2021) SARS-CoV-2 antibody prevalence in England following the first peak of the pandemic. *Nat. Commun.*, **12**, 905.
